# Developmental Associations Linking Childhood Trauma and Early Cannabis Use to Adolescent DNA Methylation and Psychotic-Like Experiences

**DOI:** 10.64898/2026.06.09.26355257

**Authors:** Giulia Trotta, Zhonghua Liu, Isabelle Austin-Zimmerman, Edoardo Spinazzola, Lucia Sideli, Monica Aas, Victoria Rodriguez, Zhikun Li, Perry BM Leung, Qiang Li, Shengmin Zhang, Pak C Sham, Evangelos Vassos, Richard Bentall, Emma Walker, Emma Dempster, Robin M Murray, Marta Di Forti, Luis Alameda, Chloe CY Wong

## Abstract

**Background:** Psychotic-like experiences (PLEs) index early risk for psychotic disorders and are consistently associated with childhood trauma, yet underlying biological mechanisms remain poorly understood. DNA methylation (DNAm) may capture the biological embedding of early adversity, while adolescent exposures such as cannabis use may modify these processes. We examined epigenome-wide associations of childhood trauma and PLEs, tested the moderating role of early cannabis use, and evaluated DNAm as a potential mediator.

**Methods:** We analysed data from the Avon Longitudinal Study of Parents and Children (ALSPAC), a UK population-based birth cohort. Childhood trauma was assessed prospectively and retrospectively. Epigenome-wide DNAm was measured in peripheral blood at ∼17 years using the Illumina 450K array, and PLEs were assessed at 18 using a structured interview. Epigenome-wide association studies were conducted for trauma-DNAm and DNAm-PLEs associations in the final sample (*n =* 1,457), adjusting for demographic, biological, and technical covariates. Differentially methylated regions (DMRs) were identified using DMRff, followed by functional enrichment analyses. Cannabis use at 15.5 was modelled as a moderator with multiple imputation for missing data. Mediation was tested using the Divide-Aggregate Composite-null Test (DACT).

**Results:** Childhood trauma was associated with widespread DNAm differences, primarily at the regional level, with enrichment in pathways related to cellular stress responses. In contrast, DNAm associated with PLEs was more limited and implicated loci involved in epigenetic regulatory processes. These signatures were largely distinct, and there was no evidence supporting mediation after multiple testing correction. Incorporating cannabis use altered the pattern and extent of DNAm associations, with stronger and more significant signals observed at both CpG and regional levels, although these did not translate into evidence of mediation.

**Conclusion:** Childhood trauma and PLEs show distinct DNAm signatures in adolescence, with trauma-related DNAm reflecting broad stress-related processes and PLE-associated DNAm implicating regulatory mechanisms. We found little evidence that DNAm mediates the trauma-PLE association. Instead, adolescent exposures, particularly cannabis use, may distinctly influence trauma-related epigenetic variation with limited detectable downstream effects on PLEs. These findings support a context-dependent model of epigenetic risk and highlight the need for larger longitudinal studies to clarify causal pathways linking early adversity to psychosis.

## Introduction

Psychotic disorders are severe and disabling mental health conditions that typically emerge in late adolescence or early adulthood and are associated with substantial personal, and socio-economic burden ^1^. A growing body of evidence supports a dimensional view of psychosis, by which subclinical psychotic like experiences (PLEs), such as hallucinations and paranoia, are relatively common in the general population and lie on a continuum with clinical psychotic disorders ^2^. Importantly, PLEs are associated with increased risk of later psychosis, poorer mental health outcomes, and functional impairment, making them a valuable phenotype for investigating early aetiological mechanisms in population-based cohorts ^3,4^.

Among environmental risk factors for psychosis, childhood trauma is one of the most consistently replicated. Meta-analytic evidence indicates that exposure to abuse, neglect, or adverse childhood experiences is associated with a two– to three-fold increased risk of psychotic outcome. These associations show a dose-response relationship and persist after adjustment for a wide range of confounders, suggesting a potentially causal role ^5–7^. However, the biological mechanisms through which early-life adversity becomes biologically embedded and gives long-term vulnerability to psychosis remain only partially understood.

Epigenetic processes, particularly DNA methylation (DNAm), have been proposed as a mechanism linking environmental exposures to long-lasting changes in gene regulation without altering DNA sequence ^8^. DNAm is sensitive to environmental influences across development, including early-life stress and substance use, and may act as a molecular mediator between exposure and later mental health outcomes ^9^. A recent case-control study in the EU-GEI sample reported that DNAm differences partially mediated the association between childhood trauma and first-episode psychosis caseness, providing initial evidence that epigenetic changes may link trauma to psychosis ^10^. However, this study was conducted in a cross-sectional clinical sample, which may confound epigenetic signals and thus limit causal inference. A replication in longitudinal, population-based cohorts and at earlier stages of the psychosis spectrum is therefore needed.

In addition to childhood trauma, cannabis use represents another well-established environmental risk factor for psychosis ^11^. Epidemiological studies consistently demonstrated associations between cannabis use, particularly early onset, frequent, and high potency use, and increased risk of psychotic outcomes ^12,13^. Evidence indicated that individuals exposed to early adversity are more likely to initiate cannabis use earlier and to use it more heavily ^14,15^. This raises the possibility that cannabis exposure may modify biological responses, such as changes in DNAm, to childhood trauma further influencing individual lability for psychosis ^16^.

The present study is the first to investigate the epigenetic correlates of childhood trauma and PLEs across development in a population-based cohort. We further tested whether early adolescent cannabis use moderates the association between childhood trauma and DNAm patterns. Finally, we assessed the potential mediating role of DNAm in linking childhood trauma to later PLEs.

## Methods and Materials

### Study design and participants

Analyses were conducted using data from the Avon Longitudinal Study of Parents and Children (ALSPAC), an ongoing population-based birth cohort based in the UK ^17,18^. ALSPAC initially recruited 14,541 pregnant women residing in Avon, with expected delivery dates between 1 April 1991 and 31 December 1992 (Phase I enrolment), resulting in 13,988 children alive at one year of age. Additional recruitment phases conducted when the index children were aged seven years or older (Phases II–IV) increased the total sample to 15,447 pregnancies, corresponding to 14,901 children alive at one year for analyses from age seven onwards. ALSPAC provides extensive prospective and retrospective data on environmental exposures, biological measures, and mental health outcomes. For the present study, participants were included if they had available genome-wide DNAm data during adolescence (approximately 17 years), completed assessments of childhood trauma exposure measured prospectively throughout childhood and adolescence and retrospectively in early adulthood (approximately age 22), and data on PLEs assessed at 18 years ^19^. Study data were collected and managed using REDCap electronic data capture tools hosted at the University of Bristol ^20^. REDCap (Research Electronic Data Capture) is a secure, web-based software platform designed to support data capture for research studies. Ethical approval for ALSPAC was obtained from the ALSPAC Ethics and Law Committee and the Local Research Ethics Committees. Consent for biological samples was collected according to the Human Tissue Act (2004), and informed consent for questionnaire and clinic data was obtained following committee recommendations. Further information about the cohort is available at http://www.bristol.ac.uk/alspac/. Please note that the study website contains details of all the data that is available through a fully searchable data dictionary and variable search tool.

### Measures of childhood trauma

Childhood trauma was assessed prospectively across childhood and adolescence, and retrospectively in early adulthood, using harmonised measures derived from repeated questionnaire and interview data collected within ALSPAC ^21^. The primary exposure was a cumulative trauma score covering ages 0-17 years, sensitivity analyses examined developmentally specific trauma exposures using: early trauma (ages 5-10 years); and late trauma (ages 11-17 years). Both indices represented cumulative counts of traumatic experiences occurring within the specified age windows. Traumatic experiences included emotional abuse, physical abuse, sexual abuse, domestic violence, bullying, and emotional neglect. In additional sensitivity analyses, we examined abuse-only trauma (0-17 years), restricted to emotional, physical, and sexual abuse, to assess whether associations were driven specifically by interpersonal abuse rather than broader trauma exposures. All trauma indices were analysed as continuous count variables, reflecting dose-response relationships between cumulative exposure and outcomes ^22^.

### Measures of cannabis use

Cannabis use was assessed via self-report questionnaire administered at approximately age 15.5 years. Participants were asked whether they had ever used cannabis, and among those reporting use, their frequency of use. Based on these responses, we derived a three-level cannabis frequency variable: never use, occasional use, and frequent use. Occasional use was defined as lower-frequency use categories, whereas frequent use was defined as higher-frequency use categories.

### Measure of PLEs

PLEs were assessed at age 18 using the Psychotic-Like Symptoms Interview (PLIKSi), a structured, interviewer-rated measure of hallucinations, delusions, and thought interference ^23^. PLEs were coded as binary outcome (yes/no), where “yes” included both suspected and definite experiences of at least one symptom.

### Genome-wide DNA methylation measurement

Genome-wide DNAm was measured using peripheral whole blood samples collected during adolescence (ages approximately 17 years) as part of the ALSPAC Accessible Resource for Integrated Epigenomic Studies (ARIES) ^24^. DNAm profiling was performed at 485,577 CpG sites using the Illumina Infinium HumanMethylation450 BeadChip microarray (Illumina, San Diego, CA, USA) ^25^. Laboratory procedures and preprocessing were conducted in accordance with established ALSPAC protocols, as described previously ^26^. For the present analyses, quality control procedures were applied at both the sample and probe level to remove low-quality samples and unreliable probes. These procedures included filtering based on detection p-values, bead count thresholds, and exclusion of probes overlapping known single nucleotide polymorphisms (SNPs). Full details of DNAm preprocessing and quality control are provided in the Supplementary Methods, Section 1.1. For the present analyses, DNAm levels were analysed as M-values, obtained from β-values using a logit transformation.

### Statistical analyses

All analyses were conducted using R (version 4.3.1) and restricted to participants with DNAm in adolescence (ages approximately 17 years) and PLEs at age 18 ^27^. Descriptive statistics were used to characterise the analytic sample. EWAS analyses were conducted to examine associations between childhood trauma, adolescent DNAm, and PLEs at age 18. Two sets of EWAS models corresponding to the mediation framework were estimated: (1) *Path A* – DNAm levels at each CpG site were regressed on childhood trauma exposure; (2) *Path B* – PLEs were regressed on DNAm level at each CpG site, adjusting for childhood trauma. All models were adjusted for a common set of technical and biological covariates, which include age at DNAm assessment, sex, maternal education as a proxy for socioeconomic status (SES), estimated blood cell proportions, smoking score, technical batch effects (plate), and the first five DNAm PCs (mPCs). mPCs were used to account for latent biological and technical variation in the methylation data. Estimated blood cell proportions were derived using established algorithms implemented within the ALSPAC ARIES resource. Multiple testing was addressed using false discovery rate (FDR) correction across all tested CpG sites ^28^. A predefined suggestive discovery threshold (p < 5 x 10^−5^) was also used and probes falling between the two thresholds were also discussed. For visualisation purposes (i.e., Manhattan plots), a Bonferroni-corrected threshold was additionally displayed to aid interpretation, but statistical inference was based on FDR correction. In addition to single-CpG analyses, differentially methylated region (DMR) analyses were performed using the DMRff approach, which aggregates information across spatially correlated CpG sites while accounting for probe-level association statistics and correlation structure ^29^. Regions were defined using a maximum gap of 500 base pairs between neighbouring CpGs and an initial CpG inclusion threshold of p < 0.05. Region-level p-values were corrected for multiple testing using FDR. Functional enrichment analyses were conducted on genes annotated to identified DMRs to explore overrepresented biological processes and pathways ^30^. To explore potential effect modification, we tested whether cannabis use frequency at age 15.5 years moderated associations between childhood trauma and DNAm. Cannabis use was modelled as three-level ordinal variable (i.e., never use, occasional use, frequent use). Missing data on cannabis frequency were imputed using multiple imputation by chained equations (MICE), with an ordinal logistic regression model ^31^. The imputation model included earlier cannabis-related measures, sex, smoking score, trauma exposure, and PLEs. Finally, exploratory mediation analyses were conducted using the Divide-Aggregate Composite-null Test (DACT) to assess whether DNAm mediated the association between childhood trauma and PLEs ^32^. This test has been developed to test mediation effects in EWAS context and was already used in our previous publication ^22^. Roughly, it integrates evidence from Path A and Path B models to identify CpG sites for which both associations are simultaneously non-zero. Composite mediation p-values were computed for each CpG site and corrected for multiple testing using FDR. Separate mediation analyses were conducted for different operationalisations of childhood trauma. In addition, exploratory mediation analyses were repeated using cannabis-moderated *Path A* estimates to assess whether incorporating cannabis exposure influenced evidence for DNAm-mediated effects.

## Results

### Sample Characteristics

Among 2,798 participants with DNAm available at age approximately 17 years, 1,457 had complete data on childhood trauma, PLEs, cannabis use data and other covariates, forming the final EWAS analytic sample (**Figure 1**).

**Figure 1.**
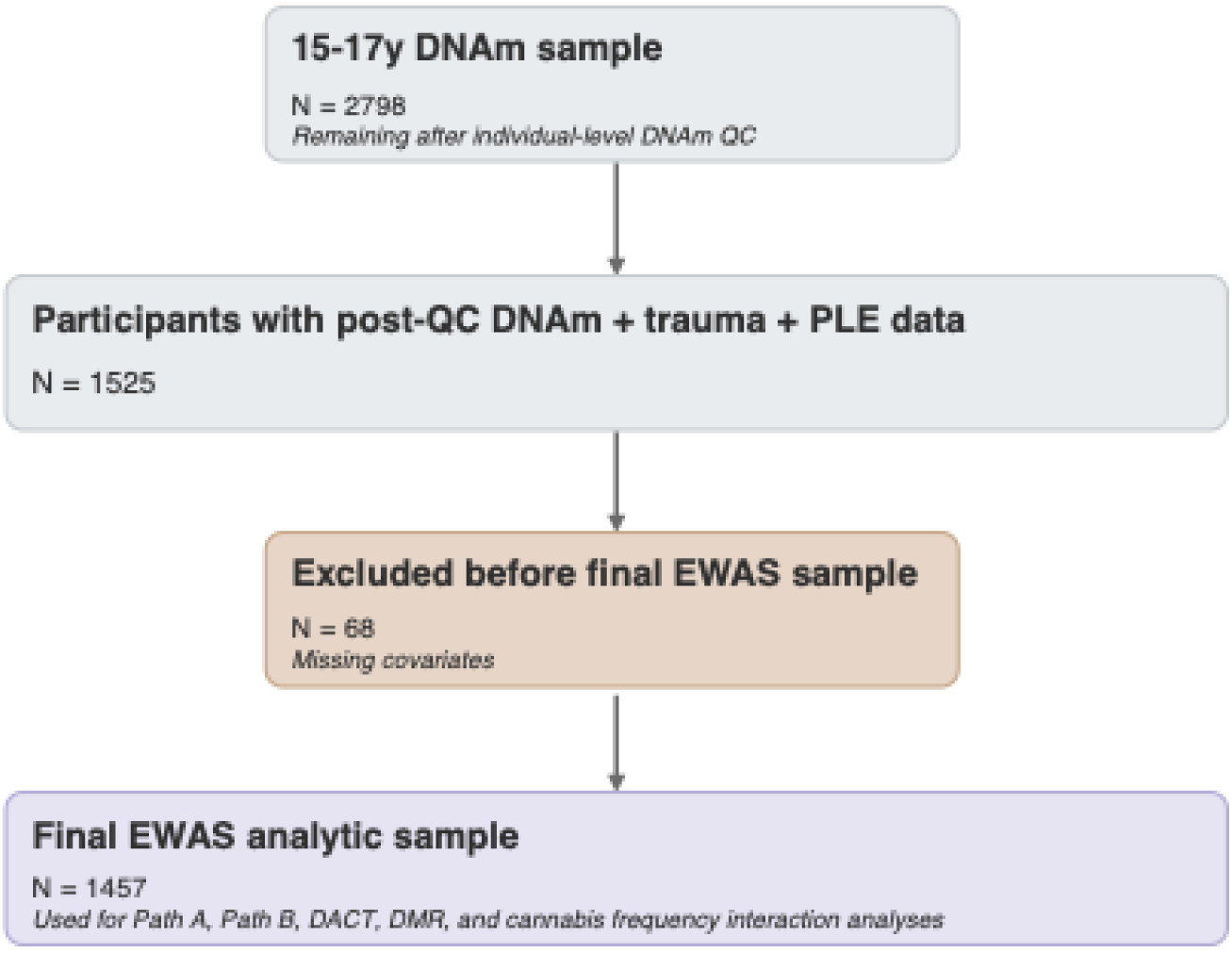
Analytic sample flowchart for EWAS analyses.

In the analytic sample (N = 1,457), the mean age at DNAm assessment was 17.66 years (SD = 0.64), and 52.3% were female. The mean cumulative childhood trauma score (0-17 years) was 1.04 (SD = 1.14). Early adolescent cannabis use was reported by 21.5% of participants. 8.2% of participants experienced at least one PLE (**Table 1**).

**Table 1.**
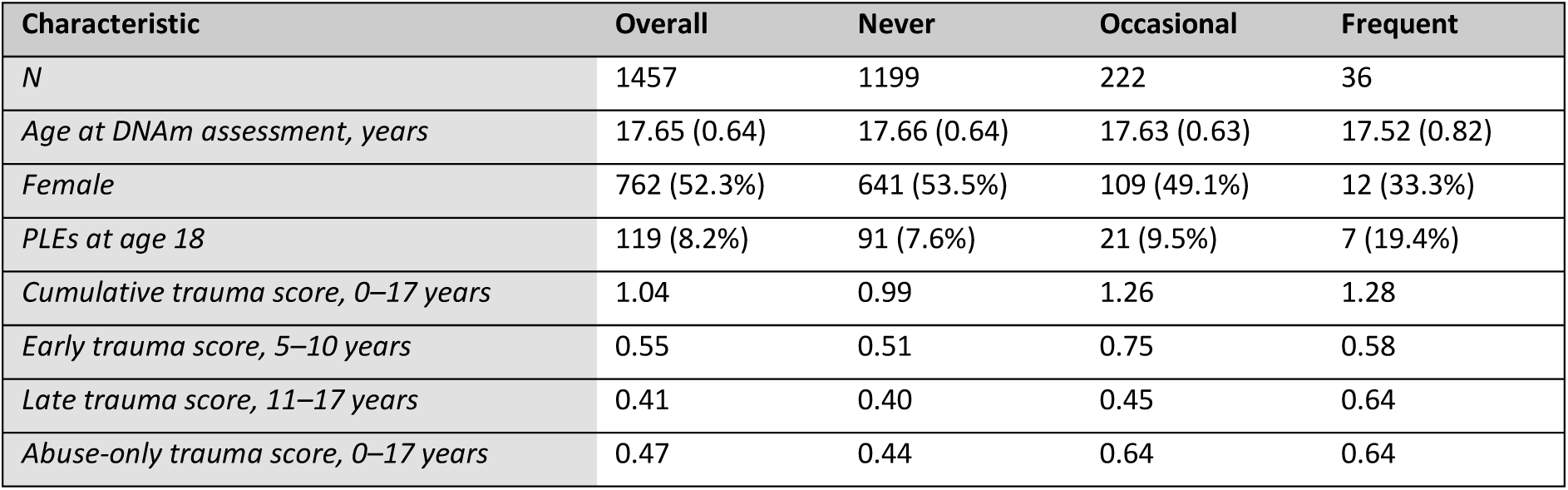
Characteristics of the analytical sample.

| Characteristic | Overall | Never | Occasional | Frequent |
| --- | --- | --- | --- | --- |
| *N* | 1457 | 1199 | 222 | 36 |
| *Age at DNAm assessment, years* | 17.65 (0.64) | 17.66 (0.64) | 17.63 (0.63) | 17.52 (0.82) |
| *Female* | 762 (52.3%) | 641 (53.5%) | 109 (49.1%) | 12 (33.3%) |
| *PLEs at age 18* | 119 (8.2%) | 91 (7.6%) | 21 (9.5%) | 7 (19.4%) |
| *Cumulative trauma score, 0–17 years* | 1.04 | 0.99 | 1.26 | 1.28 |
| *Early trauma score, 5–10 years* | 0.55 | 0.51 | 0.75 | 0.58 |
| *Late trauma score, 11–17 years* | 0.41 | 0.40 | 0.45 | 0.64 |
| *Abuse-only trauma score, 0–17 years* | 0.47 | 0.44 | 0.64 | 0.64 |

### Phenotypic associations between trauma exposure, cannabis use, and PLEs

In phenotypic analyses, adjusted for sex, age, and maternal education, cumulative trauma exposure was significantly associated with increased odds of PLEs (OR = 1.59, 95% CI 1.38-1.84, p < 0.001). Frequent, but not occasional, cannabis use was also significantly associated with increased odds of PLEs (OR = 2.94, 95% CI 1.21-7.14, p < 0.05). There was no evidence that cannabis use modified the association between trauma exposure and PLEs (interaction p-values > 0.3).

### Epigenome-wide association analyses

Differentially methylated probe (DMP) and differentially methylated region (DMR) analyses were conducted separately for Path A and Path B across four trauma definitions (i.e., cumulative trauma 0-17 years, early trauma 5-10 years, late-trauma 11-17 years, abuse-only 0-17 years). In Path A analyses, the most significant differentially methylated probe (DMP) association was observed for early trauma (5-10 years) at *cg16255307* (β = –0.0609, SE = 0.0101, *p* = 2.36 x 10^−09^), whereas associations for cumulative, late, and abuse-only trauma were largely limited to suggestive signals (**Figure 2**). In path B analyses, several DMPs showed recurrent associations based on the suggestive discovery threshold across trauma-adjusted models, including *cg26428054* (β = –3.425, SE = 0.6525, *p* = 1.54 x 10^−07^) and *cg10529789* (β = 2.898, SE = 0.5719, *p* = 4.05 x 10^−07^), particularly in late trauma models (**Figure 3**). Summary of EWAS findings for Path A and Path B are reported in **Table 2**, full results for all trauma definitions are provided in the Supplementary Material.

**Figure 2.**
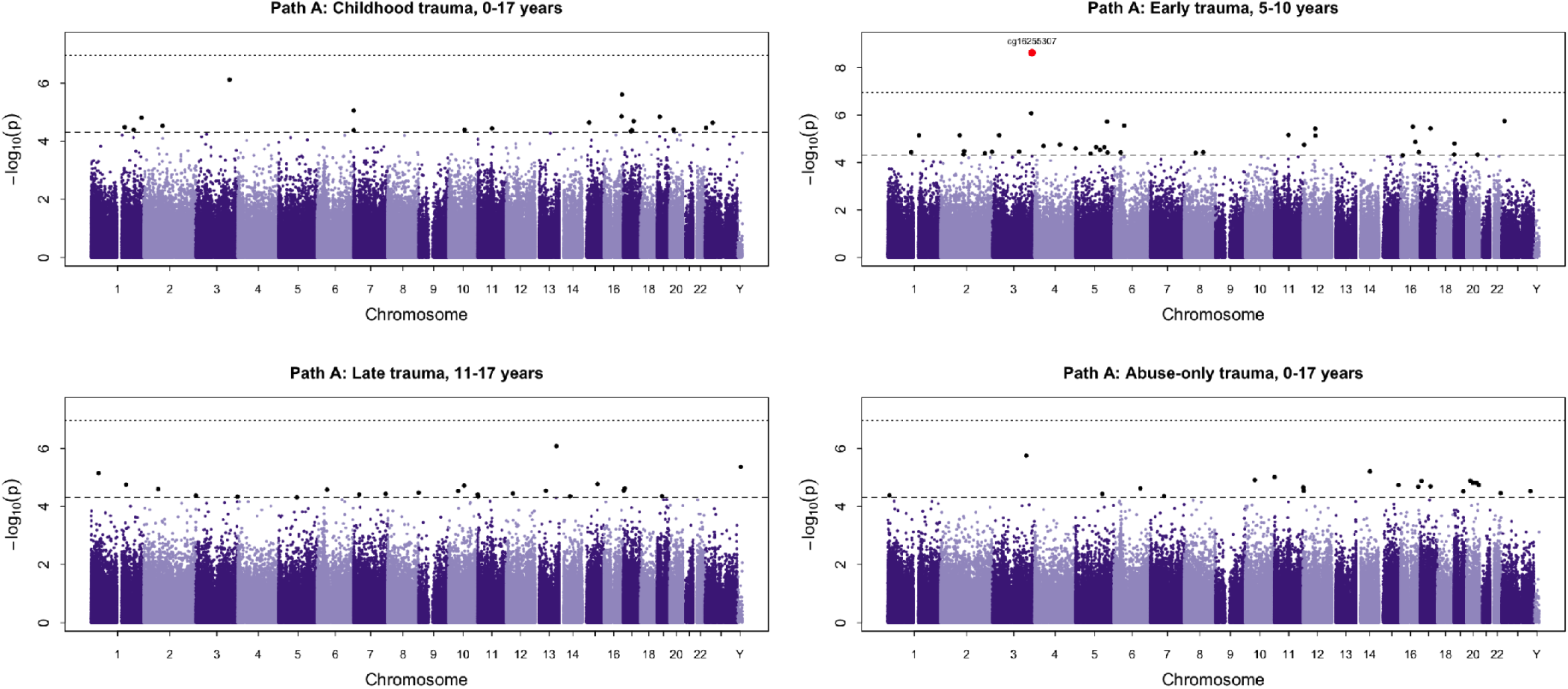
Epigenome-wide association analysis of childhood trauma and DNAm (Path A). Manhattan plot showing epigenome-wide association results between childhood trauma exposure and DNAm (Path A). Each point represents a CpG site plotted according to genomic position across chromosomes and the –log_10_-transformed p-value. The horizontal dashed line indicates a suggestive discovery threshold (p < 5 x 10^−5^), while the dotted line represents the more conservative Bonferroni-corrected threshold.

**Figure 3.**
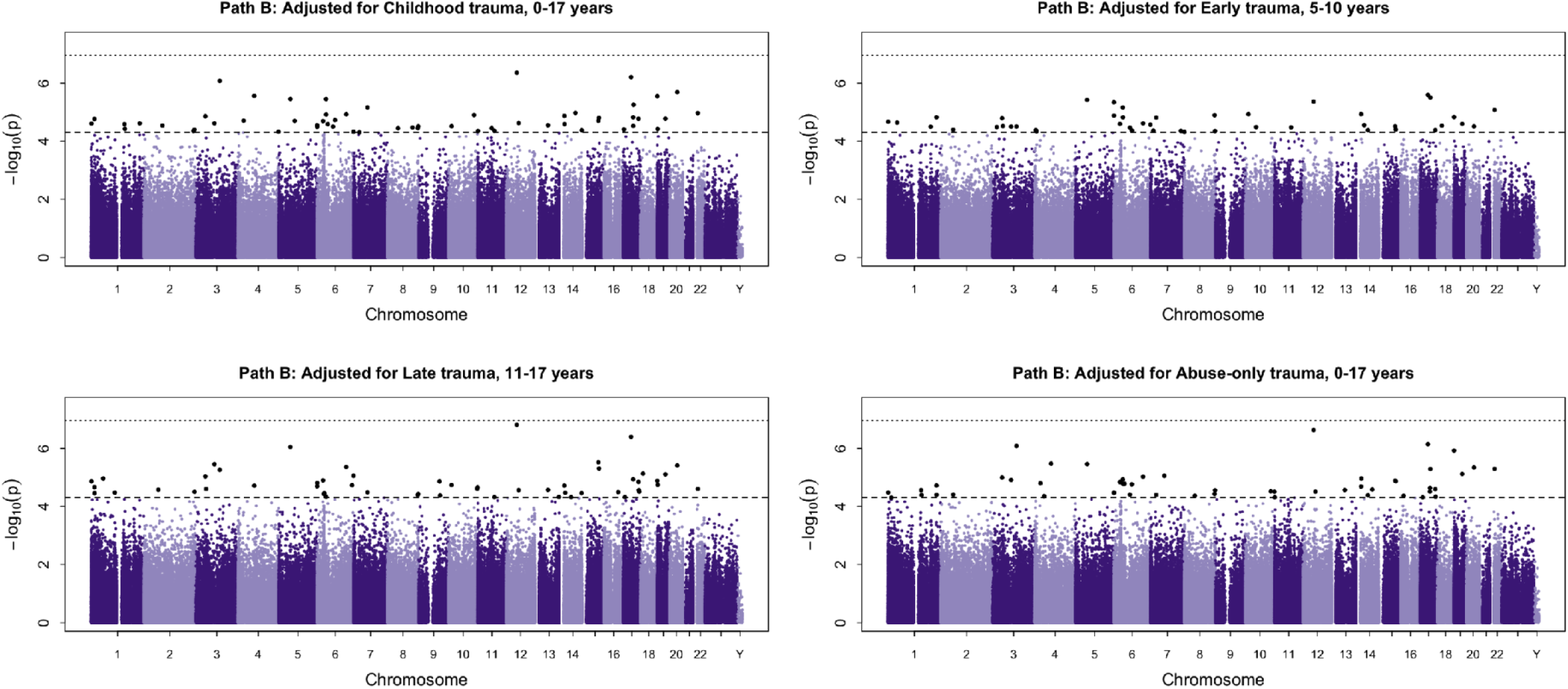
Epigenome-wide association analysis of DNAm and PLEs (Path B). Manhattan plot showing epigenome-wide association results examining the relationship between DNAm and PLEs (Path B), adjusted for childhood trauma. Each point represents a CpG site plotted according to genomic position across chromosomes and the –log_10_-transformed p-value. The horizontal dashed line indicates a suggestive discovery threshold (p < 5 x 10^−5^), while the dotted line represents the more conservative Bonferroni-corrected threshold.

**Table 2.**
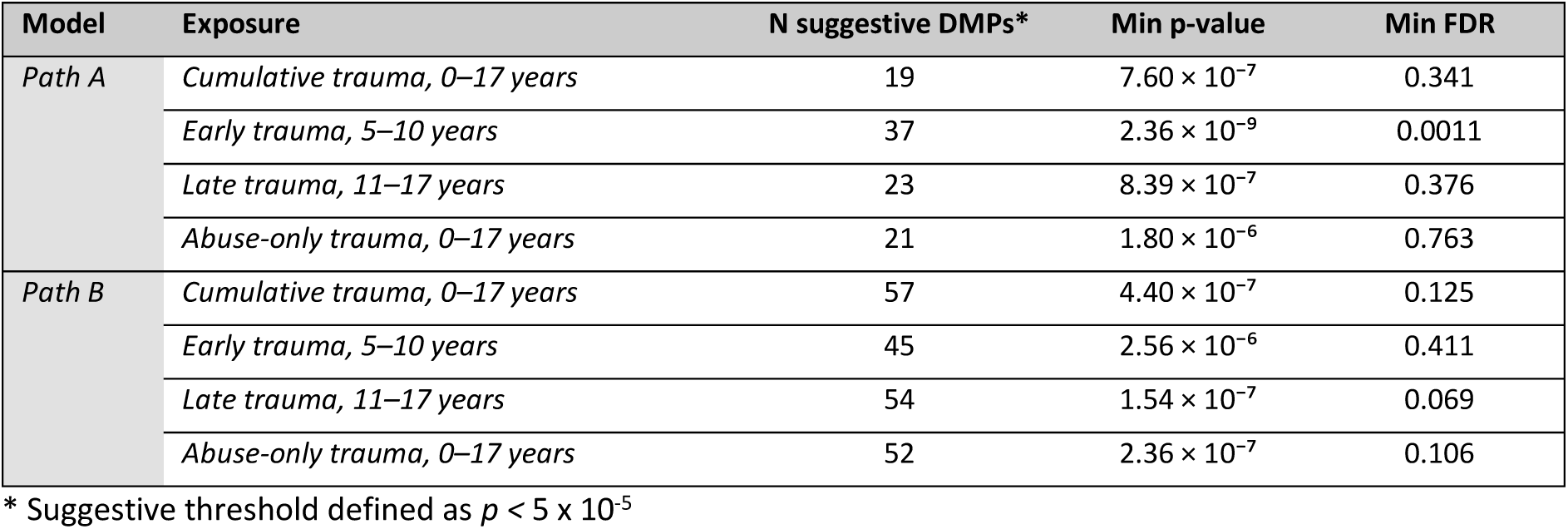
Summary of EWAS results for Path A and Path B analyses.

| Model | Exposure | N suggestive DMPs* | Min p-value | Min FDR |
| --- | --- | --- | --- | --- |
| Path A | Cumulative trauma, 0–17 years | 19 | $7.60 \times 10^{-7}$ | 0.341 |
| | Early trauma, 5–10 years | 37 | $2.36 \times 10^{-9}$ | 0.0011 |
| | Late trauma, 11–17 years | 23 | $8.39 \times 10^{-7}$ | 0.376 |
| | Abuse-only trauma, 0–17 years | 21 | $1.80 \times 10^{-6}$ | 0.763 |
| Path B | Cumulative trauma, 0–17 years | 57 | $4.40 \times 10^{-7}$ | 0.125 |
| | Early trauma, 5–10 years | 45 | $2.56 \times 10^{-6}$ | 0.411 |
| | Late trauma, 11–17 years | 54 | $1.54 \times 10^{-7}$ | 0.069 |
| | Abuse-only trauma, 0–17 years | 52 | $2.36 \times 10^{-7}$ | 0.106 |

\* Suggestive threshold defined as $p < 5 \times 10^{-5}$

Our DMR analyses identified multiple highly significant DMRs associated with childhood trauma exposure across both Path A and Path B models (**Table 3**). Path B models revealed a greater number of DMR signals than Path A models, particularly for cumulative and late trauma exposures. Importantly, the DMRs identified in Path A and Path B analyses were entirely distinct, with no overlap observed between the two pathways.

**Table 3.** Summary of DMR analyses across trauma exposures.

| Path | Exposure | N of significant DMRs | Min p-value | Min FDR |
| --- | --- | --- | --- | --- |
| PathA | Abuse-only trauma, 0-17 years | 2 | $9.125 \times 10^{-10}$ | $4.374 \times 10^{-4}$ |
| PathA | Early trauma, 5-10 years | 5 | $6.909 \times 10^{-18}$ | $3.334 \times 10^{-12}$ |
| PathA | Late trauma, 11-17 years | 2 | $9.987 \times 10^{-10}$ | $4.746 \times 10^{-4}$ |
| PathA | Childhood trauma, 0-17 years | 1 | $5.369 \times 10^{-12}$ | $2.552 \times 10^{-6}$ |
| PathB | Abuse-only trauma, 0-17 years | 5 | $5.611 \times 10^{-11}$ | $2.749 \times 10^{-5}$ |
| PathB | Early trauma, 5-10 years | 7 | $2.033 \times 10^{-12}$ | $9.897 \times 10^{-7}$ |
| PathB | Late trauma, 11-17 years | 10 | $1.231 \times 10^{-14}$ | $6.035 \times 10^{-9}$ |
| PathB | Childhood trauma, 0-17 years | 7 | $7.217 \times 10^{-13}$ | $3.535 \times 10^{-7}$ |

Table summarising the number of DMRs identified for Path A and Path B across the four trauma exposures. Results are based on region-level significance thresholds derived from the DMRff method.

Notably, several of the most significant DMRs were located within the chromosome 6 major histocompatibility complex (MHC) region, including a region annotated to *EHMT2* in the Path B late trauma model. Additional DMRs were identified across other genomic loci, including regions annotated to *KLHL34* and *DOM3Z/STK19* (Path A early trauma), and *ZDHHC9* (Path B cumulative trauma). Representative DMRs are shown in **Figure 4**.

**Figure 4.**
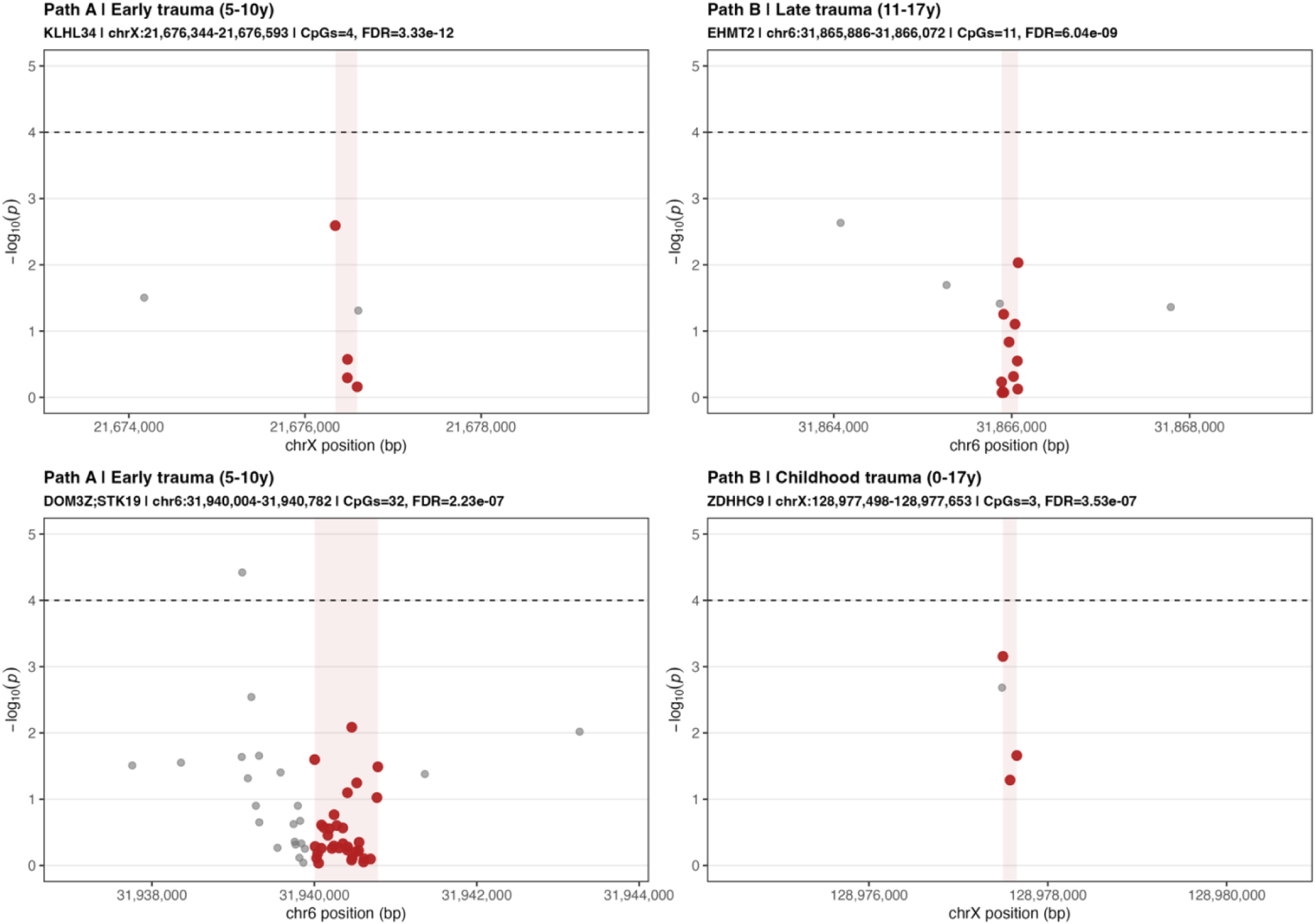
Representative DMRs associated with childhood trauma across Path A and Path B models. Locus-specific plots showing the top-ranked DMRs across Path A and Path B models. Each point represents an individual CpG site, plotted by genomic position (x-axis) and statistical significance (y-axis). CpG sites within the identified DMR are highlighted in red, while surrounding CpGs are shown in grey. The shaded region indicates the genomic span of the DMR. The dashed horizontal line indicates the nominal significance threshold (p = 1 x 10^−4^)

Downstream discovery functional enrichment analyses of genes annotated to DMRs identified a limited number of overrepresented Gene Ontology (GO) terms (Supplementary Tables S6-S7). For Path A, enriched terms showed a consistent pattern related to oxidoreductase activity, protein folding, and endoplasmic reticulum-associated processes. For Path B, enrichment results were characterised by processes related to epigenetic regulation, including histone methylation and chromatin organisation. However, it is worthnoting that enrichment signals were driven by a small number of genes, and no pathways remained significant after multiple testing correction. KEGG pathway analyses similarly did not reveal consistent pathway-level enrichment.

### Cannabis-Moderated Epigenome-Wide Models

We next examined whether associations between childhood trauma and DNAm differed according to frequency of cannabis use. Trauma x cannabis interaction models showed clearer signal at both the single-CpG and regional levels. At the CpG level, interaction analyses identified one FDR-significant locus for cumulative trauma and multiple FDR-significant loci for early, late, and abuse-only trauma, with recurrent signals involving genes such as *TTLL3, MCTP1, PAFAH2,* and *FGF4.* Summary findings are reported in **Table 4**, while full results are presented in the Supplementary Material.

**Table 4.** Summary of EWAS results for cannabis-moderated models.

| Model | Exposure | N suggestive DMPs* | Min p-value | Min FDR |
| --- | --- | --- | --- | --- |
| Cannabis-moderated Path A | Cumulative trauma, 0–17 years | 63 | $1.68 \times 10^{-10}$ | $7.52 \times 10^{-5}$ |
| | Early trauma, 5–10 years | 89 | $4.35 \times 10^{-14}$ | $1.95 \times 10^{-8}$ |
| | Late trauma, 11–17 years | 91 | $1.01 \times 10^{-11}$ | $4.52 \times 10^{-6}$ |
| | Abuse-only trauma, 0–17 years | 82 | $3.28 \times 10^{-10}$ | $1.47 \times 10^{-4}$ |

\* Suggestive threshold defined as $p < 5 \times 10^{-5}$

Regional analyses similarly identified multiple significant DMRs across all four trauma definitions, ranging from 15 to 19 regions per model (**Table 5**). Full results are presented in the Supplementary Material.

**Table 5.** Overview of DMR counts and signals across trauma x cannabis interaction analyses.

| Exposure | Number of DMRs | Min P-value | Min FDR | Top Gene |
| --- | --- | --- | --- | --- |
| Abuse-only trauma, 0-17 years | 15 | $2 \times 10^{-14}$ | $9.7 \times 10^{-9}$ | |
| Early trauma, 5-10 years | 19 | $1.06 \times 10^{-17}$ | $5.13 \times 10^{-12}$ | ZNF674; LOC401588 |
| Late trauma, 11-17 years | 17 | $2.38 \times 10^{-14}$ | $1.15 \times 10^{-8}$ | CNOT2 |
| Childhood trauma, 0-17 years | 17 | $2.69 \times 10^{-13}$ | $1.3 \times 10^{-7}$ | |

The most significant DMRs were annotated to genes including *ZNF674*, *TTLL3* and *CNOT2,* while several top regions in cumulative and abuse-only trauma models were located in unannotated genomic regions (**Figure 5**). Notably, these regions comprised multiple CpG sites showing consistent effects despite limited single-CpG significance.

**Figure 5.**
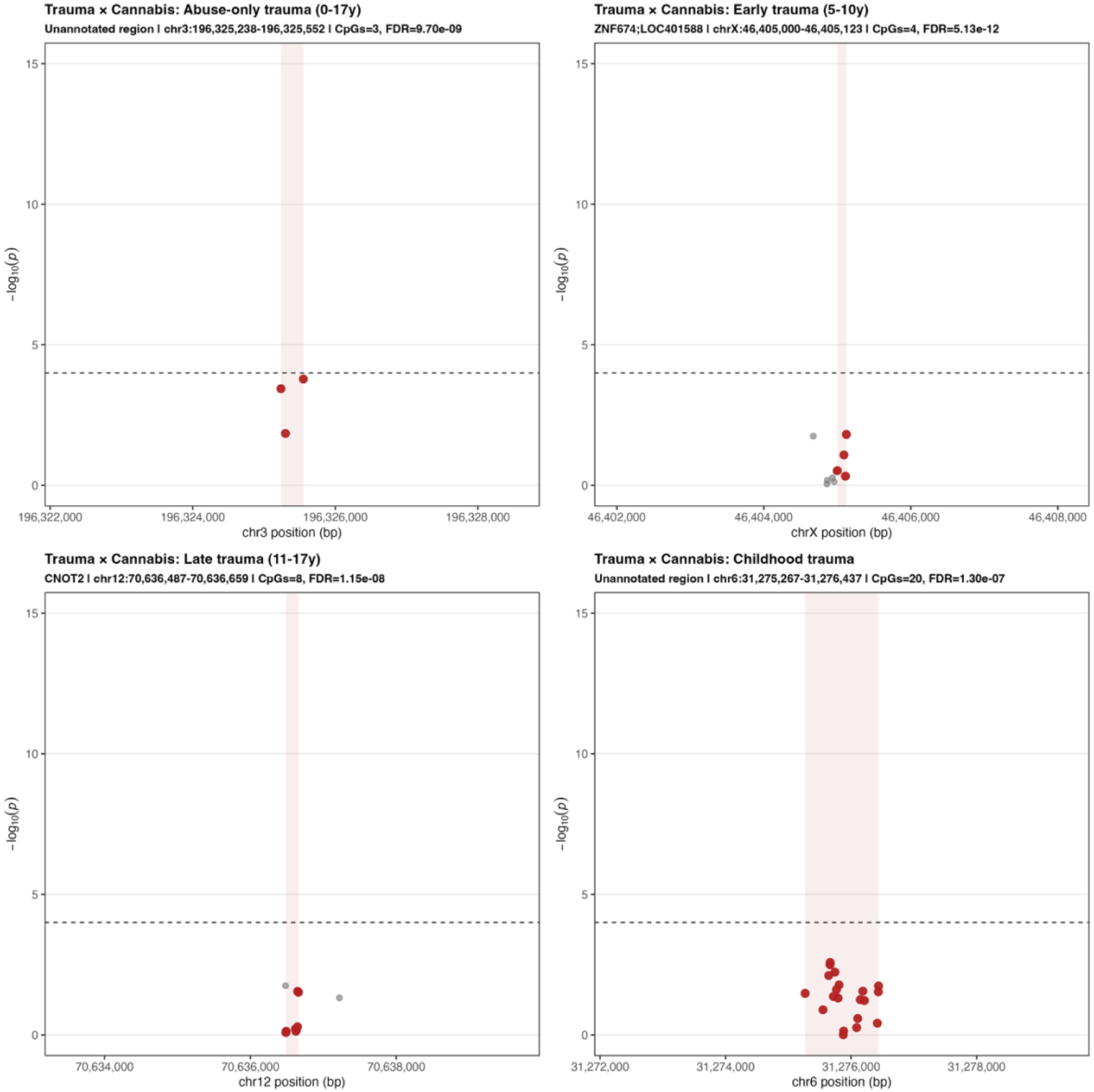
Representative DMRs associated with cannabis-moderated trauma effects. Locus-specific plots showing the top-ranked identified in trauma x cannabis interaction models. Each point represents an individual CpG site, plotted by genomic position (x-axis) and statistical significance (y-axis). CpG sites within the identified DMR are highlighted in red, while surrounding CpGs are shown in grey. The shaded region indicates the genomic span of the DMR. The dashed horizontal line indicates the nominal significance threshold (p = 1 x 10^−4^).

Functional enrichment analyses of genes mapped to DMRs identified in cannabis interaction models yielded a small number of nominally enriched GO terms related to signalling and regulatory processes (Supplementary Table S12). KEGG pathway analyses indicated involvement of signalling-related pathways, including MAPK and calcium signalling pathways, which reflect general processes of cellular signalling, proliferation, and survival. However, enrichment signals were driven by a small number of genes and did not survive correction for multiple testing. Full interaction EWAS and DMR results are presented in the Supplementary Material.

### Epigenome-Wide Mediation Analyses (DACT)

We next examined whether DNAm mediated the association between childhood trauma and PLEs using the DACT framework, integrating evidence from both Path A and Path B analyses. Consistent with the limited probe-level signal observed in EWAS, no CpG sites reached statistical significance after FDR correction. However, a small number of CpG sites showed suggestive evidence of mediation at the predefined discovery threshold across cumulative, early, late, and abuse-only trauma models, but these signals were sparse and did not support robust mediation (**Table 6**). Similar patterns were observed in cannabis-moderated DACT models, where the number of suggestive CpGs was even more limited, providing no additional evidence for mediation.

**Table 6.** Top CpG sites showing suggestive evidence of mediation.

| Exposure | Probe | Genomic location (hg19) | Gene | Location | Mediation effect | P-value | FDR |
| --- | --- | --- | --- | --- | --- | --- | --- |
| Childhood trauma, 0-17 years | cg01821557 | 3:49894282 | TRAIP | S_Shore | 0.0266 | $4.6223 \times 10^{-5}$ | $9.7874 \times 10^{-1}$ |
| Early trauma, 5-10 years | ch.11.1421098F | 11:68338008 | SAPS3 | OpenSea | 0.0318 | $4.4838 \times 10^{-5}$ | $9.7906 \times 10^{-1}$ |
| Late trauma, 11-17 years | cg02078882 | 12:30911709 | | S_Shelf | 0.0504 | $1.2155 \times 10^{-5}$ | $9.8211 \times 10^{-1}$ |
| Late trauma, 11-17 years | cg06602478 | 3:195159296 | ACAP2 | N_Shelf | -0.0582 | $1.2920 \times 10^{-5}$ | $9.8211 \times 10^{-1}$ |
| Abuse-only trauma, 0-17 years | cg01821557 | 3:49894282 | TRAIP | S_Shore | 0.0406 | $2.7658 \times 10^{-5}$ | $9.7637 \times 10^{-1}$ |
| Abuse-only trauma, 0-17 years | cg26838691 | 2:24397539 | C2orf84 | N_Shore | 0.0376 | $4.7194 \times 10^{-5}$ | $9.7637 \times 10^{-1}$ |

Genomic locations are reported according to the hg19 reference genome. Gene annotation and CpG island context correspond to the Illumina HumanMethylation450 array annotation.

## Discussion

This is the first study using a developmental model to examine, firstly, whether childhood trauma is associated with DNAm in adolescence and whether this association is moderated by early cannabis use. Secondly, it examines whether DNAm changes measured in adolescence mediate the relationship between childhood trauma and PLEs at age 18. In the primary EWAS analyses, we observed consistent significant signals at multiple DMRs associated with childhood trauma and PLEs. In contrast, mediation analyses identified no CpG sites that survived multiple testing correction, with only a small number of probes reaching the suggestive threshold. Cannabis-moderated analyses revealed stronger DNAm signals at both the probe and regional levels, suggesting that further environmental exposures in adolescence, such as cannabis use, may modify or amplify trauma-related epigenetic variation.

Childhood trauma was associated with DNAm differences primarily at regional level, with enrichment in pathways related to oxidoreductase activity, protein folding, and endoplasmic reticulum function. These processes are consistent with general cellular stress responses, supporting the hypothesis that early-life adversity becomes biologically embedded through widespread physiological mechanisms ^9^. The lack of robust single-CpG associations suggests that trauma-related DNAm effects are subtle and distributed and may be better captured using region-based approaches. This pattern is consistent with previous EWAS studies of childhood adversity, and with broader evidence indicating that DNAm signatures of trauma are typically small in magnitude and widely distributed across the genome ^33^.

In contrast, DNAm associated with PLEs was characterised by loci involved in epigenetic regulation, including histone methylation and chromatin organisation. A key region was annotated to *EHMT2*, a gene involved in transcriptional repression and chromatin modification, with known relevance to neurodevelopment and psychiatric disorders ^34,35^. Additional loci included *ZDHHC9* and regions within the MHC. Several of these genes have been linked to a range of environmental exposures and complex traits, supporting their potential role in environmentally sensitive biological pathways ^36–38^. Overall, these findings suggest that DNAm variation related to PLEs may reflect alterations in epigenetic regulatory processes. However, compared to trauma-related DNAm, evidence linking DNAm to PLEs and broader mental health outcomes remains relatively limited.

We found no overlap between trauma-associated and PLE-associated DMRs, providing limited support for a direct mediation model. Consistently, mediation analyses (DACT) did not identify any CpG sites that significantly mediated the association between trauma and PLEs. Similar null findings were observed in cannabis-moderated mediation models, where only a very small number of CpGs reached the suggestive threshold, providing no additional support for mediation. This may reflect small effect sizes, limited power, or the use of peripheral blood DNAm as a proxy for brain processes ^39–41^. It is also possible although DNAm signatures are robust biomarkers for xxx, they does not play a major mediating role in this targeted analysis. These findings contrast with previous work in clinical samples, which reported evidence for DNAm mediating the association between childhood trauma and first-episode psychosis ^22^. Differences in sample characteristics (population-based vs. clinical), including the milder and more heterogeneous nature of PLEs compared to clinical psychosis, as well as developmental timing may account for these discrepancies.

Cannabis-moderated analyses identified robust DNAm differences at both the CpG and regional levels, including multiple FDR-significant CpG sites and DMRs across all trauma definitions. These regions were annotated to genes including *ZNF674, TTLL3,* and *CNOT2*, with enrichment in signalling pathways such as MAPK and calcium signalling. These findings suggest that cannabis use may modify trauma-related epigenetic patterns during adolescence. However, given the absence of phenotypic interaction effects, these findings may reflect broader biological responses to cannabis exposure rather than mechanisms specific to psychosis risk ^9,42^.

This study has several strengths. It leverages a well-characterised longitudinal cohort with prospective measures of childhood trauma and of early adolescence cannabis use, genome-wide DNAm data collected in adolescence, and clinically assessed PLEs in early adulthood. The use of multiple trauma definitions allowed for a nuanced examination of developmental timing, and the integration of single-CpG, regional (DMR), and mediation analyses provided a comprehensive assessment of epigenetic associations.

Several limitations should also be considered. First, DNAm was measured in peripheral blood, which may not fully reflect epigenetic processes occurring in brain tissues ^39–41^. However, peripheral tissues are more accessible and enable the collection of larger, deeply phenotyped longitudinal cohorts, as in the present study. In addition, prior work has shown meaningful correspondence between peripheral and brain DNAm patterns, and the identification of multiple significant signals in our analyses supports the validity of this approach for detecting biologically relevant associations. Second, despite the relatively large sample size for an epigenetic study, statistical power may have been insufficient to detect small effect sizes, particularly in mediation analyses, as well as for the interaction models tested ^43–46^. Finally, functional enrichment results did not survive multiple testing correction and were driven by a small number of genes, indicating that further replication and validation will be important in future studies.

In summary, childhood trauma and PLEs were significantly associated with distinct patterns of DNAm in adolescence, with the strongest signals emerging at the regional level across the epigenome. We found limited evidence that DNAm mediated the association between childhood trauma and PLEs, suggesting that these epigenetic alterations may represent parallel or downstream biological correlates rather than direct mechanistic pathways. In contrast, analyses incorporating cannabis use revealed substantially different DNAm profiles at both CpG and regional levels, indicating that adolescent environmental exposures may modify or amplify trauma-related epigenetic signatures. Collectively, these findings emphasise the dynamic and context-dependent nature of epigenetic variation in pathways related to psychosis risk and highlight the importance of large-scale longitudinal studies with repeated DNAm assessments and detailed environmental characterisation to clarify the temporal sequencing, biological specificity, and potential causal relevance of epigenetic mechanisms in the development of psychosis-related outcomes.

## Acknowledgments

We are extremely grateful to all the families who took part in the ALSPAC study, the midwives for their help in recruiting them, and the whole ALSPAC team, which includes interviewers, computer and laboratory technicians, clerical workers, research scientists, volunteers, managers, receptionists and nurses. OpenAI ChatGPT was used to assist with grammar and language editing, and to verify coding syntax; all scientific content and interpretation remain the responsibility of the authors.

## Data availability statement

The informed consent obtained from ALSPAC (Avon Longitudinal Study of Parents and Children) participants does not allow the data to be made available through any third party maintained public repository. Supporting data are available from ALSPAC on request under the approved proposal number, B4002 Full instructions for applying for data access can be found here: http://www.bristol.ac.uk/alspac/researchers/access/. The ALSPAC study website contains details of all available data (http://www.bristol.ac.uk/alspac/researchers/our-data/).

