## Supplementary material for "Developmental Associations Linking Childhood Trauma and Early Cannabis Use to Adolescent DNA Methylation and Psychotic-Like Experiences"

**1. Additional Methods** **4**

*1.1. DNAm preprocessing and quality control* *4*

*1.2. DNAm Principal Component Analysis (mPCs)* *4*

*1.3. Smoking score derivation and validation* *4*
**Figure S1.** *Association between AHRR methylation and nicotine dependence at age 17.* 5

*1.4. Cannabis frequency imputation* *5*

**2. Supplementary Results: Phenotypic Associations** **6**

**Table S1.** *Phenotypic associations with psychotic-like experiences at age 18* 6

**3. EWAS diagnostics** **7**

*3.1. Path A Diagnostics* *7*
**Figure S2.** *QQ plots for EWAS analyses of childhood trauma and DNAm (Path A).* 7

*3.2. Path B Diagnostics* *8*
**Figure S3.** *QQ plots for EWAS analyses of DNAm and PLEs (Path B).* 8

**4. Epigenome-wide Association Results** **9**

*4.1. Path A – Associations between childhood trauma and DNAm* *9*
**Table S2.** *Top CpG sites associated with childhood trauma in EWAS association analyses* 9

*4.2 Path B – Associations between DNAm and PLEs* *13*
**Table S3.** *Top CpG sites associated with PLEs in EWAS association analyses (adjusted for childhood trauma)* 13

**5. Differentially methylated regions (DMR) analyses** **21**

**Table S4.** *Top differentially methylated regions associated with trauma exposure (Path A)* 21
**Table S5.** *Top differentially methylated regions associated with psychotic-like experiences (Path B)* 21
**Figure S4.** *Differentially methylated region (DMR) analyses for Path A.* 23
**Figure S5.** *Differentially methylated region (DMR) analyses for Path B.* 24

**6. Functional Enrichment Analyses** **25**

**Table S6.** *Gene Ontology and KEGG pathway enrichment results for genes annotated to DMRs - Path A* 25
**Figure S6.** *Functional enrichment of trauma-DMRs in Path A.* 27
**Table S7.** *Gene Ontology and KEGG pathway enrichment results for genes annotated to DMRs - Path B* 28
**Figure S7.** *Functional enrichment of PLEs-DMRs in Path B.* 30

**7. Cannabis interaction analyses** **31**

*7.1. Model diagnostics* *31*
**Figure S8.** *QQ plots for cannabis interaction analyses.* 31

*7.2. EWAS interaction results* *32*
**Table S8.** *Top CpG sites showing suggestive evidence of interaction (cumulative childhood trauma)* 32
**Table S9.** *Top CpG sites showing suggestive evidence of interaction (early childhood trauma)* 34
**Table S10.** *Top CpG sites showing suggestive evidence of interaction (late childhood trauma)* 37
**Table S11.** *Top CpG sites showing suggestive evidence of interaction (abuse-only trauma)* 41
**Figure S9.** *Epigenome-wide interaction analysis of childhood trauma and cannabis.* 44

*7.3. DMR interaction results* *45*
**Table S12.** *Differentially methylated regions identified in trauma x cannabis interaction analyses* 45

*7.4. Functional enrichment* *48*
**Table S13.** *Gene Ontology (GO) and KEGG pathway enrichment results for genes annotated to DMRs* 48
**Figure S10.** *Functional enrichment of cannabis-moderated trauma-DMRs.* 50

**8. Mediation Analyses (DACT)** **51**

*8.1. DACT Model Diagnostics* *51*

**Figure S11.** *QQ plots for EWAS analyses using the DACT framework.* 51

*8.2. CpG-level mediation results* *52*

**Figure S12.** *Epigenome-wide mediation analysis of early childhood trauma and PLEs (DACT).* 52

**Figure S13.** *Quadrant plots of CpG-level associations for Path A and Path B.* 53

*8.3. Cannabis-moderated Mediation (DACT)* *54*

**Figure S14.** *QQ plots for EWAS-moderated analyses using the DACT framework.* 54

**Figure S15.** *Quadrant plots of CpG-level associations for Cannabis-moderated Path A and Path B.* 55

**Table S14.** *Top CpG sites showing suggestive evidence of mediation* 56

### *1. Additional Methods*

#### *1.1. DNAm preprocessing and quality control*

Quality control (QC) procedures were systematically applied to raw DNA methylation data generated using the Illumina HumanMethylation450 BeadChip (450k). QC procedures were applied at both the sample and probe level. Samples were excluded based on sex mismatch, poor signal intensity, and low overall quality (i.e., > 10% of probes had a detection p-value > 0.01 or a bead count < 3). Probes were excluded if they had detection p-value > 0.01 in > 10% of samples, bead count < 3 in > 10% of sample, or overlapped known single nucleotide polymorphisms. The present study was restricted to participant with DNA methylation measured at age 15-17 years.

#### *1.2. DNAm Principal Component Analysis (mPCs)*

To capture latent sources of biological and technical variation in the DNAm data, principal component analysis (PCA) was performed on genome-wide methylation levels measured at age 15-17 years. PCA was conducted on a random subset of 50,000 CpG sites selected from the genome-wide dataset. Methylation $\beta$-values were transformed to M-values using the logit transformation. CpGs with more than 1% missingness were excluded and remaining missing values (< 1% of the data matrix) were imputed using the mean methylation level for each CpG. CpGs with zero variance were removed prior to analysis. PCA was performed using the *prcomp* function in R, with centring and scaling applied across CpGs. The first five components (mPC1-mPC5) were retained as covariates in all models.

#### *1.3. Smoking score derivation and validation*

To account for tobacco exposure in epigenetic analyses, we derived a DNA methylation-based smoking indicator using methylation levels at cg05575921 within the aryl-hydrocarbon receptor repressor (AHRR) gene. This CpG site is one of the most robust and consistently replicated methylation markers of cigarette smoking identified in large EWAS studies.

To evaluate whether cg05575921 methylation behaved as expected in this cohort, we examined its association with self-reported nicotine dependence assessed at age 17 using the Fagerström Test for Nicotine Dependence. Valid questionnaire data were available for a subsample of 286 participants. Higher nicotine dependence was associated with lower methylation at cg05575921 ($\beta$ = -0.0099 per unit increase in Fagerström score, SE = 0.0020, p = 1.15 x 10-6; R2 = 0.08). Spearman's rank correlation confirmed a moderate inverse association ($\rho$ = -0.27, p = 2.7 x 10-6; **Figure S1**). Consistent with this, participants with Fagerström scores greater than zero showed lower mean methylation compared to those scoring zero (mean difference ≈ -0.041, p < 0.001).

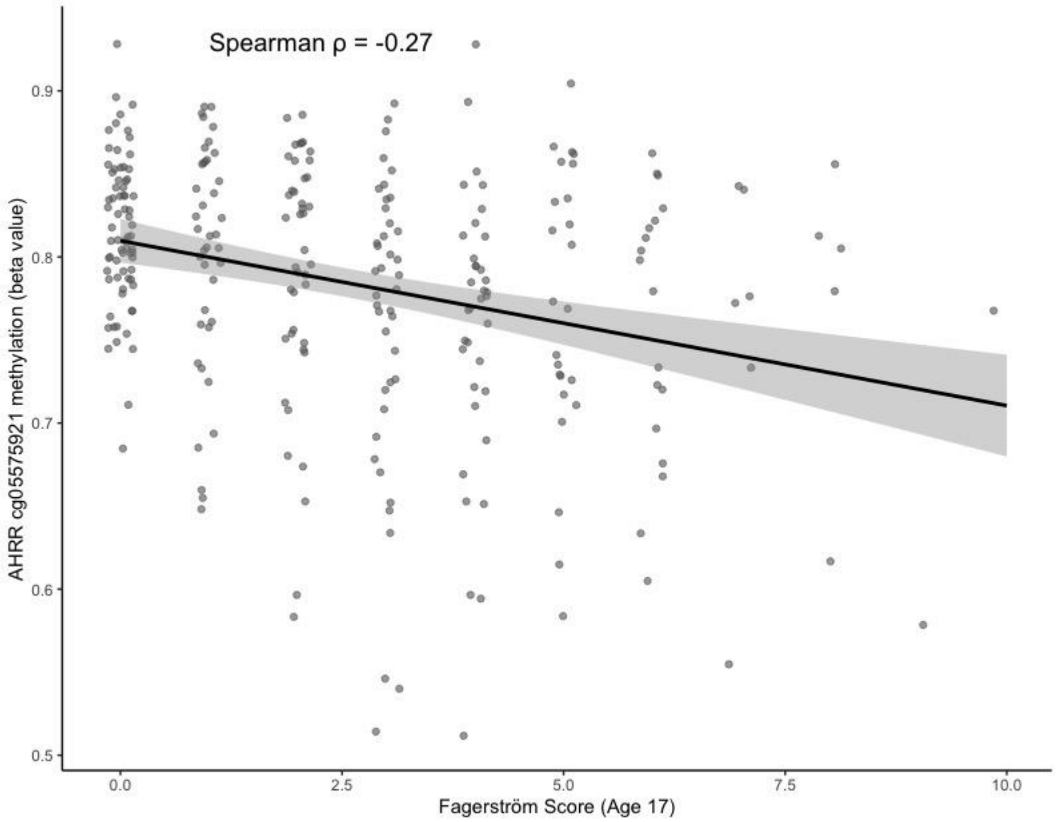


**Figure S1.** *Association between AHRR methylation and nicotine dependence at age 17.*
Scatter plot of cg05575921 (AHRR) methylation $\beta$ values against Fagerström Test for Nicotine Dependence scores at age 17 among participants with available data (N = 286). The solid line represents the fitted linear regression with 95% CI.

These findings support the use of cg05575921 methylation as a DNAm-based indicator of smoking exposure in subsequent analyses.

##### *1.4. Cannabis frequency imputation*

Cannabis use frequency at age 15.5 years was derived as a three-level ordinal variable (0 = never, 1 = occasional, 2 = frequent) based on self-reported lifetime use and frequency measures. Occasional use corresponded to lower-frequency categories, whereas frequent use reflected higher-frequency categories among users. Missing values in cannabis frequency were imputed using multiple imputation by chained equations (MICE). The imputation model used an ordinal logistic regression approach and included earlier cannabis-related variables, sex, smoking score, trauma exposure, and psychotic-like experiences (PLEs) as predictors. A total of 20 imputed datasets were generated with 20 iterations per chain. Imputed values were extracted from the completed datasets and used in downstream analyses.

#### *2. Supplementary Results: Phenotypic Associations*

**Table S1.** *Phenotypic associations with psychotic-like experiences at age 18*

| Exposure / term | Outcome | N | OR | 95% CI | P-value |
|---|---|---|---|---|---|
| *Cumulative trauma score (0–17 years)* | PLEs at age 18 | 1457 | 1.59 | 1.38 to 1.84 | $2.73\times10^{-10}$ |
| *Occasional cannabis use vs never* | PLEs at age 18 | 1457 | 1.14 | 0.68 to 1.91 | 0.6061 |
| *Frequent cannabis use vs never* | PLEs at age 18 | 1457 | 2.94 | 1.21 to 7.14 | 0.0168 |
| *Trauma × occasional cannabis use* | PLEs at age 18 | 1457 | 1.04 | 0.71 to 1.51 | 0.8396 |
| *Trauma × frequent cannabis use* | PLEs at age 18 | 1457 | 1.50 | 0.63 to 3.59 | 0.3626 |

Models adjusted for sex, age, and maternal education.

#### 3. EWAS diagnostics

##### 3.1. Path A Diagnostics

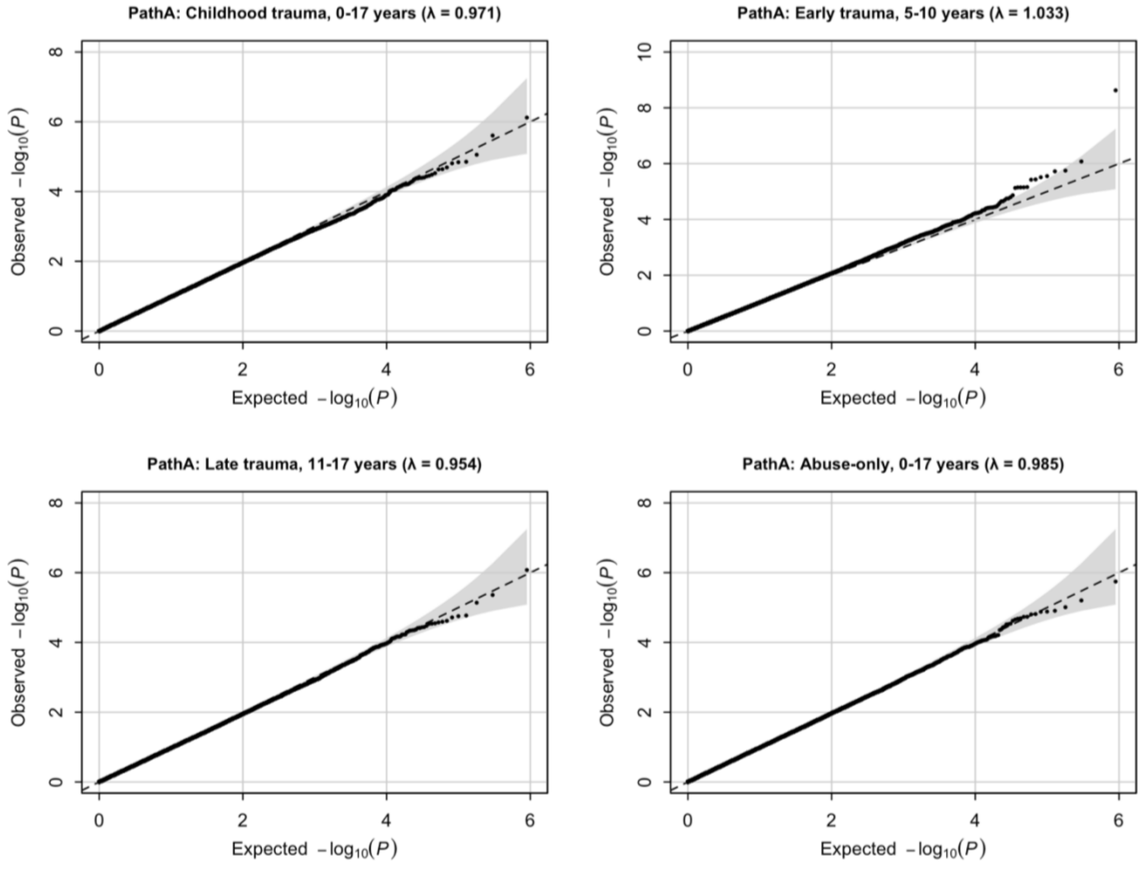


**Figure S2.** *QQ plots for EWAS analyses of childhood trauma and DNAm (Path A).*
The dashed line indicates the null expectation and the shaded region the 95% confidence envelope. Genomic inflation factors (λ) are reported in each panel.

*3.2. Path B Diagnostics*

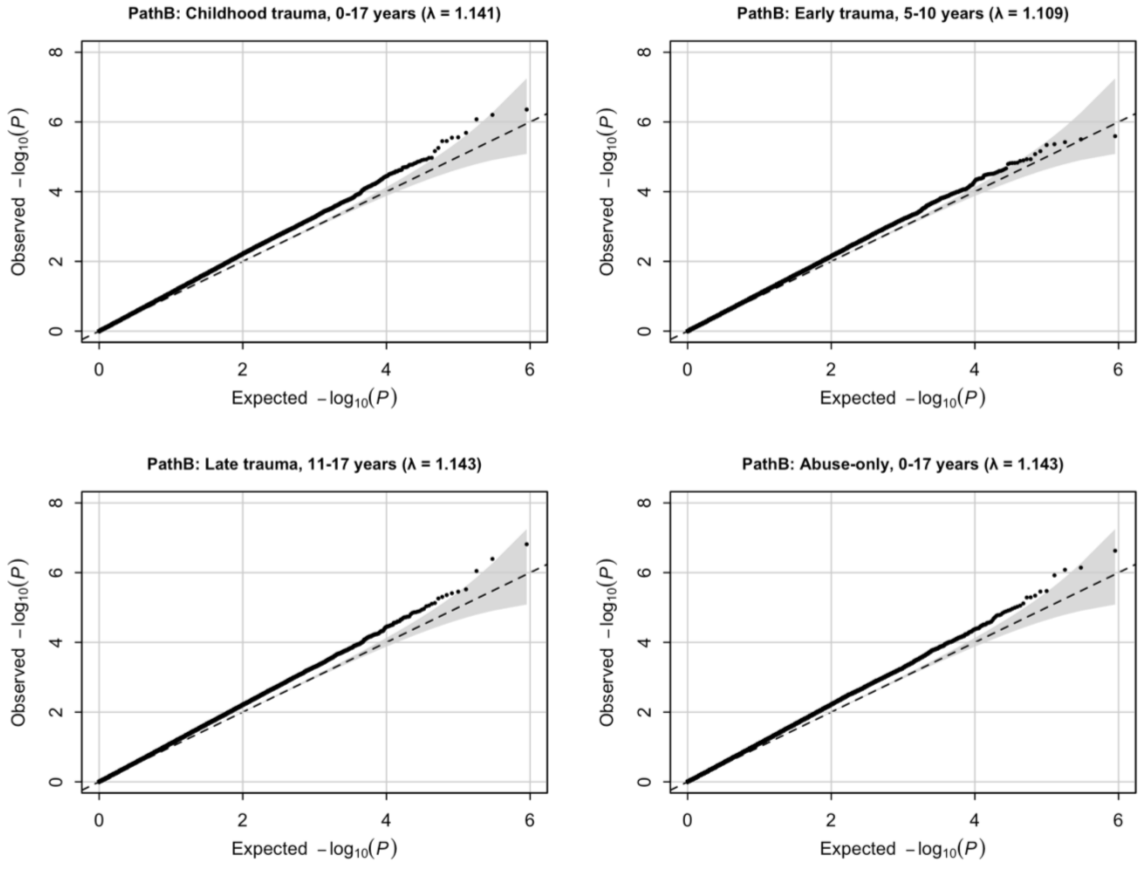


**Figure S3.** *QQ plots for EWAS analyses of DNAm and PLEs (Path B).*
The dashed line indicates the null expectation and the shaded region the 95% confidence envelope. Genomic inflation factors (λ) are reported in each panel.

#### *4. Epigenome-wide Association Results*

EWAS results are presented separately for Path A and Path B across the four childhood trauma definitions. For each analysis, top CpG associations are reported in supplementary tables.

##### *4.1. Path A – Associations between childhood trauma and DNAm*

**Table S2.** *Top CpG sites associated with childhood trauma in EWAS association analyses*

| CpG | chr | pos | gene | cpg_island_relation | effect | SE | P | exposure |
|---|---|---|---|---|---|---|---|---|
| *cg08423269* | chr3 | 157827682 | RSRC1 | N_Shore | 0.0254 | 0.0051 | 7.598E-07 | Childhood trauma, 0-17 years |
| *cg00284390* | chr16 | 84492635 | ATP2C2 | OpenSea | -0.0451 | 0.0095 | 2.482E-06 | Childhood trauma, 0-17 years |
| *cg03667641* | chr7 | 1046346 | C7orf50 | Island | 0.0195 | 0.0044 | 8.808E-06 | Childhood trauma, 0-17 years |
| *cg00816008* | chr16 | 82203996 | MPHOSPH6 | Island | -0.0242 | 0.0056 | 1.400E-05 | Childhood trauma, 0-17 years |
| *cg12442369* | chr19 | 12167216 | ZNF878 | S_Shelf | 0.0230 | 0.0053 | 1.441E-05 | Childhood trauma, 0-17 years |
| *cg16853982* | chr1 | 236849389 | ACTN2 | N_Shore | -0.0344 | 0.0079 | 1.553E-05 | Childhood trauma, 0-17 years |
| *cg11527220* | chr17 | 49041451 | SPAG9 | OpenSea | -0.0224 | 0.0052 | 2.037E-05 | Childhood trauma, 0-17 years |
| *cg09392615* | chr15 | 33023237 | GREM1 | OpenSea | 0.0253 | 0.0060 | 2.284E-05 | Childhood trauma, 0-17 years |
| *cg02770794* | chrX | 39872204 | | Island | -0.0485 | 0.0114 | 2.320E-05 | Childhood trauma, 0-17 years |
| *cg24317979* | chr2 | 87302855 | LOC285074 | Island | 0.0180 | 0.0043 | 2.975E-05 | Childhood trauma, 0-17 years |
| *cg27235829* | chr1 | 158390759 | OR10K2 | OpenSea | -0.0227 | 0.0054 | 3.271E-05 | Childhood trauma, 0-17 years |
| *cg16550477* | chrX | 8769499 | FAM9A | Island | 0.0782 | 0.0188 | 3.513E-05 | Childhood trauma, 0-17 years |
| *cg16906595* | chr11 | 67798128 | NDUFS8 | Island | 0.0247 | 0.0060 | 3.659E-05 | Childhood trauma, 0-17 years |
| *cg21982660* | chr20 | 17922310 | SNX5 | OpenSea | -0.0245 | 0.0059 | 3.954E-05 | Childhood trauma, 0-17 years |
| *cg00907272* | chr10 | 74086782 | | OpenSea | -0.0496 | 0.0120 | 4.047E-05 | Childhood trauma, 0-17 years |
| *cg08158862* | chr1 | 200012526 | NR5A2 | S_Shore | -0.0442 | 0.0107 | 4.058E-05 | Childhood trauma, 0-17 years |
| *cg26583088* | chr17 | 40322250 | KCNH4 | OpenSea | -0.0595 | 0.0145 | 4.192E-05 | Childhood trauma, 0-17 years |
| *cg22025233* | chr7 | 1022904 | CYP2W1 | N_Shore | 0.0281 | 0.0068 | 4.201E-05 | Childhood trauma, 0-17 years |
| *cg24402667* | chr17 | 36612623 | ARHGAP23 | S_Shelf | 0.0383 | 0.0094 | 4.561E-05 | Childhood trauma, 0-17 years |

| | | | | | | | | |
|---|---|---|---|---|---|---|---|---|
| *cg16255307* | chr3 | 185271383 | LIPH | OpenSea | -0.0609 | 0.0101 | 2.363E-09 | Early trauma, 5-10 years |
| *cg15057061* | chr3 | 181437299 | SOX2OT | Island | 0.0544 | 0.0110 | 8.348E-07 | Early trauma, 5-10 years |
| *cg13401681* | chrX | 16141609 | GRPR | OpenSea | 0.0390 | 0.0081 | 1.780E-06 | Early trauma, 5-10 years |
| *cg04317047* | chr5 | 148808388 | MIR143; LOC728264 | OpenSea | -0.0751 | 0.0157 | 1.889E-06 | Early trauma, 5-10 years |
| *cg06386482* | chr6 | 47624117 | GPR111 | OpenSea | 0.1664 | 0.0354 | 2.802E-06 | Early trauma, 5-10 years |
| *cg02552572* | chr16 | 56597746 | MT4 | OpenSea | -0.0490 | 0.0105 | 3.081E-06 | Early trauma, 5-10 years |
| *cg11527220* | chr17 | 49041451 | SPAG9 | OpenSea | -0.0346 | 0.0074 | 3.679E-06 | Early trauma, 5-10 years |
| *cg05491661* | chr12 | 56893671 | | OpenSea | 0.0469 | 0.0101 | 3.767E-06 | Early trauma, 5-10 years |
| *cg10523820* | chr11 | 66314211 | ACTN3; ZDHHC24 | Island | -0.0893 | 0.0198 | 6.938E-06 | Early trauma, 5-10 years |
| *cg02908285* | chr3 | 30951484 | | OpenSea | 0.0367 | 0.0081 | 7.094E-06 | Early trauma, 5-10 years |
| *cg09159389* | chr1 | 146644318 | PRKAB2 | Island | -0.0282 | 0.0063 | 7.124E-06 | Early trauma, 5-10 years |
| *cg25607225* | chr2 | 88803870 | | OpenSea | -0.0569 | 0.0126 | 7.138E-06 | Early trauma, 5-10 years |
| *cg25110782* | chr12 | 57624371 | SHMT2 | S_Shore | -0.0253 | 0.0056 | 7.437E-06 | Early trauma, 5-10 years |
| *cg10486655* | chr16 | 68624584 | | OpenSea | 0.0522 | 0.0120 | 1.364E-05 | Early trauma, 5-10 years |
| *cg22504094* | chr19 | 2115196 | AP3D1 | N_Shore | -0.0563 | 0.0130 | 1.590E-05 | Early trauma, 5-10 years |
| *cg22155281* | chr4 | 118007206 | TRAM1L1 | S_Shore | 0.0430 | 0.0100 | 1.764E-05 | Early trauma, 5-10 years |
| *cg13220569* | chr12 | 5155343 | KCNA5 | S_Shore | -0.0407 | 0.0094 | 1.787E-05 | Early trauma, 5-10 years |
| *cg08252971* | chr4 | 41218767 | | Island | 0.0395 | 0.0092 | 2.010E-05 | Early trauma, 5-10 years |
| *cg14698154* | chr5 | 96747223 | | OpenSea | -0.0400 | 0.0094 | 2.234E-05 | Early trauma, 5-10 years |
| *cg18331412* | chr5 | 135364986 | TGFBI | Island | 0.0480 | 0.0113 | 2.243E-05 | Early trauma, 5-10 years |
| *cg05823589* | chr5 | 863749 | | N_Shore | 0.0615 | 0.0146 | 2.562E-05 | Early trauma, 5-10 years |
| *cg27588515* | chr5 | 115167455 | ATG12 | OpenSea | 0.0247 | 0.0059 | 2.965E-05 | Early trauma, 5-10 years |
| *cg09019916* | chr2 | 109335672 | RANBP2 | N_Shore | -0.0572 | 0.0137 | 3.338E-05 | Early trauma, 5-10 years |
| *cg24282422* | chr3 | 125239067 | SNX4 | Island | -0.0236 | 0.0057 | 3.519E-05 | Early trauma, 5-10 years |
| *cg03743462* | chr2 | 240118958 | HDAC4 | OpenSea | -0.0758 | 0.0183 | 3.585E-05 | Early trauma, 5-10 years |
| *cg00284390* | chr16 | 84492635 | ATP2C2 | OpenSea | -0.0561 | 0.0136 | 3.700E-05 | Early trauma, 5-10 years |
| *cg05893029* | chr8 | 88760082 | | OpenSea | -0.0351 | 0.0085 | 3.738E-05 | Early trauma, 5-10 years |
| *cg03070194* | chr1 | 110210684 | GSTM2 | Island | -0.0408 | 0.0099 | 3.749E-05 | Early trauma, 5-10 years |
| *cg23906067* | chr6 | 31939112 | STK19; DOM3Z | N_Shore | -0.0337 | 0.0081 | 3.794E-05 | Early trauma, 5-10 years |

| | | | | | | | | |
|---|---|---|---|---|---|---|---|---|
| *cg10345936* | chr5 | 150727812 | SLC36A2 | OpenSea | -0.0384 | 0.0093 | 3.836E-05 | Early trauma, 5-10 years |
| *cg08584868* | chr8 | 53474887 | FAM150A | N_Shelf | -0.0479 | 0.0116 | 3.925E-05 | Early trauma, 5-10 years |
| *ch.2.4116732R* | chr2 | 206658319 | NRP2 | OpenSea | 0.0344 | 0.0083 | 4.016E-05 | Early trauma, 5-10 years |
| *cg02039022* | chr5 | 72112005 | TNPO1 | N_Shore | -0.0586 | 0.0143 | 4.201E-05 | Early trauma, 5-10 years |
| *cg00259584* | chr2 | 106986308 | | OpenSea | -0.0419 | 0.0103 | 4.582E-05 | Early trauma, 5-10 years |
| *cg22475576* | chr19 | 1252813 | MIDN | N_Shore | -0.0364 | 0.0089 | 4.616E-05 | Early trauma, 5-10 years |
| *cg17098965* | chr20 | 52199520 | ZNF217 | S_Shore | -0.0405 | 0.0099 | 4.691E-05 | Early trauma, 5-10 years |
| *cg07708806* | chr16 | 8962224 | CARHSP1 | Island | -0.0227 | 0.0056 | 4.963E-05 | Early trauma, 5-10 years |
| *cg04795788* | chr13 | 101327638 | TMTC4 | Island | -0.0379 | 0.0077 | 8.385E-07 | Late trauma, 11-17 years |
| *cg08816194* | chrY | 16733442 | NLGN4Y | OpenSea | -0.0963 | 0.0208 | 4.355E-06 | Late trauma, 11-17 years |
| *cg16902255* | chr1 | 35897100 | | OpenSea | 0.0686 | 0.0152 | 7.254E-06 | Late trauma, 11-17 years |
| *cg16906137* | chr15 | 72075886 | | OpenSea | -0.0495 | 0.0115 | 1.694E-05 | Late trauma, 11-17 years |
| *cg17240157* | chr1 | 164816968 | PBX1 | OpenSea | 0.0590 | 0.0137 | 1.767E-05 | Late trauma, 11-17 years |
| *cg17289597* | chr10 | 72433155 | ADAMTS14 | S_Shore | 0.0674 | 0.0157 | 1.922E-05 | Late trauma, 11-17 years |
| *cg27413385* | chr17 | 7515425 | FXR2 | N_Shelf | -0.0386 | 0.0091 | 2.403E-05 | Late trauma, 11-17 years |
| *cg14406727* | chr2 | 65577769 | SPRED2 | OpenSea | 0.0464 | 0.0110 | 2.548E-05 | Late trauma, 11-17 years |
| *cg10186400* | chr6 | 46138843 | ENPP5 | OpenSea | 0.0455 | 0.0108 | 2.654E-05 | Late trauma, 11-17 years |
| *cg16949378* | chr17 | 2276201 | SGSM2 | S_Shore | -0.0591 | 0.0141 | 2.834E-05 | Late trauma, 11-17 years |
| *cg10713773* | chr13 | 51796714 | FAM124A | Island | 0.0408 | 0.0097 | 2.887E-05 | Late trauma, 11-17 years |
| *cg24297509* | chr10 | 44339107 | | OpenSea | -0.0714 | 0.0170 | 2.941E-05 | Late trauma, 11-17 years |
| *cg08819934* | chr8 | 146082699 | | S_Shelf | 0.0482 | 0.0116 | 3.356E-05 | Late trauma, 11-17 years |
| *cg02078882* | chr12 | 30911709 | | S_Shelf | -0.0804 | 0.0194 | 3.614E-05 | Late trauma, 11-17 years |
| *cg16035633* | chr7 | 150700728 | NOS3 | OpenSea | -0.0611 | 0.0147 | 3.686E-05 | Late trauma, 11-17 years |
| *cg21815882* | chr7 | 26241064 | CBX3; HNRNPA2B1 | Island | -0.0355 | 0.0086 | 3.868E-05 | Late trauma, 11-17 years |
| *cg24602560* | chr11 | 788583 | CEND1 | N_Shore | 0.0929 | 0.0225 | 3.875E-05 | Late trauma, 11-17 years |
| *cg13298632* | chr2 | 242952648 | | Island | 0.0772 | 0.0188 | 4.257E-05 | Late trauma, 11-17 years |
| *cg10721834* | chr19 | 22990066 | | Island | 0.0816 | 0.0199 | 4.445E-05 | Late trauma, 11-17 years |
| *cg24153199* | chr14 | 50698328 | SOS2 | Island | -0.0317 | 0.0077 | 4.478E-05 | Late trauma, 11-17 years |
| *cg02426836* | chr11 | 2444651 | TRPM5 | S_Shelf | 0.0549 | 0.0134 | 4.553E-05 | Late trauma, 11-17 years |

| | | | | | | | | |
|---|---|---|---|---|---|---|---|---|
| *cg00190795* | chr3 | 195599898 | TNK2 | N_Shore | -0.0405 | 0.0099 | 4.607E-05 | Late trauma, 11-17 years |
| *cg01838916* | chr5 | 85577056 | NBPF22P | OpenSea | -0.0416 | 0.0102 | 4.906E-05 | Late trauma, 11-17 years |
| *cg08423269* | chr3 | 157827682 | RSRC1 | N_Shore | 0.0375 | 0.0078 | 1.795E-06 | Abuse only, 0-17 years |
| *cg23493020* | chr14 | 65112169 | | OpenSea | 0.0390 | 0.0086 | 6.259E-06 | Abuse only, 0-17 years |
| *cg15349139* | chr11 | 1293211 | | Island | -0.0558 | 0.0126 | 9.771E-06 | Abuse only, 0-17 years |
| *cg03699765* | chr10 | 43882723 | HNRNPF | OpenSea | -0.0605 | 0.0138 | 1.236E-05 | Abuse only, 0-17 years |
| *cg21982660* | chr20 | 17922310 | SNX5 | OpenSea | -0.0396 | 0.0091 | 1.305E-05 | Abuse only, 0-17 years |
| *cg27413385* | chr17 | 7515425 | FXR2 | N_Shelf | -0.0360 | 0.0082 | 1.324E-05 | Abuse only, 0-17 years |
| *cg08292485* | chr20 | 29993663 | DEFB121 | OpenSea | -0.0562 | 0.0130 | 1.535E-05 | Abuse only, 0-17 years |
| *cg00578354* | chr20 | 47251399 | PREX1 | N_Shore | -0.0556 | 0.0128 | 1.574E-05 | Abuse only, 0-17 years |
| *cg01641174* | chr15 | 91497412 | RCCD1 | N_Shore | -0.0273 | 0.0064 | 1.838E-05 | Abuse only, 0-17 years |
| *cg00400263* | chr20 | 58514201 | C20orf177; PPP1R3D | Island | -0.1140 | 0.0265 | 1.850E-05 | Abuse only, 0-17 years |
| *cg11527220* | chr17 | 49041451 | SPAG9 | OpenSea | -0.0343 | 0.0080 | 2.030E-05 | Abuse only, 0-17 years |
| *cg00816008* | chr16 | 82203996 | MPHOSPH6 | Island | -0.0362 | 0.0085 | 2.091E-05 | Abuse only, 0-17 years |
| *cg14564144* | chr12 | 650711 | B4GALNT3 | OpenSea | -0.0578 | 0.0136 | 2.215E-05 | Abuse only, 0-17 years |
| *cg14948030* | chr6 | 124122575 | | N_Shore | -0.0457 | 0.0108 | 2.379E-05 | Abuse only, 0-17 years |
| *cg02426623* | chr12 | 3259078 | TSPAN9 | OpenSea | 0.0579 | 0.0138 | 2.896E-05 | Abuse only, 0-17 years |
| *cg02994237* | chrX | 137750892 | FGF13; MIR504 | OpenSea | -0.0758 | 0.0181 | 2.993E-05 | Abuse only, 0-17 years |
| *cg21500735* | chr19 | 44576159 | ZNF284 | Island | -0.0398 | 0.0095 | 3.028E-05 | Abuse only, 0-17 years |
| *cg00743803* | chr22 | 48970600 | FAM19A5 | N_Shore | 0.0468 | 0.0113 | 3.514E-05 | Abuse only, 0-17 years |
| *cg04693701* | chr5 | 125934946 | | N_Shore | 0.0362 | 0.0087 | 3.716E-05 | Abuse only, 0-17 years |
| *cg13962846* | chr1 | 7973298 | | OpenSea | -0.0840 | 0.0204 | 4.204E-05 | Abuse only, 0-17 years |
| *cg00473749* | chr7 | 63955932 | | OpenSea | 0.0602 | 0.0147 | 4.409E-05 | Abuse only, 0-17 years |

Genomic locations are reported according to the hg19 reference genome. Gene annotation and CpG island context correspond to the Illumina HumanMethylation450 array annotation.

*4.2 Path B – Associations between DNAm and PLEs*

**Table S3.** *Top CpG sites associated with PLEs in EWAS association analyses (adjusted for childhood trauma)*

| CpG | chr | pos | gene | cpg_island_relation | effect | SE | P | exposure |
|---|---|---|---|---|---|---|---|---|
| *cg26428054* | chr12 | 49484058 | DHH | Island | -3.238 | 0.641 | 4.4E-07 | Childhood trauma, 0-17 years |
| *cg10529789* | chr17 | 37009658 | SNORA21; RPL23 | Island | -2.76 | 0.5539 | 6.25E-07 | Childhood trauma, 0-17 years |
| *cg24157392* | chr3 | 112217973 | BTLA | OpenSea | 2.298 | 0.4664 | 8.38E-07 | Childhood trauma, 0-17 years |
| *cg07992052* | chr20 | 35402295 | DSN1 | Island | -1.949 | 0.4103 | 2.05E-06 | Childhood trauma, 0-17 years |
| *cg08528970* | chr4 | 76640579 | | OpenSea | -1.523 | 0.3248 | 2.76E-06 | Childhood trauma, 0-17 years |
| *cg14663914* | chr19 | 827739 | AZU1 | OpenSea | -2.193 | 0.4683 | 2.83E-06 | Childhood trauma, 0-17 years |
| *cg11182518* | chr5 | 55117965 | | Island | 0.8451 | 0.1822 | 3.53E-06 | Childhood trauma, 0-17 years |
| *cg01312828* | chr6 | 41648365 | | N_Shelf | -1.266 | 0.2731 | 3.55E-06 | Childhood trauma, 0-17 years |
| *cg14672994* | chr17 | 48503057 | ACSF2 | Island | -1.541 | 0.3391 | 5.56E-06 | Childhood trauma, 0-17 years |
| *cg21041594* | chr7 | 65235699 | | Island | 1.717 | 0.3817 | 6.86E-06 | Childhood trauma, 0-17 years |
| *cg00980622* | chr14 | 75884845 | | OpenSea | -1.32 | 0.2998 | 1.06E-05 | Childhood trauma, 0-17 years |
| *cg25618765* | chr22 | 20670922 | | N_Shelf | -1.508 | 0.3426 | 1.08E-05 | Childhood trauma, 0-17 years |
| *cg21889703* | chr6 | 136607649 | BCLAF1 | N_Shelf | -2.093 | 0.4779 | 1.19E-05 | Childhood trauma, 0-17 years |
| *cg14010194* | chr6 | 42152817 | GUCA1B | OpenSea | -1.251 | 0.2858 | 1.21E-05 | Childhood trauma, 0-17 years |
| *cg00464520* | chr10 | 118903060 | | S_Shelf | -1.151 | 0.2635 | 1.26E-05 | Childhood trauma, 0-17 years |
| *cg07942847* | chr14 | 23420757 | HAUS4 | OpenSea | 2.374 | 0.5455 | 1.35E-05 | Childhood trauma, 0-17 years |
| *cg10755512* | chr3 | 44666543 | ZNF197 | Island | 2.091 | 0.4813 | 1.39E-05 | Childhood trauma, 0-17 years |
| *cg08362273* | chr17 | 46719577 | | Island | 1.326 | 0.3064 | 0.000015 | Childhood trauma, 0-17 years |
| *cg08201736* | chr15 | 79298769 | RASGRF1 | OpenSea | 1.582 | 0.3662 | 1.56E-05 | Childhood trauma, 0-17 years |
| *cg05640294* | chr19 | 38976794 | RYR1 | Island | -1.539 | 0.3575 | 1.67E-05 | Childhood trauma, 0-17 years |
| *cg16791444* | chr17 | 71702669 | | OpenSea | -1.346 | 0.3128 | 1.69E-05 | Childhood trauma, 0-17 years |
| *cg14841628* | chr1 | 16258280 | SPEN | N_Shelf | -1.476 | 0.3432 | 1.71E-05 | Childhood trauma, 0-17 years |
| *cg16328610* | chr6 | 83777335 | DOPEY1 | Island | -2.136 | 0.499 | 1.87E-05 | Childhood trauma, 0-17 years |
| *cg17276036* | chr4 | 26492222 | CCKAR | OpenSea | 1.792 | 0.4198 | 1.97E-05 | Childhood trauma, 0-17 years |
| *cg19013753* | chr15 | 75915192 | SNUPN | N_Shelf | 1.536 | 0.36 | 1.98E-05 | Childhood trauma, 0-17 years |

| | | | | | | | | |
|---|---|---|---|---|---|---|---|---|
| *cg08739188* | chr5 | 75377860 | SV2C | N_Shore | 0.987 | 0.2314 | 2E-05 | Childhood trauma, 0-17 years |
| *cg03315940* | chr6 | 27569167 | | OpenSea | -1.544 | 0.3624 | 2.03E-05 | Childhood trauma, 0-17 years |
| *cg05771369* | chr12 | 58021713 | B4GALNT1 | Island | 0.6714 | 0.1588 | 2.36E-05 | Childhood trauma, 0-17 years |
| *cg19754901* | chr3 | 87102643 | | S_Shore | -1.408 | 0.3333 | 2.41E-05 | Childhood trauma, 0-17 years |
| *cg27524192* | chr1 | 228773464 | | Island | -1.482 | 0.351 | 2.42E-05 | Childhood trauma, 0-17 years |
| *cg05814106* | chr1 | 2224791 | SKI | S_Shelf | -1.018 | 0.2414 | 2.47E-05 | Childhood trauma, 0-17 years |
| *cg10632722* | chr6 | 49833873 | CRISP1 | OpenSea | -1.69 | 0.4017 | 2.6E-05 | Childhood trauma, 0-17 years |
| *cg19989295* | chr14 | 24641077 | REC8 | Island | 0.9323 | 0.2217 | 2.61E-05 | Childhood trauma, 0-17 years |
| *cg16588073* | chr1 | 156647247 | NES | Island | -1.139 | 0.2708 | 2.62E-05 | Childhood trauma, 0-17 years |
| *cg01996004* | chr5 | 180338435 | BTNL8 | OpenSea | -1.021 | 0.2439 | 2.83E-05 | Childhood trauma, 0-17 years |
| *cg22936975* | chr13 | 61989412 | PCDH20 | OpenSea | 1.668 | 0.3984 | 2.83E-05 | Childhood trauma, 0-17 years |
| *cg00925244* | chr2 | 86148800 | | OpenSea | 2.066 | 0.494 | 2.89E-05 | Childhood trauma, 0-17 years |
| *cg10251328* | chr17 | 47074713 | IGF2BP1 | Island | -2.451 | 0.587 | 2.97E-05 | Childhood trauma, 0-17 years |
| *cg08791347* | chr10 | 13831250 | FRMD4A | OpenSea | 1.085 | 0.2602 | 3.05E-05 | Childhood trauma, 0-17 years |
| *cg23041896* | chr8 | 144462235 | RHPN1 | Island | -0.7632 | 0.1831 | 3.08E-05 | Childhood trauma, 0-17 years |
| *cg05259836* | chr6 | 74290516 | | OpenSea | -1.379 | 0.3314 | 3.14E-05 | Childhood trauma, 0-17 years |
| *cg02324227* | chr5 | 180591594 | | OpenSea | 1.71 | 0.4112 | 3.2E-05 | Childhood trauma, 0-17 years |
| *cg09043104* | chr8 | 117128288 | | OpenSea | -1.988 | 0.4794 | 3.39E-05 | Childhood trauma, 0-17 years |
| *cg18343437* | chr8 | 142528415 | | Island | 0.6246 | 0.1511 | 3.55E-05 | Childhood trauma, 0-17 years |
| *cg23242341* | chr11 | 66113666 | BRMS1; B3GNT1 | N_Shore | -1.047 | 0.2532 | 3.57E-05 | Childhood trauma, 0-17 years |
| *cg19414967* | chr8 | 49468296 | | N_Shore | 1.487 | 0.3598 | 3.58E-05 | Childhood trauma, 0-17 years |
| *cg23018689* | chr1 | 159173540 | DARC | OpenSea | -1.728 | 0.4191 | 3.76E-05 | Childhood trauma, 0-17 years |
| *cg03748603* | chr19 | 2494443 | | Island | -0.7823 | 0.1899 | 3.79E-05 | Childhood trauma, 0-17 years |
| *cg13561817* | chr17 | 3213102 | OR3A4 | OpenSea | -0.9209 | 0.2239 | 3.91E-05 | Childhood trauma, 0-17 years |
| *cg19052355* | chr2 | 237076306 | GBX2 | Island | 1.492 | 0.3633 | 3.99E-05 | Childhood trauma, 0-17 years |
| *cg03006175* | chr14 | 105412417 | AHNAK2 | OpenSea | -1.434 | 0.35 | 0.000042 | Childhood trauma, 0-17 years |
| *cg04177395* | chr11 | 78496497 | ODZ4 | OpenSea | 0.7618 | 0.1864 | 4.37E-05 | Childhood trauma, 0-17 years |
| *cg09361493* | chr2 | 231715028 | | S_Shelf | -1.364 | 0.334 | 4.43E-05 | Childhood trauma, 0-17 years |
| *cg19024989* | chr11 | 2024126 | | N_Shelf | -2.009 | 0.4921 | 4.46E-05 | Childhood trauma, 0-17 years |

| | | | | | | | | |
|---|---|---|---|---|---|---|---|---|
| *cg17473656* | chr4 | 190286944 | | S_Shore | -1.929 | 0.4741 | 4.73E-05 | Childhood trauma, 0-17 years |
| *cg18928900* | chr6 | 170605493 | | Island | 1.796 | 0.4417 | 4.77E-05 | Childhood trauma, 0-17 years |
| *cg25644556* | chr7 | 27209582 | MIR196B | Island | 0.7151 | 0.1762 | 4.96E-05 | Childhood trauma, 0-17 years |
| *cg10529789* | chr17 | 37009658 | SNORA21; RPL23 | Island | -2.584 | 0.5494 | 2.56E-06 | Early trauma, 5-10 years |
| *cg14672994* | chr17 | 48503057 | ACSF2 | Island | -1.62 | 0.3475 | 3.15E-06 | Early trauma, 5-10 years |
| *cg11182518* | chr5 | 55117965 | | Island | 0.8363 | 0.1809 | 3.79E-06 | Early trauma, 5-10 years |
| *cg26428054* | chr12 | 49484058 | DHH | Island | -2.823 | 0.6146 | 4.34E-06 | Early trauma, 5-10 years |
| *cg02324227* | chr5 | 180591594 | | OpenSea | 1.863 | 0.4066 | 4.58E-06 | Early trauma, 5-10 years |
| *cg01312828* | chr6 | 41648365 | | N_Shelf | -1.224 | 0.2722 | 6.91E-06 | Early trauma, 5-10 years |
| *cg25618765* | chr22 | 20670922 | | N_Shelf | -1.486 | 0.3335 | 8.39E-06 | Early trauma, 5-10 years |
| *cg07942847* | chr14 | 23420757 | HAUS4 | OpenSea | 2.3 | 0.5248 | 1.17E-05 | Early trauma, 5-10 years |
| *cg08791347* | chr10 | 13831250 | FRMD4A | OpenSea | 1.13 | 0.2578 | 1.17E-05 | Early trauma, 5-10 years |
| *cg18343437* | chr8 | 142528415 | | Island | 0.6515 | 0.1493 | 1.27E-05 | Early trauma, 5-10 years |
| *cg01996004* | chr5 | 180338435 | BTNL8 | OpenSea | -1.076 | 0.2468 | 1.32E-05 | Early trauma, 5-10 years |
| *cg14663914* | chr19 | 827739 | AZU1 | OpenSea | -1.922 | 0.4436 | 1.47E-05 | Early trauma, 5-10 years |
| *cg27524192* | chr1 | 228773464 | | Island | -1.429 | 0.33 | 1.5E-05 | Early trauma, 5-10 years |
| *cg14010194* | chr6 | 42152817 | GUCA1B | OpenSea | -1.221 | 0.2822 | 1.5E-05 | Early trauma, 5-10 years |
| *cg25644556* | chr7 | 27209582 | MIR196B | Island | 0.7503 | 0.1735 | 1.53E-05 | Early trauma, 5-10 years |
| *cg10755512* | chr3 | 44666543 | ZNF197 | Island | 2.066 | 0.4792 | 1.62E-05 | Early trauma, 5-10 years |
| *cg05814106* | chr1 | 2224791 | SKI | S_Shelf | -1.022 | 0.2406 | 2.15E-05 | Early trauma, 5-10 years |
| *cg18345219* | chr1 | 43919084 | HYI | Island | 1.808 | 0.4268 | 2.29E-05 | Early trauma, 5-10 years |
| *cg21889703* | chr6 | 136607649 | BCLAF1 | N_Shelf | -1.962 | 0.4647 | 2.43E-05 | Early trauma, 5-10 years |
| *cg05640294* | chr19 | 38976794 | RYR1 | Island | -1.487 | 0.3532 | 2.53E-05 | Early trauma, 5-10 years |
| *cg03315940* | chr6 | 27569167 | | OpenSea | -1.517 | 0.3603 | 2.55E-05 | Early trauma, 5-10 years |
| *cg18928900* | chr6 | 170605493 | | Island | 1.857 | 0.4425 | 2.7E-05 | Early trauma, 5-10 years |
| *cg05295015* | chr14 | 38069001 | | Island | 1.85 | 0.4416 | 2.81E-05 | Early trauma, 5-10 years |
| *cg20799816* | chr18 | 21594185 | TTC39C | Island | -1.941 | 0.4643 | 2.92E-05 | Early trauma, 5-10 years |
| *cg25616869* | chr3 | 48443867 | | OpenSea | -2.005 | 0.4807 | 3.03E-05 | Early trauma, 5-10 years |
| *cg19013753* | chr15 | 75915192 | SNUPN | N_Shelf | 1.381 | 0.3315 | 3.09E-05 | Early trauma, 5-10 years |

| | | | | | | | | |
|---|---|---|---|---|---|---|---|---|
| *cg07992052* | chr20 | 35402295 | DSN1 | Island | -1.689 | 0.4053 | 3.1E-05 | Early trauma, 5-10 years |
| *cg24157392* | chr3 | 112217973 | BTLA | OpenSea | 1.853 | 0.4449 | 3.12E-05 | Early trauma, 5-10 years |
| *cg19754901* | chr3 | 87102643 | | S_Shore | -1.356 | 0.3257 | 3.14E-05 | Early trauma, 5-10 years |
| *cg21964551* | chr1 | 201368791 | LAD1 | Island | 0.991 | 0.2382 | 3.17E-05 | Early trauma, 5-10 years |
| *cg25975856* | chr3 | 19930780 | EFHB | OpenSea | -0.4143 | 0.0997 | 3.25E-05 | Early trauma, 5-10 years |
| *cg03769992* | chr10 | 50010141 | WDFY4 | OpenSea | 1.981 | 0.4769 | 3.27E-05 | Early trauma, 5-10 years |
| *cg04177395* | chr11 | 78496497 | ODZ4 | OpenSea | 0.7636 | 0.1841 | 3.35E-05 | Early trauma, 5-10 years |
| *cg05259836* | chr6 | 74290516 | | OpenSea | -1.376 | 0.3321 | 3.41E-05 | Early trauma, 5-10 years |
| *cg08201736* | chr15 | 79298769 | RASGRF1 | OpenSea | 1.454 | 0.3538 | 3.98E-05 | Early trauma, 5-10 years |
| *cg09859179* | chr2 | 58655104 | | OpenSea | 1.644 | 0.4004 | 4.01E-05 | Early trauma, 5-10 years |
| *cg09573795* | chr4 | 4863874 | MSX1 | N_Shore | -0.9278 | 0.2264 | 4.16E-05 | Early trauma, 5-10 years |
| *cg17403875* | chr14 | 55596356 | LGALS3 | Island | -2.333 | 0.5696 | 4.19E-05 | Early trauma, 5-10 years |
| *cg16791444* | chr17 | 71702669 | | OpenSea | -1.263 | 0.3082 | 4.19E-05 | Early trauma, 5-10 years |
| *cg16328610* | chr6 | 83777335 | DOPEY1 | Island | -1.993 | 0.4868 | 4.25E-05 | Early trauma, 5-10 years |
| *cg01581763* | chr7 | 13821699 | | OpenSea | -1.463 | 0.3578 | 4.33E-05 | Early trauma, 5-10 years |
| *cg05533329* | chr7 | 143039088 | CLCN1 | N_Shelf | -1.299 | 0.3181 | 4.42E-05 | Early trauma, 5-10 years |
| *cg23041896* | chr8 | 144462235 | RHPN1 | Island | -0.7421 | 0.1818 | 4.48E-05 | Early trauma, 5-10 years |
| *cg26778516* | chr4 | 8646016 | | Island | -1.039 | 0.2547 | 4.52E-05 | Early trauma, 5-10 years |
| *cg07417857* | chr8 | 652403 | ERICH1 | S_Shore | -0.9627 | 0.2369 | 4.83E-05 | Early trauma, 5-10 years |
| *cg26428054* | chr12 | 49484058 | DHH | Island | -3.425 | 0.6525 | 1.54E-07 | Late trauma, 11-17 years |
| *cg10529789* | chr17 | 37009658 | SNORA21; RPL23 | Island | -2.898 | 0.5719 | 4.05E-07 | Late trauma, 11-17 years |
| *cg11182518* | chr5 | 55117965 | | Island | 0.9253 | 0.1883 | 8.98E-07 | Late trauma, 11-17 years |
| *cg19013753* | chr15 | 75915192 | SNUPN | N_Shelf | 1.756 | 0.376 | 3.01E-06 | Late trauma, 11-17 years |
| *cg19754901* | chr3 | 87102643 | | S_Shore | -1.611 | 0.3475 | 3.53E-06 | Late trauma, 11-17 years |
| *cg07992052* | chr20 | 35402295 | DSN1 | Island | -1.89 | 0.4093 | 3.9E-06 | Late trauma, 11-17 years |
| *cg21889703* | chr6 | 136607649 | BCLAF1 | N_Shelf | -2.281 | 0.4967 | 4.39E-06 | Late trauma, 11-17 years |
| *cg08201736* | chr15 | 79298769 | RASGRF1 | OpenSea | 1.708 | 0.374 | 4.97E-06 | Late trauma, 11-17 years |
| *cg24157392* | chr3 | 112217973 | BTLA | OpenSea | 2.151 | 0.4734 | 5.52E-06 | Late trauma, 11-17 years |
| *cg18848419* | chr18 | 11850153 | CHMP1B; GNAL | N_Shore | 0.8026 | 0.1791 | 7.43E-06 | Late trauma, 11-17 years |

| | | | | | | | | |
|---|---|---|---|---|---|---|---|---|
| *cg05640294* | chr19 | 38976794 | RYR1 | Island | -1.621 | 0.3629 | 7.88E-06 | Late trauma, 11-17 years |
| *cg18928900* | chr6 | 170605493 | | Island | 2.061 | 0.4638 | 8.82E-06 | Late trauma, 11-17 years |
| *cg10755512* | chr3 | 44666543 | ZNF197 | Island | 2.192 | 0.4943 | 9.26E-06 | Late trauma, 11-17 years |
| *cg09686317* | chr1 | 57111122 | PRKAA2 | Island | 0.8533 | 0.194 | 1.09E-05 | Late trauma, 11-17 years |
| *cg08362273* | chr17 | 46719577 | | Island | 1.399 | 0.319 | 1.16E-05 | Late trauma, 11-17 years |
| *cg03315940* | chr6 | 27569167 | | OpenSea | -1.605 | 0.3675 | 1.27E-05 | Late trauma, 11-17 years |
| *cg14663914* | chr19 | 827739 | AZU1 | OpenSea | -2.086 | 0.4786 | 1.31E-05 | Late trauma, 11-17 years |
| *cg05814106* | chr1 | 2224791 | SKI | S_Shelf | -1.1 | 0.2528 | 1.36E-05 | Late trauma, 11-17 years |
| *cg14272822* | chr9 | 98279166 | PTCH1 | Island | 2.611 | 0.6005 | 1.38E-05 | Late trauma, 11-17 years |
| *cg16791444* | chr17 | 71702669 | | OpenSea | -1.384 | 0.319 | 1.43E-05 | Late trauma, 11-17 years |
| *cg02324227* | chr5 | 180591594 | | OpenSea | 1.841 | 0.4261 | 1.55E-05 | Late trauma, 11-17 years |
| *cg03748603* | chr19 | 2494443 | | Island | -0.8501 | 0.1982 | 0.000018 | Late trauma, 11-17 years |
| *cg08791347* | chr10 | 13831250 | FRMD4A | OpenSea | 1.157 | 0.27 | 1.83E-05 | Late trauma, 11-17 years |
| *cg15542924* | chr6 | 163959346 | QKI | OpenSea | -0.7093 | 0.1657 | 1.86E-05 | Late trauma, 11-17 years |
| *cg07942847* | chr14 | 23420757 | HAUS4 | OpenSea | 2.344 | 0.5485 | 1.92E-05 | Late trauma, 11-17 years |
| *cg08528970* | chr4 | 76640579 | | OpenSea | -1.41 | 0.33 | 1.93E-05 | Late trauma, 11-17 years |
| *cg01996004* | chr5 | 180338435 | BTNL8 | OpenSea | -1.073 | 0.2518 | 2.03E-05 | Late trauma, 11-17 years |
| *cg14841628* | chr1 | 16258280 | SPEN | N_Shelf | -1.484 | 0.3497 | 2.2E-05 | Late trauma, 11-17 years |
| *cg10953498* | chr11 | 208469 | RIC8A; BET1L | Island | 1.728 | 0.4074 | 2.23E-05 | Late trauma, 11-17 years |
| *cg06381135* | chr10 | 134505548 | INPP5A | Island | -1.004 | 0.2381 | 2.47E-05 | Late trauma, 11-17 years |
| *cg25618765* | chr22 | 20670922 | | N_Shelf | -1.506 | 0.3574 | 2.5E-05 | Late trauma, 11-17 years |
| *cg25616869* | chr3 | 48443867 | | OpenSea | -2.122 | 0.5035 | 2.51E-05 | Late trauma, 11-17 years |
| *cg13895765* | chr2 | 66803345 | | Island | 1.063 | 0.2531 | 2.67E-05 | Late trauma, 11-17 years |
| *cg22936975* | chr13 | 61989412 | PCDH20 | OpenSea | 1.731 | 0.4125 | 2.7E-05 | Late trauma, 11-17 years |
| *cg05771369* | chr12 | 58021713 | B4GALNT1 | Island | 0.6782 | 0.1617 | 2.75E-05 | Late trauma, 11-17 years |
| *cg16257434* | chr17 | 73571444 | | S_Shelf | -0.9512 | 0.2269 | 2.76E-05 | Late trauma, 11-17 years |
| *cg11597902* | chr17 | 75096239 | | OpenSea | -0.7871 | 0.1889 | 3.1E-05 | Late trauma, 11-17 years |
| *cg19052355* | chr2 | 237076306 | GBX2 | Island | 1.597 | 0.3837 | 3.14E-05 | Late trauma, 11-17 years |
| *cg08032924* | chr16 | 66613096 | CMTM2 | Island | 0.5852 | 0.1407 | 3.21E-05 | Late trauma, 11-17 years |

| | | | | | | | | |
|---|---|---|---|---|---|---|---|---|
| *cg21041594* | chr7 | 65235699 | | Island | 1.617 | 0.3891 | 3.26E-05 | Late trauma, 11-17 years |
| *cg24694691* | chr1 | 111927153 | LOC441897 | OpenSea | -1.558 | 0.3755 | 3.36E-05 | Late trauma, 11-17 years |
| *cg10143823* | chr14 | 28192481 | | OpenSea | 2.142 | 0.5169 | 0.000034 | Late trauma, 11-17 years |
| *cg26195482* | chr14 | 104551553 | ASPG | N_Shore | 1.544 | 0.373 | 3.48E-05 | Late trauma, 11-17 years |
| *cg05232857* | chr6 | 33867321 | | OpenSea | 1.468 | 0.3548 | 3.5E-05 | Late trauma, 11-17 years |
| *cg02675083* | chr1 | 16972917 | MST1P2 | N_Shore | -2.158 | 0.5218 | 3.53E-05 | Late trauma, 11-17 years |
| *cg22827724* | chr6 | 31096189 | PSORS1C1 | OpenSea | -1.085 | 0.263 | 3.7E-05 | Late trauma, 11-17 years |
| *cg23041896* | chr8 | 144462235 | RHPN1 | Island | -0.773 | 0.1875 | 3.73E-05 | Late trauma, 11-17 years |
| *cg18343437* | chr8 | 142528415 | | Island | 0.649 | 0.158 | 4.01E-05 | Late trauma, 11-17 years |
| *cg07560534* | chr9 | 100745354 | ANP32B | Island | -2.175 | 0.531 | 4.2E-05 | Late trauma, 11-17 years |
| *cg01312828* | chr6 | 41648365 | | N_Shelf | -1.132 | 0.2778 | 4.59E-05 | Late trauma, 11-17 years |
| *cg08104283* | chr13 | 113676829 | MCF2L | Island | -0.9143 | 0.2247 | 4.73E-05 | Late trauma, 11-17 years |
| *cg00938816* | chr17 | 8058149 | | N_Shore | -2.03 | 0.4992 | 4.77E-05 | Late trauma, 11-17 years |
| *cg04177395* | chr11 | 78496497 | ODZ4 | OpenSea | 0.7883 | 0.1939 | 4.78E-05 | Late trauma, 11-17 years |
| *cg17403875* | chr14 | 55596356 | LGALS3 | Island | -2.37 | 0.5831 | 4.79E-05 | Late trauma, 11-17 years |
| *cg26428054* | chr12 | 49484058 | DHH | Island | -3.23 | 0.6249 | 2.36E-07 | Abuse only, 0-17 years |
| *cg10529789* | chr17 | 37009658 | SNORA21; RPL23 | Island | -2.726 | 0.5501 | 7.2E-07 | Abuse only, 0-17 years |
| *cg24157392* | chr3 | 112217973 | BTLA | OpenSea | 2.249 | 0.4563 | 8.29E-07 | Abuse only, 0-17 years |
| *cg14663914* | chr19 | 827739 | AZU1 | OpenSea | -2.215 | 0.456 | 1.2E-06 | Abuse only, 0-17 years |
| *cg08528970* | chr4 | 76640579 | | OpenSea | -1.463 | 0.3148 | 3.37E-06 | Abuse only, 0-17 years |
| *cg11182518* | chr5 | 55117965 | | Island | 0.845 | 0.1821 | 3.49E-06 | Abuse only, 0-17 years |
| *cg07992052* | chr20 | 35402295 | DSN1 | Island | -1.861 | 0.4059 | 4.57E-06 | Abuse only, 0-17 years |
| *cg25618765* | chr22 | 20670922 | | N_Shelf | -1.552 | 0.3404 | 5.11E-06 | Abuse only, 0-17 years |
| *cg14672994* | chr17 | 48503057 | ACSF2 | Island | -1.547 | 0.3394 | 5.18E-06 | Abuse only, 0-17 years |
| *cg05640294* | chr19 | 38976794 | RYR1 | Island | -1.572 | 0.3514 | 7.64E-06 | Abuse only, 0-17 years |
| *cg21041594* | chr7 | 65235699 | | Island | 1.678 | 0.3776 | 8.84E-06 | Abuse only, 0-17 years |
| *cg21889703* | chr6 | 136607649 | BCLAF1 | N_Shelf | -2.098 | 0.4738 | 9.49E-06 | Abuse only, 0-17 years |
| *cg10755512* | chr3 | 44666543 | ZNF197 | Island | 2.096 | 0.4747 | 1.02E-05 | Abuse only, 0-17 years |
| *cg19989295* | chr14 | 24641077 | REC8 | Island | 0.9588 | 0.2179 | 1.09E-05 | Abuse only, 0-17 years |

| | | | | | | | | |
|---|---|---|---|---|---|---|---|---|
| *cg01312828* | chr6 | 41648365 | | N_Shelf | -1.175 | 0.2681 | 1.18E-05 | Abuse only, 0-17 years |
| *cg19754901* | chr3 | 87102643 | | S_Shore | -1.434 | 0.3281 | 1.23E-05 | Abuse only, 0-17 years |
| *cg19013753* | chr15 | 75915192 | SNUPN | N_Shelf | 1.56 | 0.3576 | 1.29E-05 | Abuse only, 0-17 years |
| *cg08201736* | chr15 | 79298769 | RASGRF1 | OpenSea | 1.548 | 0.3556 | 1.35E-05 | Abuse only, 0-17 years |
| *cg03315940* | chr6 | 27569167 | | OpenSea | -1.539 | 0.355 | 1.45E-05 | Abuse only, 0-17 years |
| *cg17276036* | chr4 | 26492222 | CCKAR | OpenSea | 1.787 | 0.4141 | 1.6E-05 | Abuse only, 0-17 years |
| *cg10632722* | chr6 | 49833873 | CRISP1 | OpenSea | -1.712 | 0.3974 | 1.66E-05 | Abuse only, 0-17 years |
| *cg14010194* | chr6 | 42152817 | GUCA1B | OpenSea | -1.203 | 0.2795 | 1.66E-05 | Abuse only, 0-17 years |
| *cg16328610* | chr6 | 83777335 | DOPEY1 | Island | -2.12 | 0.4933 | 1.73E-05 | Abuse only, 0-17 years |
| *cg27524192* | chr1 | 228773464 | | Island | -1.476 | 0.3452 | 1.9E-05 | Abuse only, 0-17 years |
| *cg07942847* | chr14 | 23420757 | HAUS4 | OpenSea | 2.269 | 0.5327 | 2.06E-05 | Abuse only, 0-17 years |
| *cg10251328* | chr17 | 47074713 | IGF2BP1 | Island | -2.454 | 0.5798 | 2.32E-05 | Abuse only, 0-17 years |
| *cg16791444* | chr17 | 71702669 | | OpenSea | -1.281 | 0.3042 | 2.55E-05 | Abuse only, 0-17 years |
| *cg00980622* | chr14 | 75884845 | | OpenSea | -1.226 | 0.2916 | 2.62E-05 | Abuse only, 0-17 years |
| *cg22936975* | chr13 | 61989412 | PCDH20 | OpenSea | 1.659 | 0.3954 | 2.73E-05 | Abuse only, 0-17 years |
| *cg16588073* | chr1 | 156647247 | NES | Island | -1.117 | 0.2664 | 2.75E-05 | Abuse only, 0-17 years |
| *cg23041896* | chr8 | 144462235 | RHPN1 | Island | -0.748 | 0.1785 | 2.79E-05 | Abuse only, 0-17 years |
| *cg00464520* | chr10 | 118903060 | | S_Shelf | -1.08 | 0.2588 | 3.02E-05 | Abuse only, 0-17 years |
| *cg08362273* | chr17 | 46719577 | | Island | 1.263 | 0.3031 | 3.07E-05 | Abuse only, 0-17 years |
| *cg05771369* | chr12 | 58021713 | B4GALNT1 | Island | 0.6425 | 0.1542 | 3.11E-05 | Abuse only, 0-17 years |
| *cg10953498* | chr11 | 208469 | RIC8A; BET1L | Island | 1.64 | 0.3939 | 3.12E-05 | Abuse only, 0-17 years |
| *cg02324227* | chr5 | 180591594 | | OpenSea | 1.679 | 0.4046 | 3.34E-05 | Abuse only, 0-17 years |
| *cg05814106* | chr1 | 2224791 | SKI | S_Shelf | -0.9831 | 0.237 | 3.34E-05 | Abuse only, 0-17 years |
| *cg01996004* | chr5 | 180338435 | BTNL8 | OpenSea | -0.9993 | 0.2413 | 3.46E-05 | Abuse only, 0-17 years |
| *cg18343437* | chr8 | 142528415 | | Island | 0.6133 | 0.1487 | 3.71E-05 | Abuse only, 0-17 years |
| *cg09859179* | chr2 | 58655104 | | OpenSea | 1.624 | 0.3949 | 3.91E-05 | Abuse only, 0-17 years |
| *cg05259836* | chr6 | 74290516 | | OpenSea | -1.35 | 0.3284 | 3.93E-05 | Abuse only, 0-17 years |
| *cg19631585* | chr1 | 228773514 | | Island | -1.809 | 0.4402 | 3.98E-05 | Abuse only, 0-17 years |
| *cg25644556* | chr7 | 27209582 | MIR196B | Island | 0.7195 | 0.1752 | 4E-05 | Abuse only, 0-17 years |

| | | | | | | | | |
|---|---|---|---|---|---|---|---|---|
| *cg23018689* | chr1 | 159173540 | DARC | OpenSea | -1.686 | 0.4109 | 4.07E-05 | Abuse only, 0-17 years |
| *cg17403875* | chr14 | 55596356 | LGALS3 | Island | -2.306 | 0.5626 | 4.14E-05 | Abuse only, 0-17 years |
| *cg19414967* | chr8 | 49468296 | | N_Shore | 1.445 | 0.3534 | 4.3E-05 | Abuse only, 0-17 years |
| *cg07631533* | chr16 | 13135565 | SHISA9 | OpenSea | 1.363 | 0.3333 | 4.32E-05 | Abuse only, 0-17 years |
| *cg05899741* | chr4 | 43901084 | | OpenSea | -1.625 | 0.3979 | 4.43E-05 | Abuse only, 0-17 years |
| *cg16257434* | chr17 | 73571444 | | S_Shelf | -0.8814 | 0.2163 | 4.62E-05 | Abuse only, 0-17 years |
| *cg19024989* | chr11 | 2024126 | | N_Shelf | -1.999 | 0.4921 | 4.84E-05 | Abuse only, 0-17 years |
| *cg14855877* | chr17 | 12905834 | ELAC2 | OpenSea | -0.9195 | 0.2264 | 4.88E-05 | Abuse only, 0-17 years |
| *cg02675083* | chr1 | 16972917 | MST1P2 | N_Shore | -2.03 | 0.5003 | 4.98E-05 | Abuse only, 0-17 years |

Genomic locations are reported according to the hg19 reference genome. Gene annotation and CpG island context correspond to the Illumina HumanMethylation450 array annotation.

#### *5. Differentially methylated regions (DMR) analyses*

Region-based analyses were conducted using DMRff for Path A and Path B across the four trauma definitions. Summary counts, top-ranked DMRs, and combined Manhattan-style region plots are reported below.

**Table S4.** *Top differentially methylated regions associated with trauma exposure (Path A)*

| Exposure | Chromosome | Start (bp) | End (bp) | Region width (bp) | CpG sites | Gene(s) | Min CpG p-value | Region p-value | FDR (BH) |
|---|---|---|---|---|---|---|---|---|---|
| *Abuse-only trauma, 0-17 years* | chr6 | 30684837 | 30684905 | 69 | 4 | MDC | $8.1299\times10^{-3}$ | $9.1255\times10^{-10}$ | $4.3741\times10^{-4}$ |
| *Abuse-only trauma, 0-17 years* | chr6 | 32164382 | 32164442 | 61 | 3 | GPSM3; NOTCH4 | $8.5127\times10^{-5}$ | $7.8008\times10^{-8}$ | 0.0374 |
| *Early trauma, 5-10 years* | chrX | 21676344 | 21676593 | 250 | 4 | KLHL34 | $2.5618\times10^{-3}$ | $6.9087\times10^{-18}$ | $3.3345\times10^{-12}$ |
| *Early trauma, 5-10 years* | chr6 | 31940004 | 31940782 | 779 | 32 | DOM3Z STK19 | $8.2127\times10^{-3}$ | $4.6250\times10^{-13}$ | $2.2323\times10^{-7}$ |
| *Early trauma, 5-10 years* | chr6 | 31707202 | 31707922 | 721 | 24 | MSH5 | $4.7171\times10^{-3}$ | $4.7856\times10^{-11}$ | $2.3098\times10^{-5}$ |
| *Early trauma, 5-10 years* | chr1 | 53704012 | 53704033 | 22 | 2 | MAGOH | $1.9450\times10^{-3}$ | $1.0884\times10^{-9}$ | $5.2533\times10^{-4}$ |
| *Early trauma, 5-10 years* | chrX | 148586931 | 148587019 | 89 | 7 | IDS | 0.0195 | $2.8571\times10^{-9}$ | $1.3790\times10^{-3}$ |
| *Late trauma, 11-17 years* | chr20 | 45338083 | 45338396 | 314 | 5 | SLC2A10 | $3.7571\times10^{-3}$ | $9.9868\times10^{-10}$ | $4.7464\times10^{-4}$ |
| *Late trauma, 11-17 years* | chrX | 47483024 | 47483268 | 245 | 4 | NA | 0.0383 | $1.3340\times10^{-8}$ | $6.3400\times10^{-3}$ |
| *Childhood trauma, 0-17 years* | chr19 | 39926698 | 39926862 | 165 | 5 | RPS16 | 0.0181 | $5.3692\times10^{-12}$ | $2.5519\times10^{-6}$ |

**Table S5.** *Top differentially methylated regions associated with psychotic-like experiences (Path B)*

| Exposure | Chromosome | Start (bp) | End (bp) | Region width (bp) | CpG sites | Gene(s) | Min CpG p-value | Region p-value | FDR (BH) |
|---|---|---|---|---|---|---|---|---|---|
| *Abuse-only trauma, 0-17 years* | chrX | 107334809 | 107334922 | 114 | 3 | PSMD10; ATG4A | 0.0146 | $5.6115\times10^{-11}$ | $2.7500\times10^{-5}$ |
| *Abuse-only trauma, 0-17 years* | chr6 | 153323975 | 153324001 | 27 | 3 | MTRF1L | $3.3860\times10^{-3}$ | $1.3651\times10^{-10}$ | $6.6897\times10^{-5}$ |
| *Abuse-only trauma, 0-17 years* | chrX | 70288026 | 70288358 | 333 | 7 | SNX12 | $7.6132\times10^{-3}$ | $3.1251\times10^{-9}$ | $1.5315\times10^{-3}$ |
| *Abuse-only trauma, 0-17 years* | chr3 | 14165888 | 14166327 | 440 | 3 | TMEM43; CHCHD4 | $7.8953\times10^{-4}$ | $4.3659\times10^{-8}$ | 0.0214 |
| *Abuse-only trauma, 0-17 years* | chr2 | 10953115 | 10953237 | 123 | 2 | PDIA6 | $7.1233\times10^{-4}$ | $5.7080\times10^{-8}$ | 0.0280 |
| *Early trauma, 5-10 years* | chr6 | 31865886 | 31866072 | 187 | 11 | EHMT2 | 0.0127 | $2.0333\times10^{-12}$ | $9.8966\times10^{-7}$ |
| *Early trauma, 5-10 years* | chr6 | 32940303 | 32940921 | 619 | 24 | BRD2 | 0.0237 | $4.8506\times10^{-10}$ | $2.3609\times10^{-4}$ |

| | | | | | | | | | |
|---|---|---|---|---|---|---|---|---|---|
| *Early trauma, 5-10 years* | chr6 | 137242316 | 137243249 | 934 | 4 | SLC35D3 | $1.1282\times10^{-3}$ | $1.8288\times10^{-9}$ | $8.9014\times10^{-4}$ |
| *Early trauma, 5-10 years* | chr6 | 153323975 | 153324006 | 32 | 4 | MTRF1L | $5.0232\times10^{-3}$ | $2.2619\times10^{-9}$ | $1.1010\times10^{-3}$ |
| *Early trauma, 5-10 years* | chrX | 18372612 | 18372696 | 85 | 2 | SCML2 | $1.0838\times10^{-3}$ | $8.3810\times10^{-9}$ | $4.0793\times10^{-3}$ |
| *Early trauma, 5-10 years* | chrX | 40017092 | 40017469 | 378 | 3 | BCOR | $8.0103\times10^{-3}$ | $1.4260\times10^{-8}$ | $6.9409\times10^{-3}$ |
| *Early trauma, 5-10 years* | chrX | 107334809 | 107334815 | 7 | 2 | PSMD10; ATG4A | 0.0251 | $7.6590\times10^{-8}$ | 0.0373 |
| *Late trauma, 11-17 years* | chr6 | 31865886 | 31866072 | 187 | 11 | EHMT2 | $9.3044\times10^{-3}$ | $1.2309\times10^{-14}$ | $6.0354\times10^{-9}$ |
| *Late trauma, 11-17 years* | chr6 | 30312939 | 30313380 | 442 | 18 | RPP21 | 0.0209 | $7.6970\times10^{-12}$ | $3.7740\times10^{-6}$ |
| *Late trauma, 11-17 years* | chr6 | 153323975 | 153324006 | 32 | 4 | MTRF1L | $1.1272\times10^{-3}$ | $5.8264\times10^{-11}$ | $2.8568\times10^{-5}$ |
| *Late trauma, 11-17 years* | chr6 | 137242316 | 137243249 | 934 | 4 | SLC35D3 | $2.1558\times10^{-3}$ | $1.0586\times10^{-10}$ | $5.1907\times10^{-5}$ |
| *Late trauma, 11-17 years* | chrX | 107334809 | 107334922 | 114 | 3 | PSMD10; ATG4A | $7.6447\times10^{-3}$ | $1.3799\times10^{-9}$ | $6.7660\times10^{-4}$ |
| *Late trauma, 11-17 years* | chr8 | 23261738 | 23261967 | 230 | 3 | LOXL2 | 0.0191 | $2.3098\times10^{-9}$ | $1.1326\times10^{-3}$ |
| *Late trauma, 11-17 years* | chrX | 40017092 | 40017469 | 378 | 3 | BCOR | $8.3317\times10^{-3}$ | $2.8853\times10^{-8}$ | 0.0141 |
| *Late trauma, 11-17 years* | chr19 | 1479426 | 1480049 | 624 | 6 | C19orf25 | $1.2018\times10^{-3}$ | $4.5483\times10^{-8}$ | 0.0223 |
| *Late trauma, 11-17 years* | chr18 | 56940418 | 56940463 | 46 | 2 | RAX | 0.0104 | $5.2549\times10^{-8}$ | 0.0258 |
| *Late trauma, 11-17 years* | chr2 | 10953115 | 10953237 | 123 | 2 | PDIA6 | $4.4700\times10^{-4}$ | $5.7636\times10^{-8}$ | 0.0283 |
| *Childhood trauma, 0-17 years* | chrX | 128977498 | 128977653 | 156 | 3 | ZDHHC9 | $7.0079\times10^{-4}$ | $7.2166\times10^{-13}$ | $3.5348\times10^{-7}$ |
| *Childhood trauma, 0-17 years* | chr6 | 153323975 | 153324006 | 32 | 4 | MTRF1L | $1.7837\times10^{-3}$ | $1.3961\times10^{-11}$ | $6.8385\times10^{-6}$ |
| *Childhood trauma, 0-17 years* | chrX | 68047833 | 68048015 | 183 | 8 | EFNB1 | $6.2569\times10^{-3}$ | $1.0970\times10^{-9}$ | $5.3731\times10^{-4}$ |
| *Childhood trauma, 0-17 years* | chrX | 107334809 | 107334922 | 114 | 3 | PSMD10; ATG4A | 0.0194 | $3.3438\times10^{-9}$ | $1.6378\times10^{-3}$ |
| *Childhood trauma, 0-17 years* | chrX | 70288026 | 70288358 | 333 | 7 | SNX12 | 0.0105 | $6.4332\times10^{-9}$ | $3.1511\times10^{-3}$ |
| *Childhood trauma, 0-17 years* | chr3 | 14165888 | 14166327 | 440 | 3 | TMEM43; CHCHD4 | $5.7382\times10^{-4}$ | $2.4420\times10^{-8}$ | 0.0120 |
| *Childhood trauma, 0-17 years* | chr2 | 10953115 | 10953237 | 123 | 2 | PDIA6 | $8.5185\times10^{-4}$ | $5.1603\times10^{-8}$ | 0.0253 |

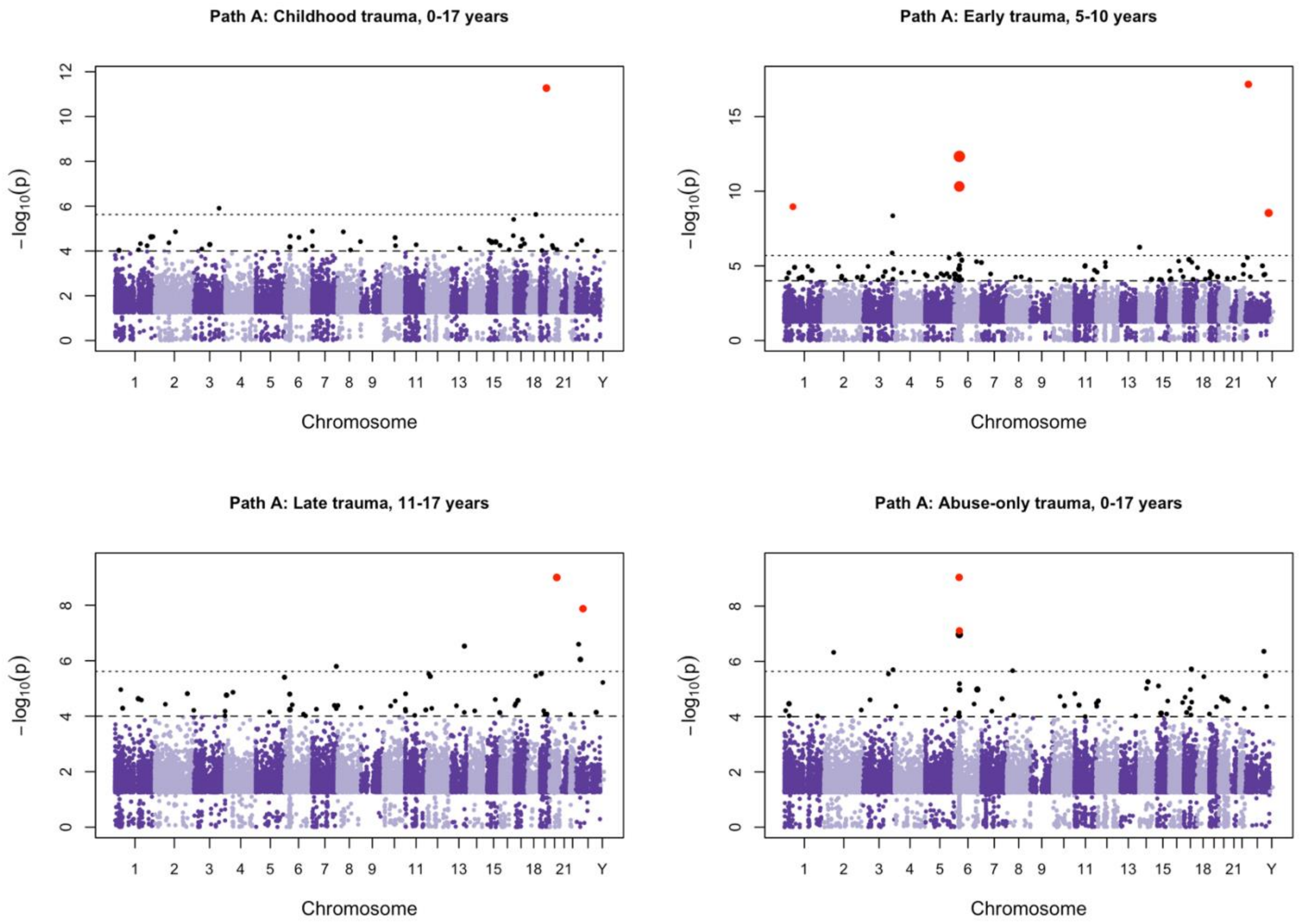


**Figure S4.** *Differentially methylated region (DMR) analyses for Path A.*

Manhattan-style plots of DMRff results for four trauma exposures. Each point represents regions plotted by genomic position and the -$log_{10}$-transformed region p-value. The horizontal dashed line indicates the Bonferroni-corrected significance threshold and a suggestive discovery threshold ($p < 1 \times 10^{-4}$).

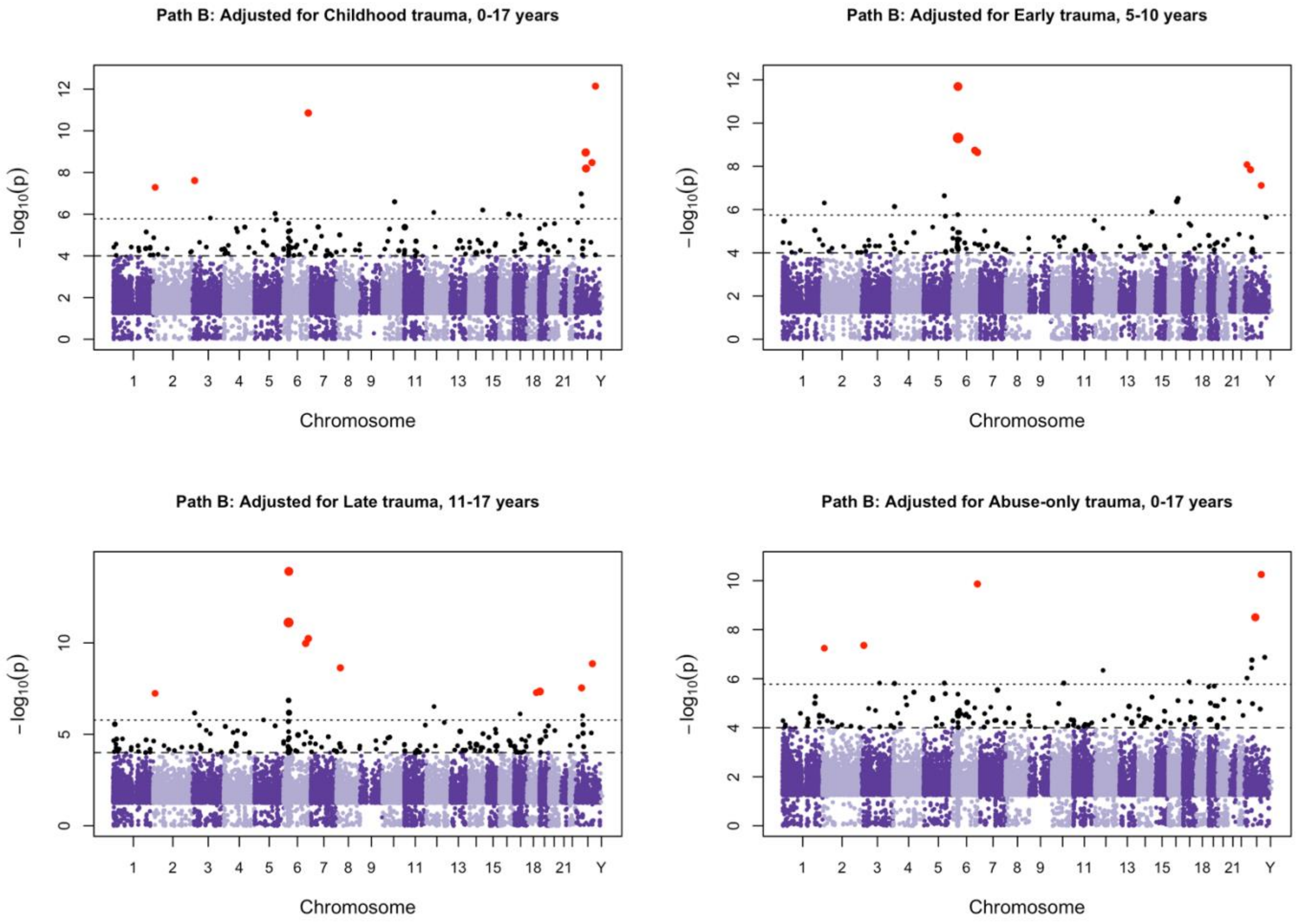


**Figure S5.** *Differentially methylated region (DMR) analyses for Path B.*

Manhattan-style plots of DMRff results for four trauma exposures. Each point represents regions plotted by genomic position and the -$\log_{10}$-transformed region p-value. The horizontal dashed line indicates the Bonferroni-corrected significance threshold and a suggestive discovery threshold ($p < 1 \times 10^{-4}$).

#### *6. Functional Enrichment Analyses*

**Table S6.** *Gene Ontology and KEGG pathway enrichment results for genes annotated to DMRs - Path A*

| Exposure | Database | Ontology | Description | N | DE | P.DE | FDR |
|---|---|---|---|---|---|---|---|
| **Abuse-only trauma, 0-17 years** | GO | BP | response to stress | 3872 | 3 | 0.0101 | 1 |
| **Abuse-only trauma, 0-17 years** | GO | BP | positive regulation of macromolecule metabolic process | 3480 | 3 | 0.0143 | 1 |
| **Abuse-only trauma, 0-17 years** | GO | BP | positive regulation of metabolic process | 3786 | 3 | 0.0182 | 1 |
| **Abuse-only trauma, 0-17 years** | GO | BP | regulation of gene expression | 4952 | 3 | 0.0273 | 1 |
| **Abuse-only trauma, 0-17 years** | GO | BP | positive regulation of transcription by RNA polymerase II | 1208 | 2 | 0.0294 | 1 |
| **Abuse-only trauma, 0-17 years** | GO | BP | gene expression | 6365 | 3 | 0.0376 | 1 |
| **Abuse-only trauma, 0-17 years** | GO | BP | positive regulation of DNA-templated transcription | 1651 | 2 | 0.0486 | 1 |
| **Abuse-only trauma, 0-17 years** | GO | BP | positive regulation of nucleic acid-templated transcription | 1651 | 2 | 0.0486 | 1 |
| **Early childhood trauma, 5-10 years** | GO | BP | nuclear-transcribed mRNA catabolic process | 125 | 2 | 7.88E-04 | 1 |
| **Early childhood trauma, 5-10 years** | GO | BP | mRNA catabolic process | 261 | 2 | 0.00302 | 1 |
| **Early childhood trauma, 5-10 years** | GO | BP | cellular macromolecule catabolic process | 1018 | 3 | 0.00384 | 1 |
| **Early childhood trauma, 5-10 years** | GO | BP | RNA catabolic process | 312 | 2 | 0.0042 | 1 |
| **Early childhood trauma, 5-10 years** | GO | BP | nucleobase-containing compound catabolic process | 439 | 2 | 0.00766 | 1 |
| **Early childhood trauma, 5-10 years** | GO | BP | cellular nitrogen compound catabolic process | 478 | 2 | 0.00845 | 1 |
| **Early childhood trauma, 5-10 years** | GO | BP | heterocycle catabolic process | 480 | 2 | 0.00877 | 1 |
| **Early childhood trauma, 5-10 years** | GO | BP | aromatic compound catabolic process | 497 | 2 | 0.00916 | 1 |

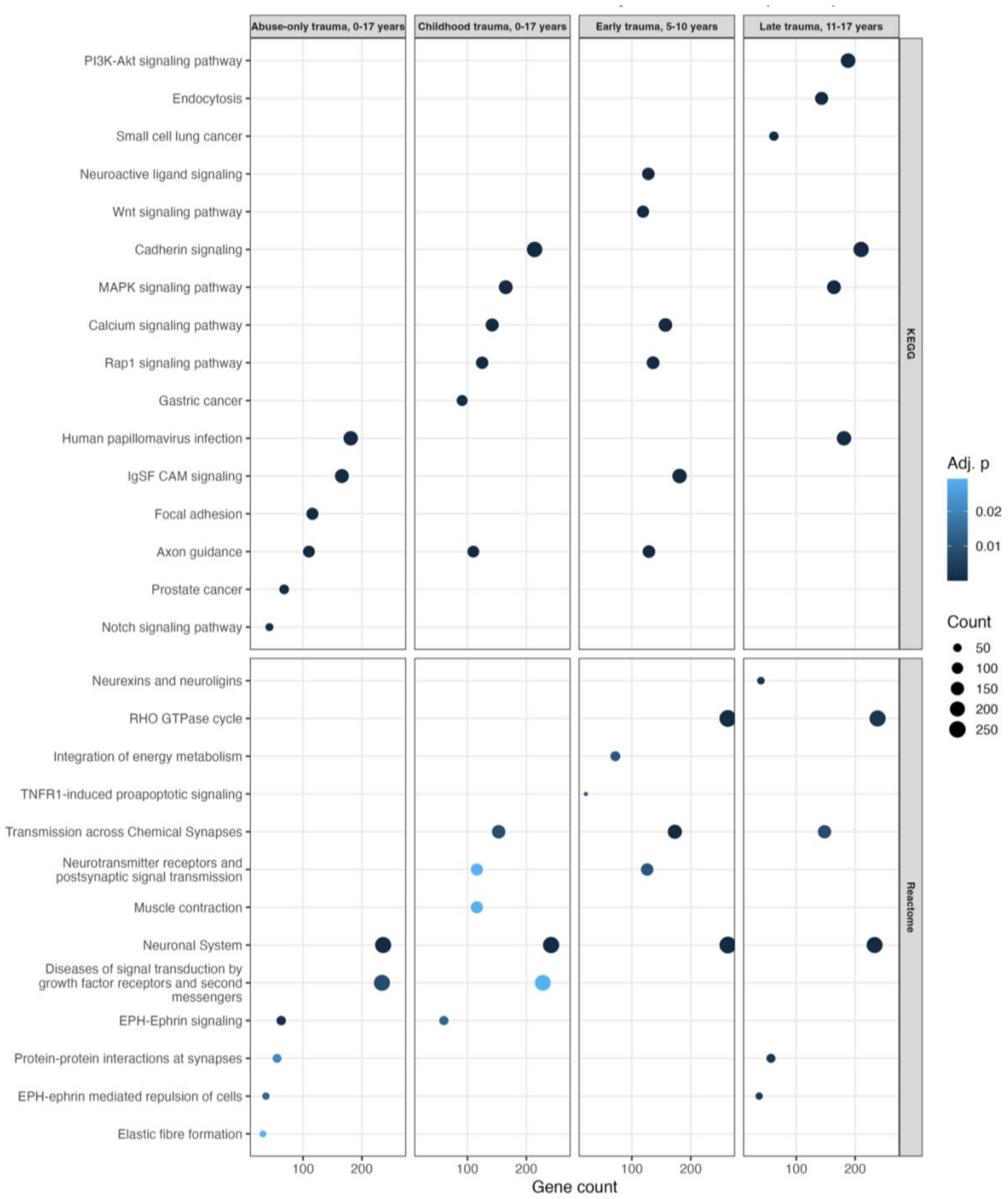


**Figure S6.** *Functional enrichment of trauma-DMRs in Path A.*
Dot plots showing the top enriched KEGG and Reactome pathways for genes mapped to DMRs identified in Path A models across the four trauma definitions. Dot sizes represent gene count and colour indicates adjusted p-value.

**Table S7.** *Gene Ontology and KEGG pathway enrichment results for genes annotated to DMRs - Path B*

| Exposure | Database | Ontology | Description | N | DE | P.DE | FDR |
|---|---|---|---|---|---|---|---|
| **Abuse-only trauma, 0-17 years** | GO | MF | protein-disulfide reductase activity | 34 | 2 | 7.64E-05 | 1 |
| **Abuse-only trauma, 0-17 years** | GO | MF | disulfide oxidoreductase activity | 39 | 2 | 1.05E-04 | 1 |
| **Abuse-only trauma, 0-17 years** | GO | MF | oxidoreductase activity, acting on a sulfur group of donors | 54 | 2 | 2.02E-04 | 1 |
| **Abuse-only trauma, 0-17 years** | GO | BP | protein folding | 209 | 2 | 0.00277 | 1 |
| **Abuse-only trauma, 0-17 years** | GO | CC | endoplasmic reticulum lumen | 309 | 2 | 0.00439 | 1 |
| **Abuse-only trauma, 0-17 years** | GO | BP | protein localization | 2432 | 4 | 0.00547 | 1 |
| **Abuse-only trauma, 0-17 years** | GO | BP | cellular macromolecule localization | 2442 | 4 | 0.00554 | 1 |
| **Abuse-only trauma, 0-17 years** | GO | BP | protein maturation | 327 | 2 | 0.00557 | 1 |
| **Early childhood trauma, 5-10 years** | GO | BP | regulation of histone methylation | 57 | 2 | 3.33E-04 | 1 |
| **Early childhood trauma, 5-10 years** | GO | BP | histone lysine methylation | 103 | 2 | 9.97E-04 | 1 |
| **Early childhood trauma, 5-10 years** | GO | BP | peptidyl-lysine methylation | 121 | 2 | 0.00127 | 1 |
| **Early childhood trauma, 5-10 years** | GO | BP | histone methylation | 132 | 2 | 0.00155 | 1 |
| **Early childhood trauma, 5-10 years** | GO | BP | regulation of histone modification | 141 | 2 | 0.00176 | 1 |
| **Early childhood trauma, 5-10 years** | GO | BP | chromatin organization | 631 | 3 | 0.00198 | 1 |
| **Early childhood trauma, 5-10 years** | GO | MF | transcription factor binding | 593 | 3 | 0.00205 | 1 |
| **Early childhood trauma, 5-10 years** | GO | BP | protein alkylation | 179 | 2 | 0.00261 | 1 |
| **Early childhood trauma, 5-10 years** | KEGG | KEGG | Polycomb repressive complex | 82 | 2 | 7.82E-04 | 0.289 |
| **Adolescent trauma, 11-17 years** | GO | BP | regulation of histone methylation | 57 | 2 | 6.16E-04 | 1 |
| **Adolescent trauma, 11-17 years** | GO | BP | peptidyl-lysine modification | 369 | 3 | 0.00132 | 1 |
| **Adolescent trauma, 11-17 years** | GO | BP | histone lysine methylation | 103 | 2 | 0.00215 | 1 |
| **Adolescent trauma, 11-17 years** | GO | BP | peptidyl-lysine methylation | 121 | 2 | 0.00267 | 1 |
| **Adolescent trauma, 11-17 years** | GO | BP | histone methylation | 132 | 2 | 0.00311 | 1 |

| | | | | | | | |
|---|---|---|---|---|---|---|---|
| **Adolescent trauma, 11-17 years** | GO | BP | negative regulation of transcription by RNA polymerase II | 938 | 4 | 0.00324 | 1 |
| **Adolescent trauma, 11-17 years** | GO | BP | regulation of histone modification | 141 | 2 | 0.00393 | 1 |
| **Adolescent trauma, 11-17 years** | GO | BP | protein alkylation | 179 | 2 | 0.0052 | 1 |
| **Childhood trauma, 0-17 years** | GO | MF | protein-disulfide reductase activity | 34 | 2 | 1.28E-04 | 1 |
| **Childhood trauma, 0-17 years** | GO | MF | disulfide oxidoreductase activity | 39 | 2 | 1.73E-04 | 1 |
| **Childhood trauma, 0-17 years** | GO | BP | peptidyl-cysteine modification | 49 | 2 | 2.7E-04 | 1 |
| **Childhood trauma, 0-17 years** | GO | MF | oxidoreductase activity, acting on a sulfur group of donors | 54 | 2 | 3.37E-04 | 1 |
| **Childhood trauma, 0-17 years** | GO | BP | protein localization | 2432 | 5 | 0.00275 | 1 |
| **Childhood trauma, 0-17 years** | GO | BP | cellular macromolecule localization | 2442 | 5 | 0.00279 | 1 |
| **Childhood trauma, 0-17 years** | GO | BP | localization within membrane | 664 | 3 | 0.00302 | 1 |
| **Childhood trauma, 0-17 years** | GO | BP | protein transport | 1547 | 4 | 0.00409 | 1 |

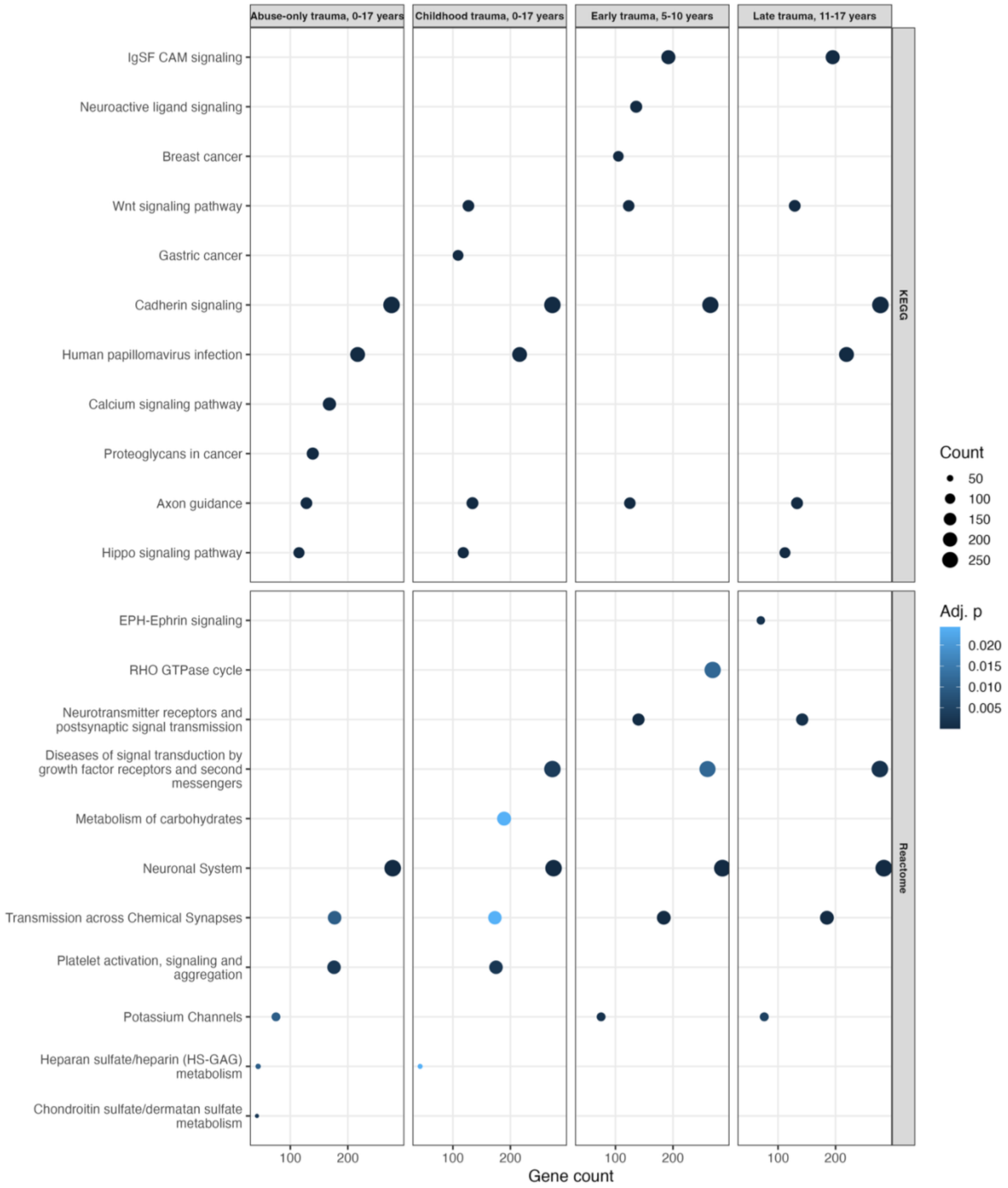


**Figure S7.** *Functional enrichment of PLEs-DMRs in Path B.*
Dot plots showing the top enriched KEGG and Reactome pathways for genes mapped to DMRs identified in Path B models across the four trauma definitions. Dot sizes represent gene count and colour indicates adjusted p-value.

#### *7. Cannabis interaction analyses*

Interaction EWAS models examining moderation by frequency of cannabis use identified several CpG sites showing evidence of trauma x cannabis interaction effects (**Figure S9**). Differentially methylated region analyses identified candidate regions associated with interaction effects across multiple trauma measures (**Table S12**).

##### *7.1. Model diagnostics*

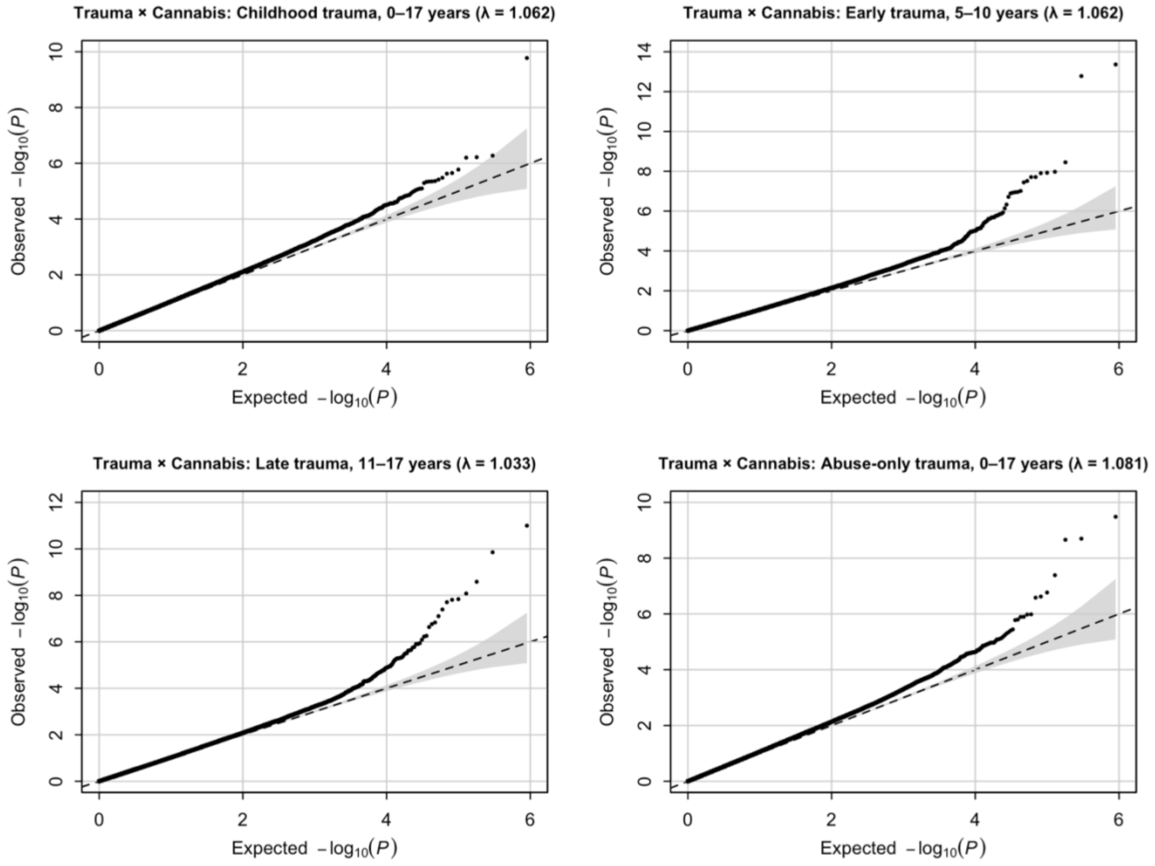


**Figure S8.** *QQ plots for cannabis interaction analyses.*
The dashed line indicates the null expectation and the shaded region the 95% confidence envelope. Genomic inflation factors (λ) are reported in each panel.

*7.2. EWAS interaction results*

**Table S8.** *Top CpG sites showing suggestive evidence of interaction (cumulative childhood trauma)*

| CpG | Chromosome | Position (bp) | Gene(s) | Gene region | CpG island relation | Effect | SE | Statistic | P | FDR |
|---|---|---|---|---|---|---|---|---|---|---|
| *cg20383084* | chr1 | 26325121 | PAFAH2 | TSS1500 | S_Shore | -0.1112 | 0.0173 | -6.4391 | $1.68\times10^{-10}$ | $7.52\times10^{-05}$ |
| *cg07840472* | chr14 | 36278484 | RALGAPA1 | TSS200 | Island | -0.0631 | 0.0125 | -5.0404 | $5.29\times10^{-07}$ | $7.03\times10^{-02}$ |
| *cg08298794* | chr2 | 1.98E+08 | | | OpenSea | -0.1223 | 0.0244 | -5.0146 | $6.03\times10^{-07}$ | $7.03\times10^{-02}$ |
| *cg26320410* | chr3 | 9851826 | TTLL3 | TSS200 | Island | 0.0408 | 0.0082 | 5.0073 | $6.26\times10^{-07}$ | $7.03\times10^{-02}$ |
| *cg11722699* | chr18 | 28622813 | DSC3 | TSS200 | Island | 0.0988 | 0.0205 | 4.81 | $1.68\times10^{-06}$ | $1.51\times10^{-01}$ |
| *ch.X.258064R* | chrX | 16776121 | SYAP1 | Body | OpenSea | -0.1361 | 0.0286 | -4.7521 | $2.23\times10^{-06}$ | $1.51\times10^{-01}$ |
| *cg04039225* | chr3 | 169629516 | SAMD7 | 5'UTR; 1stExon | OpenSea | -0.0496 | 0.0105 | -4.7411 | $2.36\times10^{-06}$ | $1.51\times10^{-01}$ |
| *cg26343001* | chr12 | 115104274 | | | N_Shore | 0.1113 | 0.0238 | 4.6708 | $3.31\times10^{-06}$ | $1.60\times10^{-01}$ |
| *cg08197226* | chr15 | 49658975 | C15orf33 | Body | OpenSea | 0.0935 | 0.0202 | 4.6397 | $3.84\times10^{-06}$ | $1.60\times10^{-01}$ |
| *cg08486065* | chr19 | 3464875 | | | Island | 0.0607 | 0.0132 | 4.6135 | $4.34\times10^{-06}$ | $1.60\times10^{-01}$ |
| *cg09035925* | chr20 | 19915769 | RIN2 | Body | OpenSea | -0.095 | 0.0206 | -4.6079 | $4.46\times10^{-06}$ | $1.60\times10^{-01}$ |
| *cg18705408* | chr2 | 20212524 | MATN3 | TSS200 | Island | 0.0798 | 0.0173 | 4.6056 | $4.51\times10^{-06}$ | $1.60\times10^{-01}$ |
| *cg08511716* | chr5 | 94621030 | MCTP1 | TSS1500 | Island | 0.0686 | 0.0149 | 4.5994 | $4.64\times10^{-06}$ | $1.60\times10^{-01}$ |
| *cg24109894* | chr1 | 36108212 | PSMB2 | TSS1500 | S_Shore | -0.0832 | 0.0182 | -4.5802 | $5.08\times10^{-06}$ | $1.63\times10^{-01}$ |
| *cg07300574* | chr12 | 132502636 | EP400 | Body | OpenSea | 0.0706 | 0.0157 | 4.4835 | $7.98\times10^{-06}$ | $2.25\times10^{-01}$ |
| *cg13041894* | chr22 | 47609321 | | | Island | -0.1316 | 0.0294 | -4.4785 | $8.16\times10^{-06}$ | $2.25\times10^{-01}$ |
| *cg24805759* | chr17 | 77997760 | TBC1D16 | 5'UTR | Island | 0.1613 | 0.0361 | 4.4694 | $8.53\times10^{-06}$ | $2.25\times10^{-01}$ |
| *cg06081306* | chr17 | 37760830 | NEUROD2 | 3'UTR | N_Shore | -0.0585 | 0.0131 | -4.4547 | $9.11\times10^{-06}$ | $2.27\times10^{-01}$ |
| *cg04050463* | chr6 | 170602486 | | | N_Shore | -0.0674 | 0.0152 | -4.4366 | $9.92\times10^{-06}$ | $2.34\times10^{-01}$ |
| *cg20163689* | chr2 | 13088414 | | | OpenSea | -0.0759 | 0.0172 | -4.4186 | $1.07\times10^{-05}$ | $2.41\times10^{-01}$ |
| *cg23239754* | chr6 | 27760147 | | | OpenSea | -0.0505 | 0.0115 | -4.3965 | $1.19\times10^{-05}$ | $2.54\times10^{-01}$ |
| *cg14252492* | chr11 | 44332224 | ALX4 | TSS1500 | N_Shore | -0.0829 | 0.019 | -4.3661 | $1.36\times10^{-05}$ | $2.63\times10^{-01}$ |
| *cg19103000* | chr3 | 9851819 | TTLL3 | TSS200 | Island | 0.0375 | 0.0086 | 4.3614 | $1.39\times10^{-05}$ | $2.63\times10^{-01}$ |
| *cg14602982* | chr11 | 119599176 | PVRL1 | Body | Island | 0.0475 | 0.0109 | 4.3509 | $1.46\times10^{-05}$ | $2.63\times10^{-01}$ |
| *cg24701575* | chr11 | 31845389 | | | N_Shore | -0.1041 | 0.0239 | -4.35 | $1.47\times10^{-05}$ | $2.63\times10^{-01}$ |

| | | | | | | | | | | |
|---|---|---|---|---|---|---|---|---|---|---|
| *cg13938035* | chr9 | 116346435 | RGS3 | Body; 5'UTR | Island | -0.1121 | 0.0259 | -4.3366 | $1.56{\times}10^{-05}$ | $2.69{\times}10^{-01}$ |
| *cg23350274* | chr14 | 68188663 | RDH12 | 5'UTR | OpenSea | -0.0639 | 0.0148 | -4.3254 | $1.64{\times}10^{-05}$ | $2.69{\times}10^{-01}$ |
| *cg23687000* | chr21 | 38071673 | SIM2 | TSS1500 | Island | 0.0756 | 0.0175 | 4.3117 | $1.74{\times}10^{-05}$ | $2.69{\times}10^{-01}$ |
| *cg13299942* | chr12 | 119798505 | CCDC60 | Body | OpenSea | -0.0314 | 0.0073 | -4.3102 | $1.75{\times}10^{-05}$ | $2.69{\times}10^{-01}$ |
| *cg23681730* | chr11 | 35683551 | TRIM44 | TSS1500 | N_Shore | -0.0986 | 0.0229 | -4.3042 | $1.80{\times}10^{-05}$ | $2.69{\times}10^{-01}$ |
| *cg09379485* | chr19 | 53758297 | ZNF677 | TSS200 | S_Shore | 0.1075 | 0.0251 | 4.2859 | $1.95{\times}10^{-05}$ | $2.83{\times}10^{-01}$ |
| *cg24818939* | chrX | 101906109 | GPRASP1 | TSS200 | Island | 0.0807 | 0.019 | 4.2534 | $2.25{\times}10^{-05}$ | $2.98{\times}10^{-01}$ |
| *cg21512185* | chr2 | 39005322 | GEMIN6 | TSS200 | OpenSea | 0.0844 | 0.0199 | 4.2351 | $2.44{\times}10^{-05}$ | $2.98{\times}10^{-01}$ |
| *cg22090592* | chr14 | 105174642 | INF2 | Body | OpenSea | 0.0538 | 0.0127 | 4.2351 | $2.44{\times}10^{-05}$ | $2.98{\times}10^{-01}$ |
| *cg27441486* | chr1 | 201708522 | NAV1 | TSS1500; Body | N_Shore | 0.1043 | 0.0246 | 4.2337 | $2.46{\times}10^{-05}$ | $2.98{\times}10^{-01}$ |
| *cg17301223* | chr8 | 145106438 | OPLAH | Body | Island | -0.0542 | 0.0129 | -4.2149 | $2.67{\times}10^{-05}$ | $2.98{\times}10^{-01}$ |
| *cg26101410* | chr4 | 174451564 | HAND2; NBLA00301 | TSS200 | N_Shore | -0.0842 | 0.02 | -4.2119 | $2.70{\times}10^{-05}$ | $2.98{\times}10^{-01}$ |
| *cg00255699* | chr1 | 40307257 | TRIT1 | 3'UTR | OpenSea | -0.0661 | 0.0157 | -4.212 | $2.70{\times}10^{-05}$ | $2.98{\times}10^{-01}$ |
| *cg02337166* | chr17 | 38257303 | NR1D1 | TSS1500 | S_Shore | -0.0532 | 0.0126 | -4.2091 | $2.74{\times}10^{-05}$ | $2.98{\times}10^{-01}$ |
| *cg24946736* | chr19 | 7542181 | PEX11G | Body | Island | -0.0778 | 0.0185 | -4.2085 | $2.74{\times}10^{-05}$ | $2.98{\times}10^{-01}$ |
| *cg02642223* | chr17 | 38109030 | | | OpenSea | -0.0356 | 0.0085 | -4.206 | $2.77{\times}10^{-05}$ | $2.98{\times}10^{-01}$ |
| *cg02444695* | chr6 | 148950185 | | | OpenSea | -0.0627 | 0.0149 | -4.1961 | $2.90{\times}10^{-05}$ | $2.98{\times}10^{-01}$ |
| *cg18500368* | chr12 | 56509603 | RPL41 | TSS1500 | N_Shelf | -0.0747 | 0.0178 | -4.1893 | $2.98{\times}10^{-05}$ | $2.98{\times}10^{-01}$ |
| *cg20142390* | chr21 | 40564147 | BRWD1 | 3'UTR; Body | OpenSea | -0.0431 | 0.0103 | -4.186 | $3.03{\times}10^{-05}$ | $2.98{\times}10^{-01}$ |
| *cg05736378* | chr16 | 89283996 | ZNF778 | TSS200 | Island | 0.0476 | 0.0114 | 4.1807 | $3.10{\times}10^{-05}$ | $2.98{\times}10^{-01}$ |
| *cg13922935* | chr15 | 42804870 | SNAP23 | Body | OpenSea | 0.0462 | 0.011 | 4.1788 | $3.12{\times}10^{-05}$ | $2.98{\times}10^{-01}$ |
| *cg21123573* | chr12 | 120740055 | SIRT4 | TSS200 | OpenSea | -0.1158 | 0.0277 | -4.1767 | $3.16{\times}10^{-05}$ | $2.98{\times}10^{-01}$ |
| *cg00013899* | chr1 | 64992433 | CACHD1 | Body | OpenSea | 0.0734 | 0.0176 | 4.1742 | $3.19{\times}10^{-05}$ | $2.98{\times}10^{-01}$ |
| *cg09719477* | chr11 | 113930430 | ZBTB16 | 1stExon; TSS1500; 5'UTR | Island | 0.0399 | 0.0096 | 4.1644 | $3.33{\times}10^{-05}$ | $3.01{\times}10^{-01}$ |
| *cg01382502* | chr16 | 22252504 | EEF2K | Body | OpenSea | -0.1139 | 0.0274 | -4.1628 | $3.35{\times}10^{-05}$ | $3.01{\times}10^{-01}$ |
| *cg27514608* | chr4 | 23892540 | PPARGC1A | TSS1500 | OpenSea | -0.0611 | 0.0147 | -4.1485 | $3.56{\times}10^{-05}$ | $3.13{\times}10^{-01}$ |
| *cg05923101* | chr21 | 45876770 | LRRC3 | Body | Island | -0.097 | 0.0235 | -4.1277 | $3.89{\times}10^{-05}$ | $3.24{\times}10^{-01}$ |
| *cg07963147* | chr3 | 141086820 | ZBTB38 | 5'UTR | OpenSea | 0.0941 | 0.0228 | 4.1271 | $3.90{\times}10^{-05}$ | $3.24{\times}10^{-01}$ |
| *cg22524998* | chr19 | 17877846 | FCHO1 | Body | S_Shore | 0.0762 | 0.0185 | 4.1201 | $4.02{\times}10^{-05}$ | $3.24{\times}10^{-01}$ |

| | | | | | | | | | | |
|---|---|---|---|---|---|---|---|---|---|---|
| *cg17263026* | chr2 | 121453143 | | | OpenSea | -0.1409 | 0.0342 | -4.1203 | $4.02\times10^{-05}$ | $3.24\times10^{-01}$ |
| *cg17264818* | chr2 | 95612985 | | | OpenSea | 0.0786 | 0.0191 | 4.1195 | $4.04\times10^{-05}$ | $3.24\times10^{-01}$ |
| *cg26363272* | chr2 | 240904871 | NDUFA10 | Body | OpenSea | 0.1462 | 0.0355 | 4.1135 | $4.17\times10^{-05}$ | $3.28\times10^{-01}$ |
| *cg05999287* | chr7 | 116312265 | MET | TSS200 | Island | 0.0474 | 0.0115 | 4.1041 | $4.31\times10^{-05}$ | $3.32\times10^{-01}$ |
| *ch.1.117057666F* | chr1 | 117256143 | | | OpenSea | 0.0884 | 0.0216 | 4.1005 | $4.37\times10^{-05}$ | $3.32\times10^{-01}$ |
| *cg01418667* | chr20 | 3387969 | C20orf194 | Body | Island | 0.0403 | 0.0098 | 4.0969 | $4.44\times10^{-05}$ | $3.32\times10^{-01}$ |
| *cg26924209* | chr1 | 193074740 | GLRX2 | 1stExon; TSS200 | Island | -0.0445 | 0.0109 | -4.0858 | $4.66\times10^{-05}$ | $3.41\times10^{-01}$ |
| *cg10163222* | chr19 | 45004637 | ZNF180 | TSS200 | Island | 0.0447 | 0.011 | 4.0799 | $4.77\times10^{-05}$ | $3.41\times10^{-01}$ |
| *cg21430685* | chr8 | 29387008 | | | OpenSea | 0.0854 | 0.0209 | 4.0791 | $4.79\times10^{-05}$ | $3.41\times10^{-01}$ |

Genomic locations are reported according to the hg19 reference genome. Gene annotation and CpG island context correspond to the Illumina HumanMethylation450 array annotation.

**Table S9.** *Top CpG sites showing suggestive evidence of interaction (early childhood trauma)*

| CpG | Chromosome | Position (bp) | Gene(s) | Gene region | CpG island relation | Effect | SE | Statistic | P | FDR |
|---|---|---|---|---|---|---|---|---|---|---|
| cg26320410 | chr3 | 9851826 | TTLL3 | TSS200 | Island | 0.0825 | 0.0108 | 7.6346 | $4.35\times10^{-14}$ | $1.95\times10^{-08}$ |
| cg19103000 | chr3 | 9851819 | TTLL3 | TSS200 | Island | 0.0848 | 0.0114 | 7.4527 | $1.66\times10^{-13}$ | $3.74\times10^{-08}$ |
| cg08511716 | chr5 | 94621030 | MCTP1 | TSS1500 | Island | 0.1134 | 0.0191 | 5.9451 | $3.54\times10^{-09}$ | $5.30\times10^{-04}$ |
| cg14252492 | chr11 | 44332224 | ALX4 | TSS1500 | N_Shore | -0.1438 | 0.025 | -5.7576 | $1.06\times10^{-08}$ | $9.36\times10^{-04}$ |
| cg25688568 | chr18 | 6730036 | | | Island | 0.1193 | 0.0208 | 5.7386 | $1.18\times10^{-08}$ | $9.36\times10^{-04}$ |
| cg23632875 | chr10 | 133110244 | TCERG1L | TSS1500 | Island | 0.1027 | 0.0179 | 5.7293 | $1.25\times10^{-08}$ | $9.36\times10^{-04}$ |
| cg13118906 | chr17 | 61524292 | CYB561 | TSS1500 | Island | 0.0745 | 0.0132 | 5.6516 | $1.95\times10^{-08}$ | $1.10\times10^{-03}$ |
| cg13928709 | chr5 | 176237221 | UNC5A | TSS1500 | Island | 0.1045 | 0.0185 | 5.6505 | $1.96\times10^{-08}$ | $1.10\times10^{-03}$ |
| cg02882381 | chr11 | 69590332 | FGF4 | TSS200 | Island | 0.0732 | 0.0131 | 5.5713 | $3.07\times10^{-08}$ | $1.53\times10^{-03}$ |
| cg24199112 | chr11 | 69590328 | FGF4 | TSS200 | Island | 0.0781 | 0.0141 | 5.5429 | $3.60\times10^{-08}$ | $1.62\times10^{-03}$ |
| cg14940308 | chr2 | 73144644 | EMX1 | 5'UTR; 1stExon | Island | 0.0918 | 0.0171 | 5.3614 | $9.76\times10^{-08}$ | $3.81\times10^{-03}$ |
| cg20629468 | chr10 | 81664583 | | | Island | 0.0803 | 0.015 | 5.3413 | $1.09\times10^{-07}$ | $3.81\times10^{-03}$ |
| cg24701575 | chr11 | 31845389 | | | N_Shore | -0.1703 | 0.0319 | -5.3348 | $1.13\times10^{-07}$ | $3.81\times10^{-03}$ |
| cg17353893 | chr7 | 73753326 | CLIP2 | Body | Island | -0.1521 | 0.0286 | -5.3248 | $1.19\times10^{-07}$ | $3.81\times10^{-03}$ |

| | | | | | | | | | | |
|---|---|---|---|---|---|---|---|---|---|---|
| cg22654039 | chr20 | 11871282 | BTBD3 | TSS200 | N_Shore | 0.0731 | 0.0138 | 5.3104 | $1.28\times10^{-07}$ | $3.85\times10^{-03}$ |
| cg00789545 | chr14 | 102770909 | RAGE | Body | N_Shore | -0.1159 | 0.0221 | -5.2349 | $1.92\times10^{-07}$ | $5.40\times10^{-03}$ |
| cg01728704 | chr16 | 20818015 | LOC81691; ERI2 | 1stExon; 5'UTR; TSS1500 | Island | -0.0461 | 0.0091 | -5.0639 | $4.70\times10^{-07}$ | $1.24\times10^{-02}$ |
| cg26583481 | chr10 | 135139100 | CALY | 3'UTR | Island | 0.068 | 0.0137 | 4.9785 | $7.27\times10^{-07}$ | $1.81\times10^{-02}$ |
| cg21798447 | chr4 | 85504053 | CDS1 | TSS200 | Island | 0.0658 | 0.0135 | 4.882 | $1.18\times10^{-06}$ | $2.79\times10^{-02}$ |
| cg08197226 | chr15 | 49658975 | C15orf33 | Body | OpenSea | 0.1309 | 0.027 | 4.8509 | $1.38\times10^{-06}$ | $3.03\times10^{-02}$ |
| cg23866687 | chr11 | 96026956 | MAML2 | Body | OpenSea | -0.2248 | 0.0464 | -4.847 | $1.42\times10^{-06}$ | $3.03\times10^{-02}$ |
| cg05126421 | chr7 | 26726046 | SKAP2 | Body | OpenSea | -0.1321 | 0.0274 | -4.8235 | $1.58\times10^{-06}$ | $3.23\times10^{-02}$ |
| cg10194160 | chr6 | 30071387 | TRIM31 | Body | Island | 0.0723 | 0.015 | 4.812 | $1.67\times10^{-06}$ | $3.26\times10^{-02}$ |
| cg26313647 | chr1 | 8085795 | ERRFI1 | 5'UTR | Island | 0.0991 | 0.0206 | 4.7991 | $1.78\times10^{-06}$ | $3.33\times10^{-02}$ |
| cg23639399 | chr10 | 135192176 | PAOX | TSS1500 | Island | -0.0577 | 0.0121 | -4.7714 | $2.04\times10^{-06}$ | $3.46\times10^{-02}$ |
| cg00261781 | chr2 | 20212517 | MATN3 | TSS200 | Island | 0.1089 | 0.0228 | 4.7709 | $2.04\times10^{-06}$ | $3.46\times10^{-02}$ |
| cg18705408 | chr2 | 20212524 | MATN3 | TSS200 | Island | 0.1109 | 0.0233 | 4.7669 | $2.08\times10^{-06}$ | $3.46\times10^{-02}$ |
| cg14602982 | chr11 | 119599176 | PVRL1 | Body | Island | 0.0695 | 0.0146 | 4.7459 | $2.30\times10^{-06}$ | $3.68\times10^{-02}$ |
| cg18899035 | chr14 | 20811563 | PARP2; RPPH1 | TSS1500; Body | OpenSea | -0.1642 | 0.0347 | -4.7341 | $2.44\times10^{-06}$ | $3.68\times10^{-02}$ |
| cg06340552 | chr4 | 142054329 | RNF150 | 1stExon; 5'UTR | Island | 0.0581 | 0.0123 | 4.7322 | $2.46\times10^{-06}$ | $3.68\times10^{-02}$ |
| cg05923101 | chr21 | 45876770 | LRRC3 | Body | Island | -0.1472 | 0.0314 | -4.6937 | $2.97\times10^{-06}$ | $4.29\times10^{-02}$ |
| cg20467168 | chr1 | 76081408 | | | Island | 0.06 | 0.0128 | 4.6782 | $3.19\times10^{-06}$ | $4.48\times10^{-02}$ |
| cg10943191 | chr1 | 1850414 | TMEM52 | Body | Island | 0.0535 | 0.0115 | 4.6548 | $3.57\times10^{-06}$ | $4.86\times10^{-02}$ |
| cg11838152 | chr13 | 102359110 | ITGBL1 | Body | OpenSea | -0.0823 | 0.0177 | -4.6437 | $3.77\times10^{-06}$ | $4.97\times10^{-02}$ |
| cg06695611 | chr2 | 180726328 | ZNF385B; MIR1258 | TSS200; TSS1500 | Island | 0.067 | 0.0145 | 4.6224 | $4.17\times10^{-06}$ | $5.35\times10^{-02}$ |
| cg27644733 | chr16 | 9857216 | GRIN2A | Body | OpenSea | -0.0979 | 0.0214 | -4.5677 | $5.40\times10^{-06}$ | $6.73\times10^{-02}$ |
| cg14233374 | chr21 | 47813461 | PCNT | Body | Island | 0.1025 | 0.0225 | 4.5537 | $5.76\times10^{-06}$ | $6.99\times10^{-02}$ |
| cg23318990 | chr6 | 31599289 | BAT2 | Body | N_Shore | 0.0939 | 0.0208 | 4.5107 | $7.04\times10^{-06}$ | $8.21\times10^{-02}$ |
| cg07057670 | chr3 | 185000824 | | | Island | 0.0995 | 0.0221 | 4.5036 | $7.28\times10^{-06}$ | $8.21\times10^{-02}$ |
| cg09187098 | chr7 | 1913848 | MAD1L1 | Body | Island | -0.0663 | 0.0147 | -4.5019 | $7.33\times10^{-06}$ | $8.21\times10^{-02}$ |
| cg09860601 | chr1 | 1374601 | VWA1 | Body; 3'UTR | Island | -0.1137 | 0.0253 | -4.4972 | $7.49\times10^{-06}$ | $8.21\times10^{-02}$ |
| cg19544459 | chr7 | 22123197 | | | S_Shore | 0.0949 | 0.0211 | 4.4903 | $7.74\times10^{-06}$ | $8.27\times10^{-02}$ |
| cg07507257 | chr18 | 21112105 | NPC1 | 3'UTR | OpenSea | 0.0907 | 0.0202 | 4.4852 | $7.92\times10^{-06}$ | $8.27\times10^{-02}$ |

| | | | | | | | | | | |
|---|---|---|---|---|---|---|---|---|---|---|
| cg01367666 | chr16 | 2052269 | ZNF598 | Body | Island | 0.0925 | 0.0208 | 4.4545 | $9.13\times10^{-06}$ | $9.24\times10^{-02}$ |
| cg15654121 | chr1 | 34630944 | CSMD2 | Body | Island | 0.0546 | 0.0123 | 4.4433 | $9.61\times10^{-06}$ | $9.24\times10^{-02}$ |
| cg01046309 | chr16 | 18995391 | TMC7 | TSS1500; 1stExon | Island | 0.062 | 0.014 | 4.4407 | $9.72\times10^{-06}$ | $9.24\times10^{-02}$ |
| cg03650946 | chr7 | 27154562 | HOXA3 | 5'UTR; TSS1500; 5'UTR | N_Shore | 0.1196 | 0.0269 | 4.4403 | $9.75\times10^{-06}$ | $9.24\times10^{-02}$ |
| cg16890428 | chr19 | 55866412 | FAM71E2; COX6B2 | 3'UTR; TSS1500 | S_Shore | 0.0737 | 0.0166 | 4.4371 | $9.88\times10^{-06}$ | $9.24\times10^{-02}$ |
| cg11281641 | chr2 | 171674855 | GAD1 | 5'UTR | Island | 0.0718 | 0.0162 | 4.4282 | $1.03\times10^{-05}$ | $9.26\times10^{-02}$ |
| cg12648600 | chr6 | 30585819 | MRPS18B; PPP1R10 | Body; TSS1500 | S_Shore | -0.0728 | 0.0165 | -4.4192 | $1.07\times10^{-05}$ | $9.26\times10^{-02}$ |
| cg04743859 | chr11 | 67817488 | TCIRG1 | Body | OpenSea | -0.1414 | 0.032 | -4.4173 | $1.08\times10^{-05}$ | $9.26\times10^{-02}$ |
| cg03517073 | chr2 | 239957283 | | | OpenSea | 0.1358 | 0.0308 | 4.4171 | $1.08\times10^{-05}$ | $9.26\times10^{-02}$ |
| cg09964012 | chr14 | 105036453 | | | OpenSea | 0.1353 | 0.0306 | 4.4151 | $1.09\times10^{-05}$ | $9.26\times10^{-02}$ |
| ch.X.258064R | chrX | 16776121 | SYAP1 | Body | OpenSea | -0.1692 | 0.0385 | -4.391 | $1.22\times10^{-05}$ | $1.01\times10^{-01}$ |
| cg26229895 | chr6 | 30301504 | TRIM39 | Body | OpenSea | -0.1157 | 0.0264 | -4.3817 | $1.27\times10^{-05}$ | $1.04\times10^{-01}$ |
| cg03967192 | chr2 | 10861604 | ATP6V1C2 | TSS200 | Island | 0.1123 | 0.0257 | 4.3692 | $1.35\times10^{-05}$ | $1.08\times10^{-01}$ |
| cg18222759 | chr14 | 22989081 | | | OpenSea | -0.0987 | 0.0227 | -4.3402 | $1.53\times10^{-05}$ | $1.21\times10^{-01}$ |
| cg07389273 | chr10 | 133020009 | TCERG1L | Body | OpenSea | -0.1066 | 0.0247 | -4.3256 | $1.64\times10^{-05}$ | $1.27\times10^{-01}$ |
| cg19780993 | chr14 | 34529242 | | | Island | 0.1267 | 0.0294 | 4.3037 | $1.80\times10^{-05}$ | $1.37\times10^{-01}$ |
| cg12646452 | chr2 | 144560348 | | | OpenSea | -0.0654 | 0.0152 | -4.2983 | $1.85\times10^{-05}$ | $1.38\times10^{-01}$ |
| cg07920148 | chr11 | 33397821 | | | Island | 0.0878 | 0.0205 | 4.2892 | $1.92\times10^{-05}$ | $1.40\times10^{-01}$ |
| cg01691856 | chr15 | 93215561 | | | OpenSea | 0.0879 | 0.0205 | 4.2885 | $1.93\times10^{-05}$ | $1.40\times10^{-01}$ |
| cg03965044 | chr17 | 48712212 | ABCC3 | TSS200 | Island | 0.0761 | 0.0179 | 4.2458 | $2.33\times10^{-05}$ | $1.66\times10^{-01}$ |
| cg18083704 | chr3 | 9851847 | TTLL3 | TSS200 | Island | 0.0759 | 0.0179 | 4.2365 | $2.43\times10^{-05}$ | $1.68\times10^{-01}$ |
| cg09379485 | chr19 | 53758297 | ZNF677 | TSS200 | S_Shore | 0.1417 | 0.0334 | 4.2356 | $2.44\times10^{-05}$ | $1.68\times10^{-01}$ |
| cg13706365 | chr10 | 11504219 | USP6NL | 3'UTR | N_Shore | -0.0968 | 0.023 | -4.2103 | $2.73\times10^{-05}$ | $1.85\times10^{-01}$ |
| cg27514608 | chr4 | 23892540 | PPARGC1A | TSS1500 | OpenSea | -0.0827 | 0.0197 | -4.2055 | $2.78\times10^{-05}$ | $1.86\times10^{-01}$ |
| cg05654340 | chr6 | 111197834 | AMD1 | 5'UTR; Body | S_Shore | -0.1339 | 0.0319 | -4.1953 | $2.91\times10^{-05}$ | $1.92\times10^{-01}$ |
| cg18468511 | chr14 | 27067638 | NOVA1 | TSS1500 | S_Shore | 0.121 | 0.0289 | 4.1807 | $3.10\times10^{-05}$ | $2.02\times10^{-01}$ |
| cg16206511 | chr12 | 62860399 | MON2 | TSS200 | Island | -0.1082 | 0.026 | -4.167 | $3.29\times10^{-05}$ | $2.09\times10^{-01}$ |
| cg12750884 | chr1 | 11303204 | MTOR | Body | OpenSea | -0.0657 | 0.0158 | -4.1651 | $3.32\times10^{-05}$ | $2.09\times10^{-01}$ |
| cg09571420 | chr7 | 145813008 | CNTNAP2 | TSS1500 | N_Shore | 0.0539 | 0.0129 | 4.1632 | $3.34\times10^{-05}$ | $2.09\times10^{-01}$ |

| | | | | | | | | | | |
|---|---|---|---|---|---|---|---|---|---|---|
| cg13971154 | chr7 | 128828460 | SMO | TSS1500 | Island | 0.0538 | 0.013 | 4.1529 | $3.49\times10^{-05}$ | $2.10\times10^{-01}$ |
| cg09357483 | chr16 | 87904491 | SLC7A5 | TSS1500 | S_Shore | 0.3037 | 0.0732 | 4.1474 | $3.58\times10^{-05}$ | $2.10\times10^{-01}$ |
| cg07897077 | chr19 | 56705386 | ZSCAN5B | TSS1500 | N_Shelf | -0.0677 | 0.0163 | -4.147 | $3.58\times10^{-05}$ | $2.10\times10^{-01}$ |
| cg16176837 | chr1 | 5919155 | | | OpenSea | -0.1431 | 0.0345 | -4.1461 | $3.60\times10^{-05}$ | $2.10\times10^{-01}$ |
| cg21773322 | chr8 | 61836533 | | | S_Shore | 0.0633 | 0.0153 | 4.1456 | $3.61\times10^{-05}$ | $2.10\times10^{-01}$ |
| cg07963147 | chr3 | 141086820 | ZBTB38 | 5'UTR | OpenSea | 0.1262 | 0.0305 | 4.1435 | $3.64\times10^{-05}$ | $2.10\times10^{-01}$ |
| cg06228857 | chr15 | 35372947 | | | OpenSea | 0.0769 | 0.0186 | 4.1396 | $3.70\times10^{-05}$ | $2.10\times10^{-01}$ |
| cg20338300 | chr2 | 56411534 | CCDC85A | 1stExon; 5'UTR | Island | 0.0636 | 0.0155 | 4.1133 | $4.14\times10^{-05}$ | $2.30\times10^{-01}$ |
| cg13042252 | chr8 | 74005756 | C8orf84 | TSS1500 | Island | 0.096 | 0.0233 | 4.1131 | $4.15\times10^{-05}$ | $2.30\times10^{-01}$ |
| cg13759808 | chr11 | 64885159 | ZNHIT2 | 1stExon; 5'UTR | Island | 0.0395 | 0.0096 | 4.1078 | $4.24\times10^{-05}$ | $2.30\times10^{-01}$ |
| cg02979703 | chr8 | 37558858 | | | Island | 0.2028 | 0.0494 | 4.1049 | $4.29\times10^{-05}$ | $2.30\times10^{-01}$ |
| cg21625464 | chr11 | 314044 | IFITM1 | 1stExon; 5'UTR | N_Shore | -0.1347 | 0.0328 | -4.104 | $4.31\times10^{-05}$ | $2.30\times10^{-01}$ |
| cg16027727 | chr17 | 78586328 | RPTOR | Body | OpenSea | 0.0862 | 0.021 | 4.0958 | $4.47\times10^{-05}$ | $2.32\times10^{-01}$ |
| cg05672174 | chr5 | 150701709 | SLC36A2 | Body | OpenSea | -0.1312 | 0.032 | -4.095 | $4.48\times10^{-05}$ | $2.32\times10^{-01}$ |
| cg09809720 | chr5 | 162886885 | NUDCD2; HMMR | 1stExon; TSS1500 | Island | -0.0425 | 0.0104 | -4.0947 | $4.49\times10^{-05}$ | $2.32\times10^{-01}$ |
| cg02681842 | chr8 | 145033310 | PLEC1 | Body | OpenSea | 0.0906 | 0.0222 | 4.082 | $4.74\times10^{-05}$ | $2.39\times10^{-01}$ |
| cg05075416 | chr15 | 99500703 | IGF1R | 3'UTR | Island | -0.1804 | 0.0442 | -4.0816 | $4.74\times10^{-05}$ | $2.39\times10^{-01}$ |

Genomic locations are reported according to the hg19 reference genome. Gene annotation and CpG island context correspond to the Illumina HumanMethylation450 array annotation.

**Table S10.** *Top CpG sites showing suggestive evidence of interaction (late childhood trauma)*

| CpG | Chromosome | Position (bp) | Gene(s) | Gene region | CpG island relation | Effect | SE | Statistic | P | FDR |
|---|---|---|---|---|---|---|---|---|---|---|
| cg26320410 | chr3 | 9851826 | TTLL3 | TSS200 | Island | 0.091 | 0.0133 | 6.8678 | $1.01\times10^{-11}$ | $4.52\times10^{-06}$ |
| cg19103000 | chr3 | 9851819 | TTLL3 | TSS200 | Island | 0.0907 | 0.014 | 6.4683 | $1.40\times10^{-10}$ | $3.15\times10^{-05}$ |
| cg09719477 | chr11 | 113930430 | ZBTB16 | 1stExon; TSS1500; 5'UTR | Island | 0.0936 | 0.0156 | 5.9971 | $2.61\times10^{-09}$ | $3.90\times10^{-04}$ |
| cg02936752 | chr19 | 54963326 | LENG8 | Body | S_Shelf | 0.0946 | 0.0163 | 5.7995 | $8.34\times10^{-09}$ | $9.36\times10^{-04}$ |
| cg02882381 | chr11 | 69590332 | FGF4 | TSS200 | Island | 0.0917 | 0.0161 | 5.7025 | $1.46\times10^{-08}$ | $1.16\times10^{-03}$ |
| cg20383084 | chr1 | 26325121 | PAFAH2 | TSS1500 | S_Shore | -0.1619 | 0.0285 | -5.6912 | $1.56\times10^{-08}$ | $1.16\times10^{-03}$ |

| | | | | | | | | | | |
|---|---|---|---|---|---|---|---|---|---|---|
| cg02507480 | chr7 | 112032146 | | | Island | -0.1187 | 0.021 | -5.6515 | $1.97\times10^{-08}$ | $1.27\times10^{-03}$ |
| cg25934863 | chr15 | 101419506 | ALDH1A3 | TSS1500 | Island | 0.1216 | 0.022 | 5.521 | $4.06\times10^{-08}$ | $2.28\times10^{-03}$ |
| cg23239754 | chr6 | 27760147 | | | OpenSea | -0.1012 | 0.0187 | -5.4008 | $7.88\times10^{-08}$ | $3.93\times10^{-03}$ |
| cg26915774 | chr2 | 162094867 | | | Island | 0.1302 | 0.0246 | 5.2851 | $1.47\times10^{-07}$ | $6.61\times10^{-03}$ |
| cg21847751 | chr2 | 232379328 | C2orf52 | TSS1500 | Island | -0.1155 | 0.022 | -5.2559 | $1.72\times10^{-07}$ | $7.03\times10^{-03}$ |
| cg21123573 | chr12 | 120740055 | SIRT4 | TSS200 | OpenSea | -0.2338 | 0.0449 | -5.2012 | $2.31\times10^{-07}$ | $8.64\times10^{-03}$ |
| cg24199112 | chr11 | 69590328 | FGF4 | TSS200 | Island | 0.0869 | 0.0173 | 5.0319 | $5.55\times10^{-07}$ | $1.91\times10^{-02}$ |
| cg10208370 | chr4 | 90758469 | SNCA | TSS1500; 5'UTR; TSS200 | Island | 0.1412 | 0.0281 | 5.0176 | $5.96\times10^{-07}$ | $1.91\times10^{-02}$ |
| cg01357429 | chr7 | 27285563 | EVX1 | Body | Island | 0.0839 | 0.0169 | 4.9557 | $8.16\times10^{-07}$ | $2.44\times10^{-02}$ |
| cg14602982 | chr11 | 119599176 | PVRL1 | Body | Island | 0.0873 | 0.0179 | 4.8875 | $1.15\times10^{-06}$ | $3.12\times10^{-02}$ |
| cg23010344 | chr1 | 228871301 | RHOU | 1stExon; 5'UTR | Island | 0.0861 | 0.0177 | 4.871 | $1.25\times10^{-06}$ | $3.12\times10^{-02}$ |
| cg00530638 | chr5 | 98108543 | RGMB | Body | N_Shore | -0.1961 | 0.0403 | -4.8703 | $1.25\times10^{-06}$ | $3.12\times10^{-02}$ |
| cg20142390 | chr21 | 40564147 | BRWD1 | 3'UTR; Body | OpenSea | -0.0811 | 0.0169 | -4.812 | $1.67\times10^{-06}$ | $3.94\times10^{-02}$ |
| cg05633605 | chr5 | 55530180 | ANKRD55 | TSS1500 | OpenSea | -0.1121 | 0.0234 | -4.7993 | $1.78\times10^{-06}$ | $3.99\times10^{-02}$ |
| cg17572445 | chr13 | 98893146 | FARP1 | Body | OpenSea | -0.172 | 0.0362 | -4.7526 | $2.23\times10^{-06}$ | $4.68\times10^{-02}$ |
| cg02242344 | chr2 | 85640943 | | | N_Shore | 0.1257 | 0.0265 | 4.7437 | $2.34\times10^{-06}$ | $4.68\times10^{-02}$ |
| cg26286805 | chr14 | 51411267 | PYGL | TSS200 | Island | 0.0759 | 0.016 | 4.7376 | $2.40\times10^{-06}$ | $4.68\times10^{-02}$ |
| cg26665891 | chr1 | 204159832 | KISS1 | Body | Island | 0.0883 | 0.0189 | 4.6841 | $3.11\times10^{-06}$ | $5.63\times10^{-02}$ |
| cg18705408 | chr2 | 20212524 | MATN3 | TSS200 | Island | 0.1331 | 0.0284 | 4.6824 | $3.13\times10^{-06}$ | $5.63\times10^{-02}$ |
| cg21875802 | chr2 | 45231382 | | | Island | 0.0958 | 0.0206 | 4.6619 | $3.45\times10^{-06}$ | $5.97\times10^{-02}$ |
| cg05143332 | chr7 | 109218838 | | | OpenSea | -0.1031 | 0.0223 | -4.6227 | $4.16\times10^{-06}$ | $6.92\times10^{-02}$ |
| cg02924619 | chr14 | 65007171 | HSPA2 | TSS200 | Island | -0.0822 | 0.0179 | -4.598 | $4.68\times10^{-06}$ | $7.12\times10^{-02}$ |
| cg20566450 | chr5 | 31854651 | PDZD2 | Body | N_Shore | -0.1433 | 0.0312 | -4.5941 | $4.77\times10^{-06}$ | $7.12\times10^{-02}$ |
| cg04543008 | chr11 | 71955332 | PHOX2A | TSS200 | Island | 0.0964 | 0.021 | 4.5835 | $5.02\times10^{-06}$ | $7.12\times10^{-02}$ |
| cg10080155 | chr22 | 45405904 | PHF21B | TSS1500; TSS200 | Island | 0.0555 | 0.0121 | 4.5806 | $5.08\times10^{-06}$ | $7.12\times10^{-02}$ |
| cg06340552 | chr4 | 142054329 | RNF150 | 1stExon; 5'UTR | Island | 0.0688 | 0.015 | 4.5773 | $5.16\times10^{-06}$ | $7.12\times10^{-02}$ |
| cg22654039 | chr20 | 11871282 | BTBD3 | TSS200 | N_Shore | 0.0777 | 0.017 | 4.5744 | $5.23\times10^{-06}$ | $7.12\times10^{-02}$ |
| cg07112562 | chr10 | 16741619 | RSU1;RSU1 | Body | OpenSea | 0.1066 | 0.0234 | 4.5548 | $5.73\times10^{-06}$ | $7.57\times10^{-02}$ |
| cg18286127 | chr10 | 120070743 | C10orf84 | Body | OpenSea | -0.1124 | 0.0247 | -4.5434 | $6.05\times10^{-06}$ | $7.76\times10^{-02}$ |

| | | | | | | | | | | |
|---|---|---|---|---|---|---|---|---|---|---|
| cg08511716 | chr5 | 94621030 | MCTP1 | TSS1500 | Island | 0.1111 | 0.0246 | 4.5183 | $6.80\times10^{-06}$ | $8.48\times10^{-02}$ |
| cg08324796 | chrX | 48979771 | GPKOW | Body | N_Shore | -0.2265 | 0.0504 | -4.4888 | $7.79\times10^{-06}$ | $9.46\times10^{-02}$ |
| cg15846718 | chr6 | 75953307 | COX7A2 | Body | OpenSea | -0.1428 | 0.0321 | -4.45 | $9.34\times10^{-06}$ | $1.10\times10^{-01}$ |
| cg15654121 | chr1 | 34630944 | CSMD2 | Body | Island | 0.0671 | 0.0151 | 4.4431 | $9.62\times10^{-06}$ | $1.11\times10^{-01}$ |
| cg03933495 | chr16 | 3493614 | NAT15; ZNF597 | TSS200 | S_Shore | -0.1176 | 0.0267 | -4.4111 | $1.11\times10^{-05}$ | $1.22\times10^{-01}$ |
| cg14492800 | chr5 | 37839923 | GDNF | TSS200 | Island | 0.1229 | 0.0279 | 4.4107 | $1.12\times10^{-05}$ | $1.22\times10^{-01}$ |
| cg06695611 | chr2 | 180726328 | ZNF385B; MIR1258 | TSS200; TSS1500 | Island | 0.0784 | 0.0178 | 4.4025 | $1.16\times10^{-05}$ | $1.24\times10^{-01}$ |
| cg22058122 | chr18 | 6414958 | L3MBTL4 | TSS200 | S_Shore | 0.1088 | 0.0248 | 4.3912 | $1.22\times10^{-05}$ | $1.26\times10^{-01}$ |
| cg13118906 | chr17 | 61524292 | CYB561 | TSS1500 | Island | 0.0714 | 0.0163 | 4.3877 | $1.24\times10^{-05}$ | $1.26\times10^{-01}$ |
| cg19591206 | chr3 | 133748809 | SLCO2A1 | 1stExon; 5'UTR | Island | 0.1037 | 0.0237 | 4.3786 | $1.29\times10^{-05}$ | $1.27\times10^{-01}$ |
| cg13324546 | chr8 | 23564031 | NKX2-6 | TSS200 | Island | 0.1406 | 0.0321 | 4.3766 | $1.30\times10^{-05}$ | $1.27\times10^{-01}$ |
| cg09518742 | chr11 | 119020115 | ABCG4 | TSS200; 5'UTR | Island | 0.0509 | 0.0117 | 4.3617 | $1.39\times10^{-05}$ | $1.33\times10^{-01}$ |
| cg15993083 | chr12 | 4381788 | CCND2 | TSS1500 | Island | 0.0796 | 0.0183 | 4.3546 | $1.44\times10^{-05}$ | $1.34\times10^{-01}$ |
| cg10498524 | chr2 | 178937663 | PDE11A | Body; TSS200 | Island | 0.1146 | 0.0265 | 4.3255 | $1.64\times10^{-05}$ | $1.47\times10^{-01}$ |
| cg12252090 | chr8 | 49833279 | SNAI2 | Body | N_Shelf | -0.1496 | 0.0346 | -4.3221 | $1.66\times10^{-05}$ | $1.47\times10^{-01}$ |
| cg01418667 | chr20 | 3387969 | C20orf194 | Body | Island | 0.0696 | 0.0161 | 4.3214 | $1.67\times10^{-05}$ | $1.47\times10^{-01}$ |
| cg15424477 | chr13 | 113428288 | ATP11A | Body | N_Shore | -0.1643 | 0.0381 | -4.3131 | $1.73\times10^{-05}$ | $1.49\times10^{-01}$ |
| cg15087347 | chr12 | 41086227 | CNTN1 | TSS200 | N_Shore | 0.1014 | 0.0236 | 4.301 | $1.83\times10^{-05}$ | $1.55\times10^{-01}$ |
| cg10624122 | chr7 | 19158747 | TWIST1 | TSS1500 | S_Shore | -0.0816 | 0.019 | -4.2888 | $1.93\times10^{-05}$ | $1.58\times10^{-01}$ |
| cg14059988 | chr7 | 127669170 | SND1; LRRC4 | Body | N_Shore | -0.1133 | 0.0264 | -4.2884 | $1.93\times10^{-05}$ | $1.58\times10^{-01}$ |
| cg01116059 | chr5 | 121647630 | SNCAIP | TSS200 | Island | 0.1031 | 0.0241 | 4.2825 | $1.98\times10^{-05}$ | $1.59\times10^{-01}$ |
| cg02022224 | chr17 | 39967428 | SC65 | Body | Island | 0.0543 | 0.0127 | 4.2728 | $2.07\times10^{-05}$ | $1.63\times10^{-01}$ |
| cg06967120 | chr10 | 25464259 | LOC100128811; GPR158 | Body; TSS200 | Island | 0.2031 | 0.0476 | 4.2658 | $2.14\times10^{-05}$ | $1.65\times10^{-01}$ |
| cg03006716 | chr7 | 98741357 | SMURF1 | 1stExon | Island | 0.0505 | 0.0118 | 4.26 | $2.19\times10^{-05}$ | $1.67\times10^{-01}$ |
| cg22871668 | chr6 | 133562492 | EYA4 | TSS200 | Island | 0.1099 | 0.0258 | 4.2516 | $2.27\times10^{-05}$ | $1.70\times10^{-01}$ |
| cg12956507 | chr3 | 193854670 | HES1 | Body | Island | 0.0929 | 0.0219 | 4.2443 | $2.35\times10^{-05}$ | $1.73\times10^{-01}$ |
| cg16674467 | chr3 | 46854311 | | | S_Shore | -0.0936 | 0.0221 | -4.2395 | $2.40\times10^{-05}$ | $1.74\times10^{-01}$ |
| cg20713035 | chr5 | 14410284 | TRIO | Body | OpenSea | -0.0987 | 0.0234 | -4.2207 | $2.60\times10^{-05}$ | $1.83\times10^{-01}$ |
| cg07810961 | chr5 | 139927271 | EIF4EBP3; ANKHD1 | 1stExon; 5'UTR; Body | Island | 0.0645 | 0.0153 | 4.2202 | $2.61\times10^{-05}$ | $1.83\times10^{-01}$ |

| | | | | | | | | | | |
|---|---|---|---|---|---|---|---|---|---|---|
| cg00402812 | chr1 | 109633307 | TMEM167B | TSS200 | Island | -0.0864 | 0.0206 | -4.1971 | $2.89\times10^{-05}$ | $2.00\times10^{-01}$ |
| cg26829088 | chr19 | 3626949 | C19orf29 | TSS200 | Island | -0.0849 | 0.0202 | -4.1912 | $2.96\times10^{-05}$ | $2.02\times10^{-01}$ |
| cg13904562 | chr12 | 48690974 | | | OpenSea | -0.1538 | 0.0367 | -4.1848 | $3.04\times10^{-05}$ | $2.02\times10^{-01}$ |
| cg20467168 | chr1 | 76081408 | | | Island | 0.0657 | 0.0157 | 4.1843 | $3.05\times10^{-05}$ | $2.02\times10^{-01}$ |
| cg27608102 | chr17 | 39680504 | KRT19 | Body | N_Shelf | -0.1934 | 0.0463 | -4.1795 | $3.12\times10^{-05}$ | $2.03\times10^{-01}$ |
| cg04153241 | chr12 | 133562589 | ZNF26 | TSS1500 | N_Shore | -0.2099 | 0.0504 | -4.161 | $3.38\times10^{-05}$ | $2.17\times10^{-01}$ |
| cg26999902 | chr19 | 6108921 | RFX2; RFX2 | 5'UTR | Island | 0.0998 | 0.024 | 4.1566 | $3.44\times10^{-05}$ | $2.18\times10^{-01}$ |
| cg14448830 | chr9 | 79627216 | | | N_Shore | 0.0929 | 0.0224 | 4.1479 | $3.57\times10^{-05}$ | $2.23\times10^{-01}$ |
| cg22979807 | chr19 | 55837197 | TMEM150B | TSS1500 | OpenSea | 0.1457 | 0.0353 | 4.1275 | $3.90\times10^{-05}$ | $2.37\times10^{-01}$ |
| cg15491911 | chr6 | 167764620 | | | Island | 0.0605 | 0.0147 | 4.1256 | $3.93\times10^{-05}$ | $2.37\times10^{-01}$ |
| cg24946736 | chr19 | 7542181 | PEX11G | Body | Island | -0.1245 | 0.0302 | -4.1239 | $3.96\times10^{-05}$ | $2.37\times10^{-01}$ |
| cg06836497 | chr1 | 29063467 | YTHDF2 | 5'UTR; 1stExon | Island | 0.0452 | 0.011 | 4.1187 | $4.05\times10^{-05}$ | $2.39\times10^{-01}$ |
| cg03155112 | chr7 | 47556928 | TNS3 | 5'UTR | OpenSea | -0.1432 | 0.0349 | -4.1019 | $4.37\times10^{-05}$ | $2.44\times10^{-01}$ |
| cg13733797 | chr8 | 121824218 | SNTB1 | 5'UTR; 1stExon | Island | -0.0563 | 0.0137 | -4.0983 | $4.42\times10^{-05}$ | $2.44\times10^{-01}$ |
| cg11722699 | chr18 | 28622813 | DSC3 | TSS200 | Island | 0.1379 | 0.0337 | 4.0959 | $4.47\times10^{-05}$ | $2.44\times10^{-01}$ |
| cg16259904 | chr10 | 134146220 | LRRC27 | 5'UTR | S_Shore | -0.258 | 0.0631 | -4.0923 | $4.53\times10^{-05}$ | $2.44\times10^{-01}$ |
| cg15775218 | chr2 | 3286338 | TSSC1 | Body | Island | -0.2308 | 0.0565 | -4.0854 | $4.67\times10^{-05}$ | $2.44\times10^{-01}$ |
| cg13653153 | chr9 | 130885858 | PTGES2 | Body | N_Shelf | 0.1025 | 0.0251 | 4.0816 | $4.75\times10^{-05}$ | $2.44\times10^{-01}$ |
| cg06664872 | chr5 | 96519548 | RIOK2 | TSS1500 | Island | -0.0853 | 0.0209 | -4.0801 | $4.78\times10^{-05}$ | $2.44\times10^{-01}$ |
| cg13579574 | chr15 | 75975227 | CSPG4 | Body | Island | -0.0761 | 0.0187 | -4.0789 | $4.80\times10^{-05}$ | $2.44\times10^{-01}$ |
| cg13928709 | chr5 | 176237221 | UNC5A | TSS1500 | Island | 0.093 | 0.0228 | 4.0759 | $4.86\times10^{-05}$ | $2.44\times10^{-01}$ |
| cg11664500 | chr6 | 133562479 | EYA4 | TSS200 | Island | 0.0789 | 0.0194 | 4.0752 | $4.88\times10^{-05}$ | $2.44\times10^{-01}$ |
| cg12980477 | chr6 | 27156454 | | | OpenSea | -0.0768 | 0.0189 | -4.0735 | $4.91\times10^{-05}$ | $2.44\times10^{-01}$ |
| cg08189989 | chr2 | 105459164 | | | Island | 0.0642 | 0.0158 | 4.073 | $4.92\times10^{-05}$ | $2.44\times10^{-01}$ |
| cg04754916 | chr19 | 10381498 | ICAM1 | TSS200 | Island | 0.0605 | 0.0148 | 4.0716 | $4.95\times10^{-05}$ | $2.44\times10^{-01}$ |
| cg25815683 | chr6 | 28742918 | | | OpenSea | -0.0828 | 0.0203 | -4.0714 | $4.96\times10^{-05}$ | $2.44\times10^{-01}$ |
| cg00192031 | chr7 | 47620790 | | | N_Shore | -0.0808 | 0.0199 | -4.0701 | $4.98\times10^{-05}$ | $2.44\times10^{-01}$ |

Genomic locations are reported according to the hg19 reference genome. Gene annotation and CpG island context correspond to the Illumina HumanMethylation450 array annotation.

**Table S11.** *Top CpG sites showing suggestive evidence of interaction (abuse-only trauma)*

| CpG | Chromosome | Position (bp) | Gene(s) | Gene region | CpG island relation | Effect | SE | Statistic | P | FDR |
|---|---|---|---|---|---|---|---|---|---|---|
| cg26320410 | chr3 | 9851826 | TTLL3 | TSS200 | Island | 0.0781 | 0.0123 | 6.3334 | $3.28{\times}10^{-10}$ | $1.47{\times}10^{-04}$ |
| cg20383084 | chr1 | 26325121 | PAFAH2 | TSS1500 | S_Shore | -0.1589 | 0.0263 | -6.04 | $2.00{\times}10^{-09}$ | $3.28{\times}10^{-04}$ |
| cg04852989 | chr22 | 19974685 | ARVCF | Body | Island | -0.1317 | 0.0219 | -6.025 | $2.19{\times}10^{-09}$ | $3.28{\times}10^{-04}$ |
| cg02882381 | chr11 | 69590332 | FGF4 | TSS200 | Island | 0.0824 | 0.0149 | 5.5196 | $4.08{\times}10^{-08}$ | $4.58{\times}10^{-03}$ |
| cg24199112 | chr11 | 69590328 | FGF4 | TSS200 | Island | 0.0843 | 0.016 | 5.2575 | $1.70{\times}10^{-07}$ | $1.53{\times}10^{-02}$ |
| cg19103000 | chr3 | 9851819 | TTLL3 | TSS200 | Island | 0.0677 | 0.013 | 5.1943 | $2.38{\times}10^{-07}$ | $1.68{\times}10^{-02}$ |
| cg01357429 | chr7 | 27285563 | EVX1 | Body | Island | 0.081 | 0.0157 | 5.1763 | $2.61{\times}10^{-07}$ | $1.68{\times}10^{-02}$ |
| cg14602982 | chr11 | 119599176 | PVRL1 | Body | Island | 0.0814 | 0.0166 | 4.9077 | $1.04{\times}10^{-06}$ | $5.23{\times}10^{-02}$ |
| cg08197226 | chr15 | 49658975 | C15orf33 | Body | OpenSea | 0.1502 | 0.0306 | 4.9043 | $1.05{\times}10^{-06}$ | $5.23{\times}10^{-02}$ |
| cg10668926 | chr13 | 28527255 | | | N_Shore | 0.0786 | 0.0162 | 4.8673 | $1.27{\times}10^{-06}$ | $5.23{\times}10^{-02}$ |
| cg16315582 | chr11 | 104839349 | CASP4 | TSS200 | OpenSea | -0.1133 | 0.0233 | -4.8649 | $1.28{\times}10^{-06}$ | $5.23{\times}10^{-02}$ |
| cg13118906 | chr17 | 61524292 | CYB561 | TSS1500 | Island | 0.0726 | 0.0151 | 4.8207 | $1.60{\times}10^{-06}$ | $5.74{\times}10^{-02}$ |
| cg08511716 | chr5 | 94621030 | MCTP1 | TSS1500 | Island | 0.1091 | 0.0227 | 4.8125 | $1.66{\times}10^{-06}$ | $5.74{\times}10^{-02}$ |
| cg25245636 | chr17 | 79848873 | ANAPC11 | TSS1500; Body | Island | -0.0619 | 0.0133 | -4.6538 | $3.59{\times}10^{-06}$ | $1.15{\times}10^{-01}$ |
| cg12110327 | chr10 | 38264581 | ZNF25 | 5'UTR | N_Shore | -0.1157 | 0.025 | -4.6326 | $3.97{\times}10^{-06}$ | $1.19{\times}10^{-01}$ |
| cg18705408 | chr2 | 20212524 | MATN3 | TSS200 | Island | 0.1213 | 0.0263 | 4.6037 | $4.55{\times}10^{-06}$ | $1.28{\times}10^{-01}$ |
| cg10008501 | chr8 | 9763558 | | | Island | -0.1674 | 0.0365 | -4.5822 | $5.03{\times}10^{-06}$ | $1.33{\times}10^{-01}$ |
| cg04402633 | chr11 | 2907113 | CDKN1C | TSS200 | N_Shore | 0.0937 | 0.0206 | 4.5517 | $5.81{\times}10^{-06}$ | $1.45{\times}10^{-01}$ |
| cg22892043 | chr11 | 61731930 | BEST1; FTH1 | 3'UTR | N_Shelf | -0.0852 | 0.0188 | -4.524 | $6.61{\times}10^{-06}$ | $1.56{\times}10^{-01}$ |
| cg08298794 | chr2 | 198243396 | | | OpenSea | -0.1671 | 0.0371 | -4.499 | $7.42{\times}10^{-06}$ | $1.61{\times}10^{-01}$ |
| cg20163689 | chr2 | 13088414 | | | OpenSea | -0.1171 | 0.0261 | -4.4877 | $7.83{\times}10^{-06}$ | $1.61{\times}10^{-01}$ |
| cg00789545 | chr14 | 102770909 | RAGE | Body | N_Shore | -0.1138 | 0.0254 | -4.4784 | $8.17{\times}10^{-06}$ | $1.61{\times}10^{-01}$ |
| cg23346227 | chr12 | 57024964 | BAZ2A | Body | S_Shore | 0.1033 | 0.0231 | 4.477 | $8.22{\times}10^{-06}$ | $1.61{\times}10^{-01}$ |
| cg04528961 | chr2 | 32289715 | SPAST | Body | S_Shore | -0.1014 | 0.0228 | -4.4484 | $9.38{\times}10^{-06}$ | $1.73{\times}10^{-01}$ |
| cg00926285 | chr3 | 122398533 | PARP14 | TSS1500 | N_Shore | -0.1461 | 0.0331 | -4.4209 | $1.06{\times}10^{-05}$ | $1.73{\times}10^{-01}$ |
| cg20629468 | chr10 | 81664583 | | | Island | 0.0759 | 0.0172 | 4.4204 | $1.07{\times}10^{-05}$ | $1.73{\times}10^{-01}$ |
| cg18555073 | chr15 | 45002662 | B2M | TSS1500 | N_Shore | -0.1056 | 0.0239 | -4.4189 | $1.07{\times}10^{-05}$ | $1.73{\times}10^{-01}$ |

| | | | | | | | | | | |
|---|---|---|---|---|---|---|---|---|---|---|
| cg18791121 | chr11 | 7694814 | CYB5R2 | 1stExon; 5'UTR | Island | 0.0676 | 0.0153 | 4.4175 | $1.08\times10^{-05}$ | $1.73\times10^{-01}$ |
| cg00261781 | chr2 | 20212517 | MATN3 | TSS200 | Island | 0.1148 | 0.026 | 4.4105 | $1.12\times10^{-05}$ | $1.73\times10^{-01}$ |
| cg01392544 | chr10 | 18429760 | CACNB2 | 1stExon; 5'UTR | Island | 0.0874 | 0.0199 | 4.3979 | $1.18\times10^{-05}$ | $1.75\times10^{-01}$ |
| cg05127380 | chr11 | 63953685 | STIP1 | 1stExon; 5'UTR | Island | -0.0657 | 0.015 | -4.3901 | $1.22\times10^{-05}$ | $1.75\times10^{-01}$ |
| cg03110787 | chr19 | 6217641 | MLLT1 | Body | S_Shelf | -0.1162 | 0.0265 | -4.3862 | $1.24\times10^{-05}$ | $1.75\times10^{-01}$ |
| cg05869491 | chr6 | 209763 | | | OpenSea | -0.1351 | 0.0309 | -4.3737 | $1.32\times10^{-05}$ | $1.75\times10^{-01}$ |
| cg27608102 | chr17 | 39680504 | KRT19 | Body | N_Shelf | -0.1871 | 0.0429 | -4.3665 | $1.36\times10^{-05}$ | $1.75\times10^{-01}$ |
| cg10937890 | chr2 | 170440953 | PPIG | 1stExon; 5'UTR | Island | -0.0583 | 0.0134 | -4.3617 | $1.39\times10^{-05}$ | $1.75\times10^{-01}$ |
| cg18440523 | chr21 | 42690652 | FAM3B | Body | S_Shore | -0.1148 | 0.0263 | -4.3598 | $1.40\times10^{-05}$ | $1.75\times10^{-01}$ |
| cg06527052 | chr1 | 109633875 | TMEM167B | Body | S_Shore | -0.0933 | 0.0215 | -4.3301 | $1.60\times10^{-05}$ | $1.94\times10^{-01}$ |
| cg21306775 | chr18 | 74402010 | | | OpenSea | -0.1386 | 0.0321 | -4.3191 | $1.68\times10^{-05}$ | $1.98\times10^{-01}$ |
| cg08364607 | chr11 | 64677562 | ATG2A | Body | OpenSea | -0.1477 | 0.0342 | -4.314 | $1.72\times10^{-05}$ | $1.98\times10^{-01}$ |
| cg20467168 | chr1 | 76081408 | | | Island | 0.0625 | 0.0146 | 4.2864 | $1.95\times10^{-05}$ | $2.03\times10^{-01}$ |
| cg04700861 | chr10 | 13865934 | FRMD4A | Body | OpenSea | -0.0771 | 0.018 | -4.2848 | $1.96\times10^{-05}$ | $2.03\times10^{-01}$ |
| cg05903046 | chr12 | 50629130 | LIMA1; MIR1293 | Body; TSS1500 | OpenSea | -0.0895 | 0.0209 | -4.2816 | $1.99\times10^{-05}$ | $2.03\times10^{-01}$ |
| cg12657380 | chr1 | 9795590 | CLSTN1 | Body | OpenSea | 0.1443 | 0.0337 | 4.2783 | $2.02\times10^{-05}$ | $2.03\times10^{-01}$ |
| cg25813235 | chr6 | 33397883 | SYNGAP1 | Body | S_Shore | -0.075 | 0.0176 | -4.2561 | $2.23\times10^{-05}$ | $2.03\times10^{-01}$ |
| cg16594913 | chr12 | 5607455 | | | S_Shelf | -0.0972 | 0.0229 | -4.252 | $2.27\times10^{-05}$ | $2.03\times10^{-01}$ |
| cg22654039 | chr20 | 11871282 | BTBD3 | TSS200 | N_Shore | 0.0671 | 0.0158 | 4.2433 | $2.36\times10^{-05}$ | $2.03\times10^{-01}$ |
| cg02507480 | chr7 | 112032146 | | | Island | -0.083 | 0.0196 | -4.2433 | $2.36\times10^{-05}$ | $2.03\times10^{-01}$ |
| cg05735765 | chr18 | 32556655 | MAPRE2 | TSS1500 | Island | -0.1131 | 0.0267 | -4.2421 | $2.37\times10^{-05}$ | $2.03\times10^{-01}$ |
| cg09257635 | chr1 | 228346181 | GJC2 | Body | Island | 0.1192 | 0.0281 | 4.2401 | $2.39\times10^{-05}$ | $2.03\times10^{-01}$ |
| cg27564373 | chrX | 47050093 | UBA1 | TSS200 | Island | -0.1356 | 0.032 | -4.2372 | $2.42\times10^{-05}$ | $2.03\times10^{-01}$ |
| cg04039225 | chr3 | 169629516 | SAMD7 | 5'UTR; 1stExon | OpenSea | -0.0675 | 0.0159 | -4.2339 | $2.45\times10^{-05}$ | $2.03\times10^{-01}$ |
| cg20116266 | chr3 | 127771061 | SEC61A1 | TSS200 | Island | -0.0467 | 0.011 | -4.2312 | $2.48\times10^{-05}$ | $2.03\times10^{-01}$ |
| cg10953002 | chr4 | 74241932 | | | OpenSea | -0.0826 | 0.0195 | -4.2292 | $2.51\times10^{-05}$ | $2.03\times10^{-01}$ |
| cg09586060 | chr11 | 114099086 | ZBTB16 | Body | OpenSea | -0.079 | 0.0187 | -4.2289 | $2.51\times10^{-05}$ | $2.03\times10^{-01}$ |
| cg00004979 | chr12 | 111885466 | SH2B3 | Body | OpenSea | -0.2097 | 0.0497 | -4.2219 | $2.59\times10^{-05}$ | $2.03\times10^{-01}$ |
| cg11351687 | chr6 | 170863423 | PSMB1; TBP | TSS1500; TSS200 | Island | 0.0729 | 0.0173 | 4.2194 | $2.62\times10^{-05}$ | $2.03\times10^{-01}$ |

| | | | | | | | | | | |
|---|---|---|---|---|---|---|---|---|---|---|
| cg09035925 | chr20 | 19915769 | RIN2 | Body | OpenSea | -0.1323 | 0.0314 | -4.2158 | $2.66\times10^{-05}$ | $2.03\times10^{-01}$ |
| cg06081306 | chr17 | 37760830 | NEUROD2 | 3'UTR | N_Shore | -0.0842 | 0.02 | -4.2134 | $2.69\times10^{-05}$ | $2.03\times10^{-01}$ |
| cg23385838 | chr12 | 1755068 | WNT5B | Body | Island | 0.1639 | 0.0389 | 4.2125 | $2.70\times10^{-05}$ | $2.03\times10^{-01}$ |
| cg18426014 | chr12 | 54582795 | SMUG1 | TSS200 | OpenSea | -0.0756 | 0.018 | -4.2114 | $2.71\times10^{-05}$ | $2.03\times10^{-01}$ |
| cg18050233 | chr5 | 42800967 | CCDC152; SEPP1 | 3'UTR; Body | OpenSea | -0.1005 | 0.0239 | -4.2032 | $2.81\times10^{-05}$ | $2.07\times10^{-01}$ |
| cg02460371 | chr1 | 150266604 | MRPS21 | 5'UTR; 1stExon | Island | -0.069 | 0.0165 | -4.1884 | $3.00\times10^{-05}$ | $2.17\times10^{-01}$ |
| cg21412992 | chr1 | 61547497 | NFIA | Body; TSS1500; TSS200 | N_Shore | -0.1265 | 0.0302 | -4.1839 | $3.05\times10^{-05}$ | $2.18\times10^{-01}$ |
| cg06340552 | chr4 | 142054329 | RNF150 | 1stExon; 5'UTR | Island | 0.0583 | 0.0139 | 4.1778 | $3.14\times10^{-05}$ | $2.19\times10^{-01}$ |
| cg00577629 | chr8 | 104426828 | SLC25A32; DCAF13 | Body; TSS200 | N_Shore | -0.0574 | 0.0138 | -4.1705 | $3.24\times10^{-05}$ | $2.19\times10^{-01}$ |
| cg06551997 | chr17 | 33815451 | SLFN12L | Body | S_Shore | -0.1504 | 0.0361 | -4.1673 | $3.28\times10^{-05}$ | $2.19\times10^{-01}$ |
| cg19286500 | chr7 | 29652942 | | | OpenSea | -0.0779 | 0.0187 | -4.1626 | $3.35\times10^{-05}$ | $2.19\times10^{-01}$ |
| cg03819713 | chr6 | 32764987 | | | OpenSea | -0.0811 | 0.0195 | -4.1606 | $3.39\times10^{-05}$ | $2.19\times10^{-01}$ |
| cg21875802 | chr2 | 45231382 | | | Island | 0.0796 | 0.0192 | 4.1559 | $3.45\times10^{-05}$ | $2.19\times10^{-01}$ |
| cg11722699 | chr18 | 28622813 | DSC3 | TSS200 | Island | 0.13 | 0.0313 | 4.154 | $3.48\times10^{-05}$ | $2.19\times10^{-01}$ |
| cg01966091 | chr16 | 69141478 | HAS3 | 5'UTR; 1stExon | Island | 0.1463 | 0.0352 | 4.1539 | $3.48\times10^{-05}$ | $2.19\times10^{-01}$ |
| cg08486065 | chr19 | 3464875 | | | Island | 0.0831 | 0.02 | 4.1516 | $3.51\times10^{-05}$ | $2.19\times10^{-01}$ |
| cg06834431 | chr12 | 45049153 | NELL2 | Body | OpenSea | -0.1311 | 0.0317 | -4.142 | $3.66\times10^{-05}$ | $2.25\times10^{-01}$ |
| cg21174055 | chr9 | 139097029 | LHX3 | TSS200 | S_Shore | -0.103 | 0.025 | -4.125 | $3.94\times10^{-05}$ | $2.39\times10^{-01}$ |
| cg23632875 | chr10 | 133110244 | TCERG1L | TSS1500 | Island | 0.0858 | 0.0208 | 4.1165 | $4.08\times10^{-05}$ | $2.44\times10^{-01}$ |
| cg08791424 | chr2 | 208795653 | PLEKHM3 | Body | Island | -0.1874 | 0.0457 | -4.0992 | $4.40\times10^{-05}$ | $2.60\times10^{-01}$ |
| cg09512575 | chr5 | 107008490 | | | S_Shore | -0.1027 | 0.0251 | -4.0961 | $4.46\times10^{-05}$ | $2.60\times10^{-01}$ |
| cg21847751 | chr2 | 232379328 | C2orf52 | TSS1500 | Island | -0.0834 | 0.0204 | -4.0931 | $4.52\times10^{-05}$ | $2.60\times10^{-01}$ |
| cg01365762 | chr6 | 30457827 | HLA-E | Body | Island | 0.1284 | 0.0314 | 4.0897 | $4.58\times10^{-05}$ | $2.60\times10^{-01}$ |
| cg15599382 | chr1 | 244084377 | | | S_Shelf | -0.0958 | 0.0235 | -4.0833 | $4.71\times10^{-05}$ | $2.64\times10^{-01}$ |
| cg21833640 | chr8 | 124287665 | ZHX1 | TSS1500 | Island | -0.0559 | 0.0137 | -4.0784 | $4.81\times10^{-05}$ | $2.66\times10^{-01}$ |
| cg01883966 | chr6 | 31938886 | STK19; DOM3Z | TSS200; TSS1500; Body | N_Shore | -0.1263 | 0.031 | -4.0748 | $4.88\times10^{-05}$ | $2.67\times10^{-01}$ |

Genomic locations are reported according to the hg19 reference genome. Gene annotation and CpG island context correspond to the Illumina HumanMethylation450 array annotation.

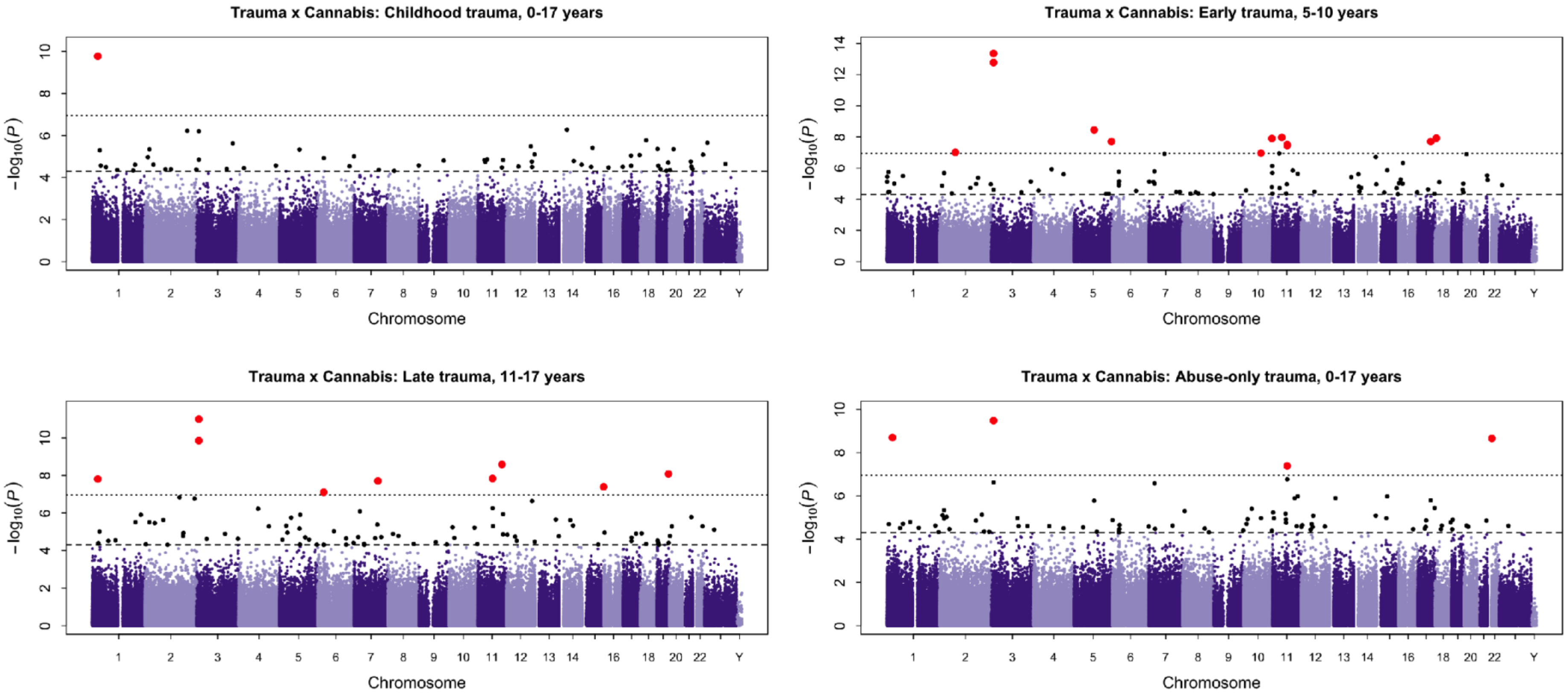


**Figure S9.** *Epigenome-wide interaction analysis of childhood trauma and cannabis.*

Manhattan plot showing epigenome-wide interaction results examining trauma x cannabis effects on DNAm. Each point represents a CpG site plotted according to genomic position across chromosomes and the $-\log_{10}$-transformed p-value. The horizontal dashed line indicates a suggestive discovery threshold ($p < 5 \times 10^{-5}$), while the dotted line represents the Bonferroni-corrected threshold.

#### *7.3. DMR interaction results*

**Table S12.** *Differentially methylated regions identified in trauma x cannabis interaction analyses*

| Exposure | Chromosome | Start (bp) | End (bp) | Region width (bp) | CpG sites | Gene(s) | Min CpG p-value | Region p-value | FDR (BH) |
|---|---|---|---|---|---|---|---|---|---|
| *Abuse-only trauma, 0-17 years* | chr3 | 196325238 | 196325552 | 315 | 3 | | $1.6600\times10^{-4}$ | $2.0000\times10^{-14}$ | $9.7000\times10^{-9}$ |
| *Abuse-only trauma, 0-17 years* | chr11 | 63997961 | 63998032 | 72 | 3 | DNAJC4 | $6.9900\times10^{-3}$ | $5.3400\times10^{-13}$ | $2.5900\times10^{-7}$ |
| *Abuse-only trauma, 0-17 years* | chr6 | 26196580 | 26197071 | 492 | 4 | HIST1H3D | $7.0700\times10^{-5}$ | $3.3200\times10^{-12}$ | $1.6100\times10^{-6}$ |
| *Abuse-only trauma, 0-17 years* | chrX | 118986947 | 118987202 | 256 | 9 | UPF3B | $1.8500\times10^{-3}$ | $3.6200\times10^{-12}$ | $1.7600\times10^{-6}$ |
| *Abuse-only trauma, 0-17 years* | chr6 | 110736772 | 110737053 | 282 | 5 | DDO | $9.2300\times10^{-3}$ | $2.1300\times10^{-11}$ | $1.0300\times10^{-5}$ |
| *Abuse-only trauma, 0-17 years* | chr6 | 31275643 | 31275807 | 165 | 8 | | $3.7800\times10^{-3}$ | $4.3800\times10^{-9}$ | $2.1300\times10^{-3}$ |
| *Abuse-only trauma, 0-17 years* | chr5 | 94621030 | 94621055 | 26 | 2 | MCTP1 | $1.6600\times10^{-6}$ | $5.3700\times10^{-9}$ | $2.6100\times10^{-3}$ |
| *Abuse-only trauma, 0-17 years* | chr11 | 69590328 | 69590332 | 5 | 2 | FGF4 | $4.0800\times10^{-8}$ | $9.9500\times10^{-9}$ | $4.8300\times10^{-3}$ |
| *Abuse-only trauma, 0-17 years* | chr15 | 49658865 | 49658975 | 111 | 2 | C15orf33 | $1.0500\times10^{-6}$ | $1.0200\times10^{-8}$ | $4.9500\times10^{-3}$ |
| *Abuse-only trauma, 0-17 years* | chr8 | 125313885 | 125313940 | 56 | 2 | | $5.0100\times10^{-3}$ | $2.0800\times10^{-8}$ | 0.0101 |
| *Abuse-only trauma, 0-17 years* | chr11 | 2292905 | 2293201 | 297 | 8 | ASCL2 | $6.8500\times10^{-5}$ | $2.6100\times10^{-8}$ | 0.0127 |
| *Abuse-only trauma, 0-17 years* | chrX | 56590097 | 56590113 | 17 | 2 | UBQLN2 | $1.3900\times10^{-3}$ | $3.6400\times10^{-8}$ | 0.0177 |
| *Abuse-only trauma, 0-17 years* | chr2 | 73052964 | 73053217 | 254 | 2 | EXOC6B | $4.4800\times10^{-3}$ | $4.0500\times10^{-8}$ | 0.0197 |
| *Abuse-only trauma, 0-17 years* | chr1 | 36108206 | 36108212 | 7 | 2 | PSMB2 | $2.0900\times10^{-4}$ | $4.4700\times10^{-8}$ | 0.0217 |
| *Abuse-only trauma, 0-17 years* | chr11 | 20408613 | 20408972 | 360 | 5 | PRMT3 | 0.0179 | $5.7200\times10^{-8}$ | 0.0278 |
| *Early trauma, 5-10 years* | chrX | 46405000 | 46405123 | 124 | 4 | ZNF674; LOC401588 | 0.0154 | $1.0600\times10^{-17}$ | $5.1300\times10^{-12}$ |
| *Early trauma, 5-10 years* | chr6 | 31275643 | 31275807 | 165 | 8 | | $1.2400\times10^{-4}$ | $1.4600\times10^{-15}$ | $7.0600\times10^{-10}$ |
| *Early trauma, 5-10 years* | chr3 | 9851819 | 9851847 | 29 | 3 | TTLL3 | $4.3500\times10^{-14}$ | $4.5600\times10^{-15}$ | $2.2000\times10^{-9}$ |
| *Early trauma, 5-10 years* | chr6 | 29720530 | 29720720 | 191 | 13 | | $1.3400\times10^{-3}$ | $3.6300\times10^{-11}$ | $1.7500\times10^{-5}$ |
| *Early trauma, 5-10 years* | chr5 | 94621030 | 94621055 | 26 | 2 | MCTP1 | $3.5400\times10^{-9}$ | $1.6500\times10^{-10}$ | $7.9900\times10^{-5}$ |
| *Early trauma, 5-10 years* | chr19 | 51226046 | 51226849 | 804 | 6 | CLEC11A | $6.0300\times10^{-3}$ | $2.5400\times10^{-10}$ | $1.2300\times10^{-4}$ |
| *Early trauma, 5-10 years* | chrX | 125687011 | 125687207 | 197 | 4 | DCAF12L1 | $1.2300\times10^{-3}$ | $8.3900\times10^{-10}$ | $4.0500\times10^{-4}$ |
| *Early trauma, 5-10 years* | chr8 | 42234614 | 42234803 | 190 | 6 | DKK4 | $9.8300\times10^{-4}$ | $1.3200\times10^{-9}$ | $6.3900\times10^{-4}$ |
| *Early trauma, 5-10 years* | chr5 | 140579296 | 140579644 | 349 | 5 | PCDHB11 | $5.6400\times10^{-4}$ | $1.7300\times10^{-9}$ | $8.3700\times10^{-4}$ |
| *Early trauma, 5-10 years* | chr11 | 69590328 | 69590332 | 5 | 2 | FGF4 | $3.0700\times10^{-8}$ | $3.6800\times10^{-9}$ | $1.7800\times10^{-3}$ |

| | | | | | | | | | |
|---|---|---|---|---|---|---|---|---|---|
| *Early trauma, 5-10 years* | chr15 | 66679100 | 66679417 | 318 | 4 | MAP2K1 | $8.8600\times10^{-3}$ | $5.0700\times10^{-9}$ | $2.4500\times10^{-3}$ |
| *Early trauma, 5-10 years* | chr4 | 10459929 | 10460009 | 81 | 2 | ZNF518B | $2.6100\times10^{-3}$ | $2.0500\times10^{-8}$ | $9.9100\times10^{-3}$ |
| *Early trauma, 5-10 years* | chr21 | 38362727 | 38362754 | 28 | 3 | HLCS | $3.2400\times10^{-3}$ | $2.8500\times10^{-8}$ | 0.0138 |
| *Early trauma, 5-10 years* | chr19 | 58258135 | 58258646 | 512 | 4 | ZNF776 | $3.9200\times10^{-4}$ | $2.9700\times10^{-8}$ | 0.0143 |
| *Early trauma, 5-10 years* | chr20 | 11871282 | 11871311 | 30 | 2 | BTBD3 | $1.2800\times10^{-7}$ | $3.3400\times10^{-8}$ | 0.0161 |
| *Early trauma, 5-10 years* | chr2 | 31806234 | 31806781 | 548 | 7 | SRD5A2 | $8.1500\times10^{-3}$ | $3.9300\times10^{-8}$ | 0.0190 |
| *Early trauma, 5-10 years* | chr2 | 20212517 | 20212524 | 8 | 2 | MATN3 | $2.0400\times10^{-6}$ | $4.6600\times10^{-8}$ | 0.0225 |
| *Early trauma, 5-10 years* | chr8 | 145106299 | 145106438 | 140 | 2 | OPLAH | $7.1700\times10^{-4}$ | $6.9300\times10^{-8}$ | 0.0335 |
| *Early trauma, 5-10 years* | chr2 | 180726252 | 180726328 | 77 | 2 | ZNF385B; MIR1258 | $4.1700\times10^{-6}$ | $7.7900\times10^{-8}$ | 0.0377 |
| *Late trauma, 11-17 years* | chr12 | 70636487 | 70636659 | 173 | 8 | CNOT2 | 0.0282 | $2.3800\times10^{-14}$ | $1.1500\times10^{-8}$ |
| *Late trauma, 11-17 years* | chr5 | 98108198 | 98108543 | 346 | 2 | RGMB | $1.2500\times10^{-6}$ | $3.1600\times10^{-14}$ | $1.5300\times10^{-8}$ |
| *Late trauma, 11-17 years* | chr3 | 9851819 | 9851826 | 8 | 2 | TTLL3 | $1.0100\times10^{-11}$ | $3.4500\times10^{-12}$ | $1.6700\times10^{-6}$ |
| *Late trauma, 11-17 years* | chr2 | 54785178 | 54785406 | 229 | 5 | SPTBN1 | $8.8000\times10^{-3}$ | $1.0500\times10^{-10}$ | $5.0900\times10^{-5}$ |
| *Late trauma, 11-17 years* | chr4 | 90758469 | 90758494 | 26 | 2 | SNCA | $5.9600\times10^{-7}$ | $6.7300\times10^{-10}$ | $3.2500\times10^{-4}$ |
| *Late trauma, 11-17 years* | chr8 | 144373356 | 144373369 | 14 | 3 | ZNF696 | $3.1300\times10^{-3}$ | $2.3700\times10^{-9}$ | $1.1400\times10^{-3}$ |
| *Late trauma, 11-17 years* | chr11 | 69590328 | 69590332 | 5 | 2 | FGF4 | $1.4600\times10^{-8}$ | $1.0000\times10^{-8}$ | $4.8300\times10^{-3}$ |
| *Late trauma, 11-17 years* | chr19 | 54963304 | 54963326 | 23 | 3 | LENG8 | $8.3400\times10^{-9}$ | $1.2100\times10^{-8}$ | $5.8700\times10^{-3}$ |
| *Late trauma, 11-17 years* | chr1 | 26233376 | 26233623 | 248 | 8 | STMN1 | $5.2600\times10^{-3}$ | $1.3300\times10^{-8}$ | $6.4500\times10^{-3}$ |
| *Late trauma, 11-17 years* | chr2 | 240171712 | 240171748 | 37 | 2 | HDAC4 | $5.3800\times10^{-3}$ | $2.1000\times10^{-8}$ | 0.0101 |
| *Late trauma, 11-17 years* | chr6 | 31275664 | 31276146 | 483 | 12 | | 0.0145 | $2.3000\times10^{-8}$ | 0.0111 |
| *Late trauma, 11-17 years* | chr13 | 47472200 | 47472349 | 150 | 5 | HTR2A | $3.3800\times10^{-4}$ | $2.7400\times10^{-8}$ | 0.0132 |
| *Late trauma, 11-17 years* | chr6 | 26196580 | 26196794 | 215 | 3 | | $3.9500\times10^{-4}$ | $3.7600\times10^{-8}$ | 0.0182 |
| *Late trauma, 11-17 years* | chr6 | 32064656 | 32064956 | 301 | 14 | TNXB | 0.0107 | $4.1100\times10^{-8}$ | 0.0199 |
| *Late trauma, 11-17 years* | chr22 | 24891129 | 24891158 | 30 | 4 | C22orf45; UPB1 | 0.0134 | $5.2400\times10^{-8}$ | 0.0253 |
| *Late trauma, 11-17 years* | chr12 | 49297508 | 49297846 | 339 | 6 | CCDC65 | $4.7700\times10^{-3}$ | $6.1500\times10^{-8}$ | 0.0297 |
| *Late trauma, 11-17 years* | chr5 | 94621030 | 94621055 | 26 | 2 | MCTP1 | $6.8000\times10^{-6}$ | $7.3700\times10^{-8}$ | 0.0356 |
| *Childhood trauma, 0-17 years* | chr6 | 31275267 | 31276437 | 1171 | 20 | | $2.6400\times10^{-3}$ | $2.6900\times10^{-13}$ | $1.3000\times10^{-7}$ |
| *Childhood trauma, 0-17 years* | chr4 | 10459929 | 10460237 | 309 | 3 | ZNF518B | $6.4900\times10^{-4}$ | $4.8600\times10^{-12}$ | $2.3500\times10^{-6}$ |
| *Childhood trauma, 0-17 years* | chr1 | 36108212 | 36108225 | 14 | 2 | PSMB2 | $5.0800\times10^{-6}$ | $2.9000\times10^{-11}$ | $1.4000\times10^{-5}$ |

| | | | | | | | | | |
|---|---|---|---|---|---|---|---|---|---|
| *Childhood trauma, 0-17 years* | chr6 | 110736772 | 110737053 | 282 | 5 | DDO | $4.9400\times10^{-3}$ | $3.5800\times10^{-11}$ | $1.7300\times10^{-5}$ |
| *Childhood trauma, 0-17 years* | chrX | 118986954 | 118987202 | 249 | 8 | UPF3B | $2.3400\times10^{-3}$ | $5.3000\times10^{-11}$ | $2.5600\times10^{-5}$ |
| *Childhood trauma, 0-17 years* | chr7 | 94023308 | 94023557 | 250 | 3 | COL1A2 | $9.6900\times10^{-3}$ | $1.1600\times10^{-10}$ | $5.6100\times10^{-5}$ |
| *Childhood trauma, 0-17 years* | chrX | 153744384 | 153744612 | 229 | 5 | FAM3A | 0.0178 | $3.5100\times10^{-10}$ | $1.7000\times10^{-4}$ |
| *Childhood trauma, 0-17 years* | chr2 | 240171712 | 240171748 | 37 | 2 | HDAC4 | $2.7600\times10^{-3}$ | $2.1700\times10^{-9}$ | $1.0500\times10^{-3}$ |
| *Childhood trauma, 0-17 years* | chr2 | 31806234 | 31806767 | 534 | 6 | SRD5A2 | $9.0900\times10^{-3}$ | $5.1700\times10^{-9}$ | $2.5000\times10^{-3}$ |
| *Childhood trauma, 0-17 years* | chr8 | 145106299 | 145106438 | 140 | 2 | OPLAH | $2.6700\times10^{-5}$ | $7.9200\times10^{-9}$ | $3.8200\times10^{-3}$ |
| *Childhood trauma, 0-17 years* | chr5 | 140579296 | 140579644 | 349 | 5 | PCDHB11 | $7.7000\times10^{-5}$ | $1.4400\times10^{-8}$ | $6.9700\times10^{-3}$ |
| *Childhood trauma, 0-17 years* | chr1 | 26233404 | 26233623 | 220 | 7 | STMN1 | $9.8800\times10^{-4}$ | $1.6900\times10^{-8}$ | $8.1900\times10^{-3}$ |
| *Childhood trauma, 0-17 years* | chrX | 49687068 | 49687084 | 17 | 3 | CLCN5 | $8.6800\times10^{-4}$ | $2.1000\times10^{-8}$ | 0.0102 |
| *Childhood trauma, 0-17 years* | chrX | 151999309 | 151999547 | 239 | 5 | NSDHL; CETN2 | 0.0101 | $2.2800\times10^{-8}$ | 0.0110 |
| *Childhood trauma, 0-17 years* | chr15 | 49658865 | 49658975 | 111 | 2 | C15orf33 | $3.8400\times10^{-6}$ | $5.5300\times10^{-8}$ | 0.0267 |
| *Childhood trauma, 0-17 years* | chr2 | 208988863 | 208988967 | 105 | 3 | CRYGD | $5.6400\times10^{-3}$ | $6.1200\times10^{-8}$ | 0.0296 |
| *Childhood trauma, 0-17 years* | chr3 | 196325238 | 196325552 | 315 | 3 | | $4.6600\times10^{-3}$ | $8.4400\times10^{-8}$ | 0.0408 |

*7.4. Functional enrichment*

**Table S13.** *Gene Ontology (GO) and KEGG pathway enrichment results for genes annotated to DMRs*

| Exposure | Database | Ontology | Description | N | DE | P.DE | FDR |
|---|---|---|---|---|---|---|---|
| **Abuse-only trauma, 0-17 years** | GO | BP | negative regulation of endocytosis | 56 | 2 | 5.85E-04 | 1 |
| **Abuse-only trauma, 0-17 years** | GO | BP | somatic stem cell population maintenance | 68 | 2 | 0.00107 | 1 |
| **Abuse-only trauma, 0-17 years** | GO | BP | stem cell population maintenance | 177 | 2 | 0.00712 | 1 |
| **Abuse-only trauma, 0-17 years** | GO | BP | maintenance of cell number | 181 | 2 | 0.00745 | 1 |
| **Abuse-only trauma, 0-17 years** | GO | BP | hormone metabolic process | 240 | 2 | 0.00818 | 1 |
| **Abuse-only trauma, 0-17 years** | GO | BP | regulation of endocytosis | 213 | 2 | 0.0087 | 1 |
| **Abuse-only trauma, 0-17 years** | GO | BP | cellular macromolecule catabolic process | 1018 | 3 | 0.0301 | 1 |
| **Abuse-only trauma, 0-17 years** | GO | BP | negative regulation of transport | 478 | 2 | 0.0328 | 1 |
| **Abuse-only trauma, 0-17 years** | KEGG | KEGG | Amyotrophic lateral sclerosis | 347 | 2 | 0.0225 | 1 |
| **Early childhood trauma, 5-10 years** | GO | MF | ligase activity, forming carbon-nitrogen bonds | 48 | 2 | 0.00124 | 1 |
| **Early childhood trauma, 5-10 years** | GO | MF | signalling receptor regulator activity | 515 | 3 | 0.00737 | 1 |
| **Early childhood trauma, 5-10 years** | GO | MF | growth factor activity | 159 | 2 | 0.00819 | 1 |
| **Early childhood trauma, 5-10 years** | GO | MF | ion binding | 5867 | 11 | 0.0113 | 1 |
| **Early childhood trauma, 5-10 years** | GO | MF | ligase activity | 160 | 2 | 0.0114 | 1 |
| **Early childhood trauma, 5-10 years** | GO | BP | pallium development | 181 | 2 | 0.0145 | 1 |
| **Early childhood trauma, 5-10 years** | GO | BP | positive regulation of ERK1 and ERK2 cascade | 219 | 2 | 0.0153 | 1 |
| **Early childhood trauma, 5-10 years** | GO | BP | skeletal system development | 533 | 3 | 0.0155 | 1 |
| **Early childhood trauma, 5-10 years** | KEGG | KEGG | Melanoma | 72 | 2 | 0.00232 | 0.452 |
| **Early childhood trauma, 5-10 years** | KEGG | KEGG | Prostate cancer | 105 | 2 | 0.00505 | 0.623 |
| **Early childhood trauma, 5-10 years** | KEGG | KEGG | Gastric cancer | 149 | 2 | 0.00977 | 0.701 |
| **Early childhood trauma, 5-10 years** | KEGG | KEGG | Breast cancer | 147 | 2 | 0.00978 | 0.701 |
| **Adolescent trauma, 11-17 years** | GO | BP | negative regulation of thrombin-activated receptor signalling pathway | 3 | 2 | 8E-06 | 0.0916 |

| | | | | | | | |
|---|---|---|---|---|---|---|---|
| **Adolescent trauma, 11-17 years** | GO | BP | regulation of thrombin-activated receptor signalling pathway | 3 | 2 | 8E-06 | 0.0916 |
| **Adolescent trauma, 11-17 years** | GO | BP | regulation of purine nucleotide metabolic process | 90 | 3 | 7.99E-05 | 0.363 |
| **Adolescent trauma, 11-17 years** | GO | BP | regulation of nucleotide metabolic process | 91 | 3 | 8.09E-05 | 0.363 |
| **Adolescent trauma, 11-17 years** | GO | BP | negative regulation of protein polymerization | 78 | 3 | 8.74E-05 | 0.363 |
| **Adolescent trauma, 11-17 years** | GO | BP | thrombin-activated receptor signalling pathway | 13 | 2 | 9.76E-05 | 0.363 |
| **Adolescent trauma, 11-17 years** | GO | BP | negative regulation of microtubule polymerization | 13 | 2 | 1.11E-04 | 0.363 |
| **Adolescent trauma, 11-17 years** | GO | BP | regulation of generation of precursor metabolites and energy | 132 | 3 | 2.76E-04 | 0.79 |
| **Adolescent trauma, 11-17 years** | KEGG | KEGG | MicroRNAs in cancer | 305 | 3 | 0.00164 | 0.606 |
| **Adolescent trauma, 11-17 years** | KEGG | KEGG | Calcium signalling pathway | 250 | 2 | 0.0355 | 1 |
| **Adolescent trauma, 11-17 years** | KEGG | KEGG | MAPK signalling pathway | 298 | 2 | 0.0492 | 1 |
| **Childhood trauma, 0-17 years** | GO | MF | steroid dehydrogenase activity, acting on the CH-OH group of donors, NAD or NADP as acceptor | 30 | 2 | 1.43E-04 | 1 |
| **Childhood trauma, 0-17 years** | GO | MF | steroid dehydrogenase activity | 34 | 2 | 1.88E-04 | 1 |
| **Childhood trauma, 0-17 years** | GO | BP | skin development | 304 | 3 | 0.00188 | 1 |
| **Childhood trauma, 0-17 years** | GO | MF | oxidoreductase activity, acting on the CH-OH group of donors, NAD or NADP as acceptor | 122 | 2 | 0.00403 | 1 |
| **Childhood trauma, 0-17 years** | GO | MF | oxidoreductase activity, acting on CH-OH group of donors | 132 | 2 | 0.00454 | 1 |
| **Childhood trauma, 0-17 years** | GO | MF | hydrolase activity, acting on carbon-nitrogen (but not peptide) bonds | 131 | 2 | 0.00517 | 1 |
| **Childhood trauma, 0-17 years** | GO | BP | mRNA transport | 127 | 2 | 0.00592 | 1 |
| **Childhood trauma, 0-17 years** | GO | BP | skin epidermis development | 121 | 2 | 0.00618 | 1 |
| **Childhood trauma, 0-17 years** | KEGG | KEGG | Neutrophil extracellular trap formation | 175 | 2 | 0.00673 | 1 |
| **Childhood trauma, 0-17 years** | KEGG | KEGG | MicroRNAs in cancer | 305 | 2 | 0.0151 | 1 |
| **Childhood trauma, 0-17 years** | KEGG | KEGG | Metabolic pathways | 1531 | 4 | 0.0382 | 1 |

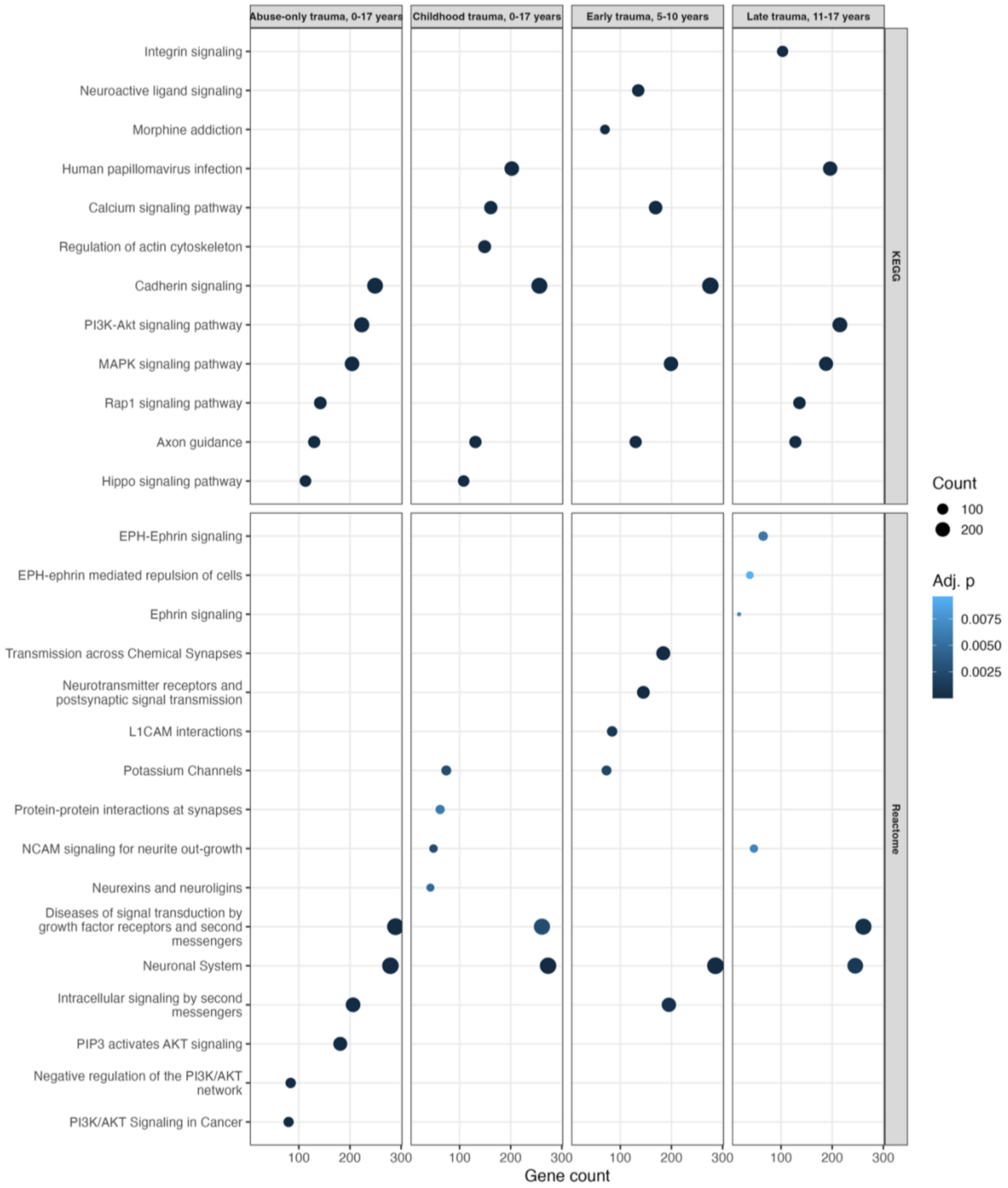


**Figure S10.** *Functional enrichment of cannabis-moderated trauma-DMRs.*
Dot plots showing the top enriched KEGG and Reactome pathways for genes mapped to DMRs identified in childhood trauma x cannabis interaction models across the four trauma definitions. Dot sizes represent gene count and colour indicates adjusted p-value.

#### *8. Mediation Analyses (DACT)*

Epigenome-wide mediation analyses were conducted using the Divide-Aggregate Composite-null Test (DACT) framework to evaluate whether DNAm mediates the association between childhood trauma and PLEs. Diagnostic plots and genome-wide summaries of DACT results are presented below. These analyses are exploratory and interpreted cautiously.

##### *8.1. DACT Model Diagnostics*

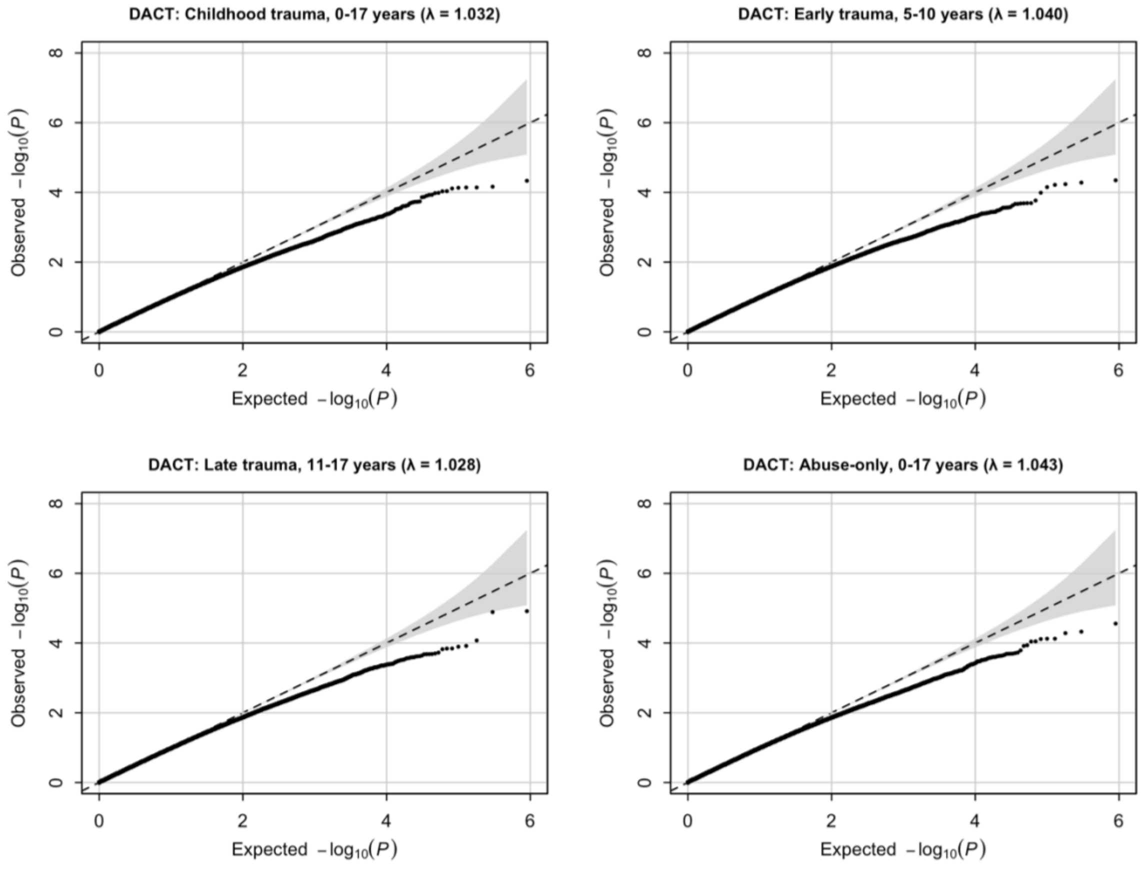


**Figure S11.** *QQ plots for EWAS analyses using the DACT framework.*
The dashed line indicates the null expectation and the shaded region the 95% confidence envelope. Genomic inflation factors (λ) are reported in each panel.

#### *8.2. CpG-level mediation results*

CpG-level mediation results from the DACT framework are summarised using Manhattan and quadrant plots (**Figures S12-S13**).

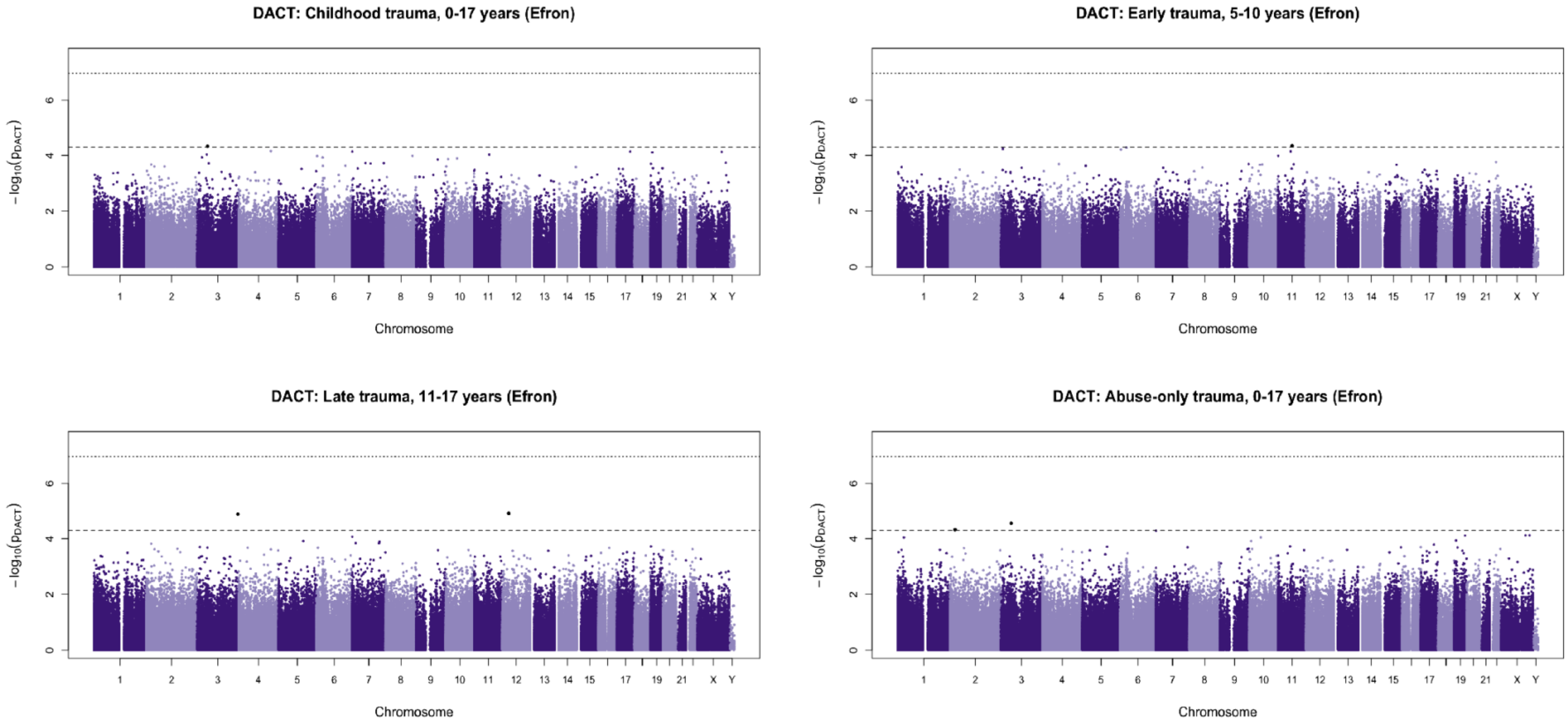


**Figure S12.** *Epigenome-wide mediation analysis of early childhood trauma and PLEs (DACT).*
Manhattan plot showing epigenome-wide mediation results from the DACT analysis examining DNAm as a mediator of the association between childhood trauma and PLEs at age 18. Each point represents a CpG site plotted according to genomic position across chromosomes and the -$\log_{10}$-transformed DACT p-value. The horizontal dashed line indicates a suggestive discovery threshold ($p < 5 \times 10^{-5}$), while the dotted line represents the Bonferroni-corrected threshold.

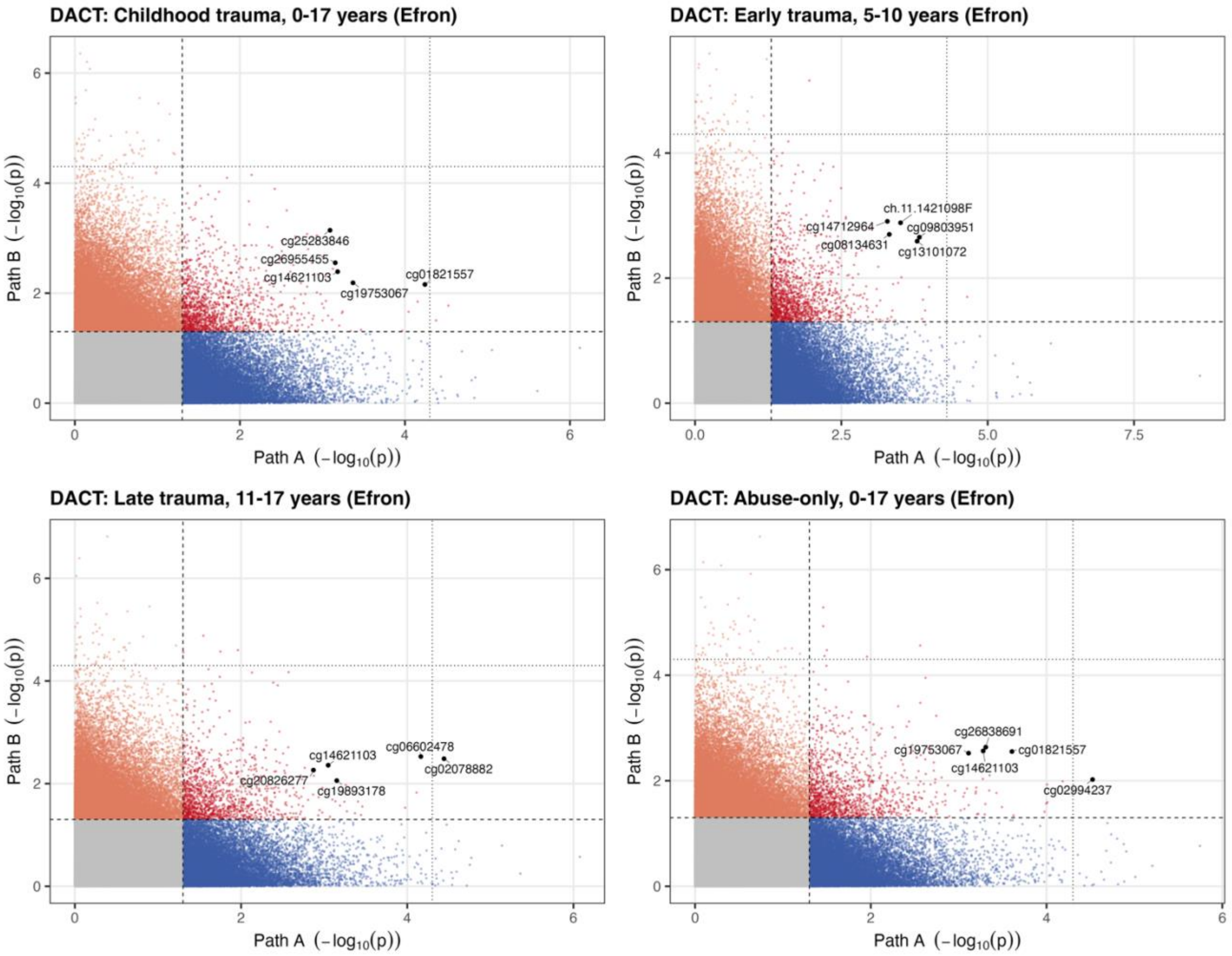


**Figure S13.** *Quadrant plots of CpG-level associations for Path A and Path B.*

Each point represents a CpG site plotted according to the -$\log_{10}$-transformed p-values from the Path A model and the Path B model. Dashed lines indicate the nominal significance thresholds (p = 0.05). points are coloured according to significance pattern: both models, path A only, path B only, or neither. CpG sites showing suggestive mediation signal are highlighted and labelled.

*8.3. Cannabis-moderated Mediation (DACT)*

Exploratory mediation analyses were conducted using the DACT framework to assess whether DNAm mediated the association between childhood trauma and PLEs under cannabis-moderated conditions. Across models, there was little evidence supporting a mediating role of DNAm after correction for multiple testing (**Table S14**). While a small number of CpG sites showed nominal significance, these did not survive FDR correction and should be interpreted cautiously. Overall, these findings are consistent with the primary analyses, providing limited support for DNAm as a mediator of the trauma-PLE association, even when accounting for potential modification by cannabis use.

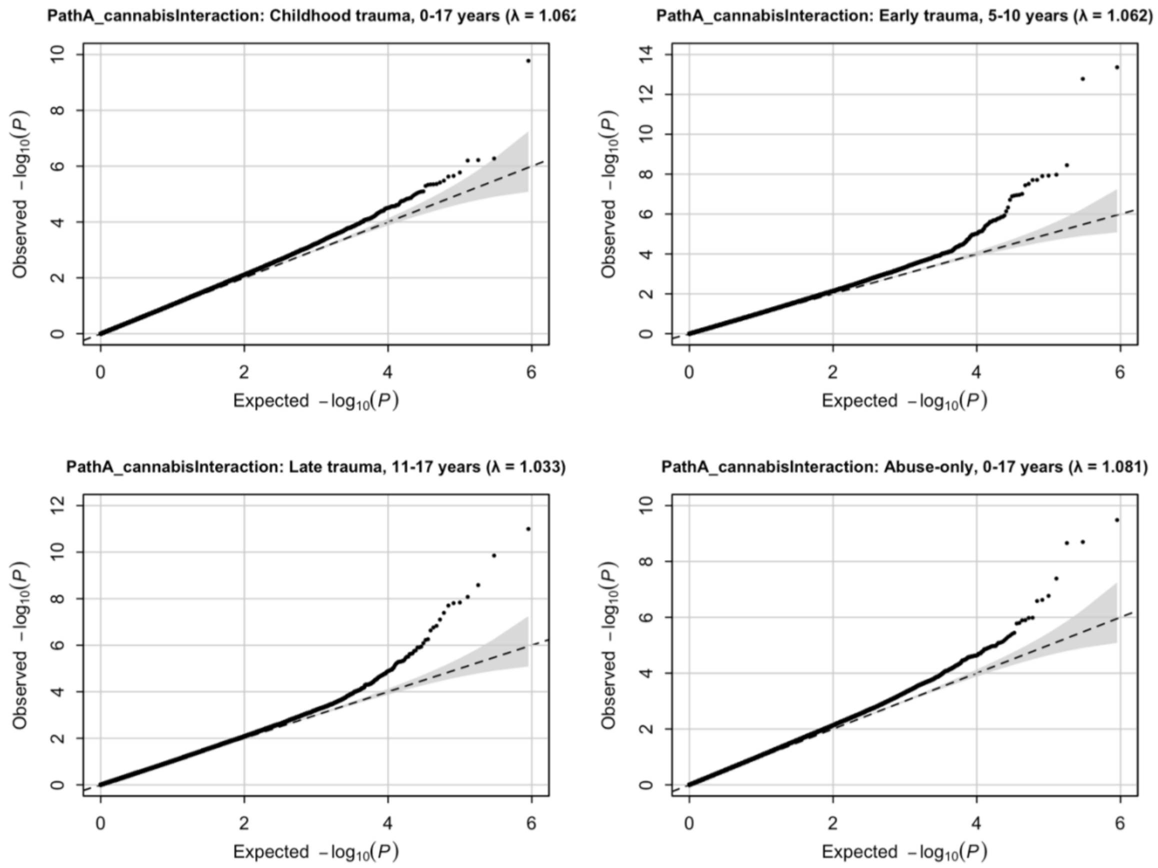


**Figure S14.** *QQ plots for EWAS-moderated analyses using the DACT framework.*
The dashed line indicates the null expectation and the shaded region the 95% confidence envelope. Genomic inflation factors (λ) are reported in each panel.

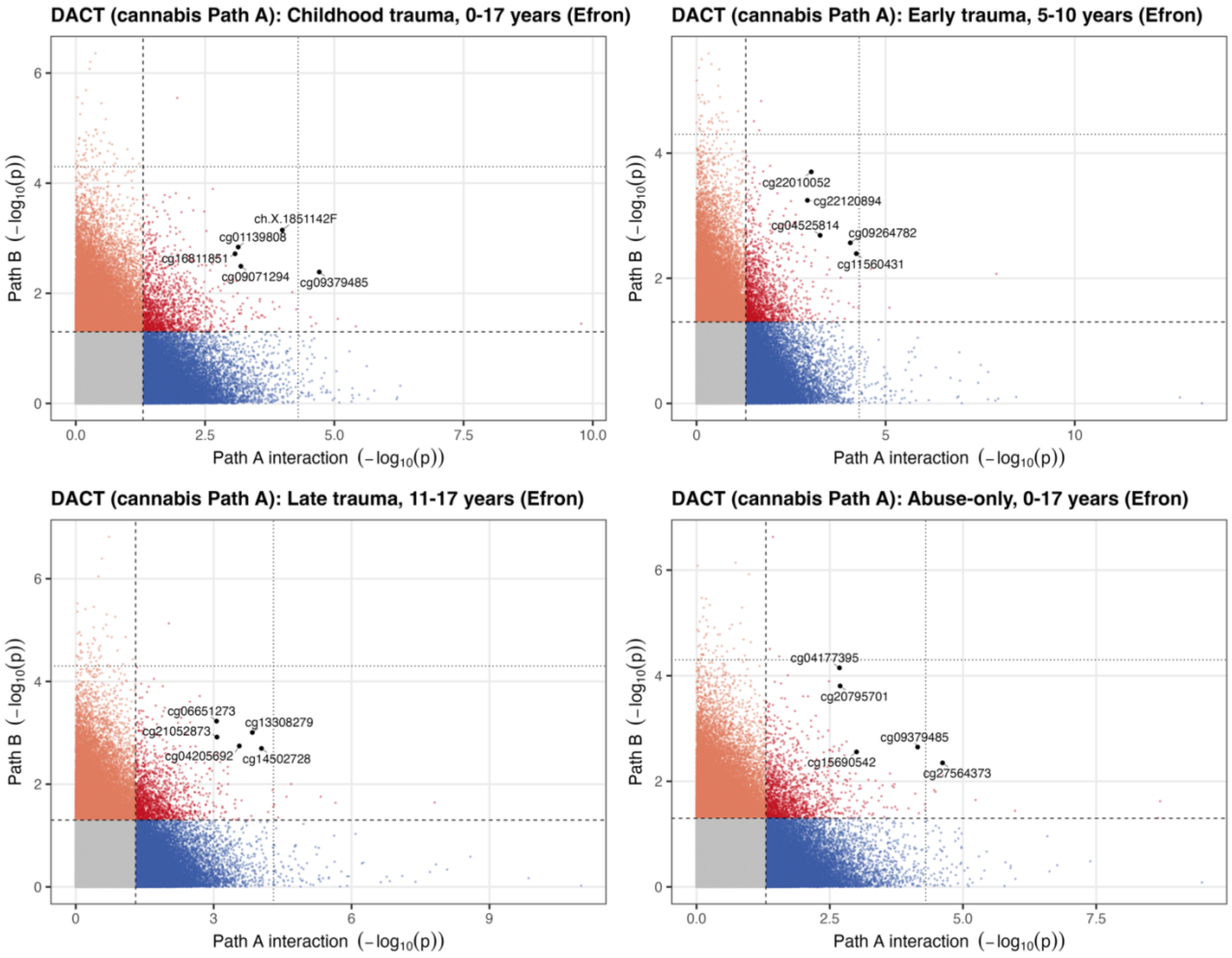


**Figure S15.** *Quadrant plots of CpG-level associations for Cannabis-moderated Path A and Path B.*
Each point represents a CpG site plotted according to the -$\log_{10}$-transformed p-values from the Path A model and the Path B model. Dashed lines indicate the nominal significance thresholds (p = 0.05). points are coloured according to significance pattern: both models, path A only, path B only, or neither. CpG sites showing suggestive mediation signal are highlighted and labelled.

**Table S14.** *Top CpG sites showing suggestive evidence of mediation*

| Exposure | Probe | Genomic location (hg19) | Gene | Location | Mediation effect | P-value | FDR |
|---|---|---|---|---|---|---|---|
| *Childhood trauma, 0-17 years* | *ch.X.1851142F* | X:128216049 | | OpenSea | -0.2173 | $3.5463 \times 10^{-5}$ | 0.9678 |
| *Late trauma, 11-17 years* | *cg13308279* | 2:197456953 | HECW2 | N_Shore | 0.1252 | $3.8818 \times 10^{-5}$ | 0.9725 |

Genomic locations are reported according to the hg19 reference genome. Gene annotation and CpG island context correspond to the Illumina HumanMethylation450 array annotation.
